# Conversational Artificial Intelligence-Based Integration of Clinical and Genomic Data Identifies MAPK Alterations in Colorectal Cancer

**DOI:** 10.64898/2025.12.18.25342607

**Authors:** Fernando C. Diaz, Brigette Waldrup, Francisco G. Carranza, Sophia Manjarrez, Enrique Velazquez-Villarreal

## Abstract

**Background:** Colorectal cancer (CRC) exhibits marked heterogeneity across age, ancestry, and treatment context, underscored by the rising incidence of early-onset disease (EOCRC). The mitogen-activated protein kinase (MAPK) signaling axis is a central regulator of CRC biology and treatment response, yet the frequency, distribution, and prognostic relevance of MAPK pathway alterations across demographically and clinically defined subgroups remain unclear.

**Methods:** We analyzed 2,515 CRC tumors with harmonized clinical, genomic, and treatment metadata. Patients were stratified by ancestry (H/L vs. non-Hispanic White [NHW]), age at diagnosis (early-onset vs. late-onset), and FOLFOX chemotherapy exposure. Somatic MAPK alterations were identified using a curated gene set spanning canonical and regulatory components of EGFR-RAS-RAF-MEK-ERK signaling. Conversational artificial intelligence (AI-HOPE and AI-HOPE-MAPK) enabled cohort construction and exploratory analytics using natural language queries, with all findings validated using standard statistical methods. Overall survival (OS) associations were evaluated using Kaplan-Meier analyses.

**Results:** MAPK pathway alterations demonstrated marked heterogeneity across ancestry and treatment contexts. Among EO H/L patients, FGFR3, NF1, and RPS6KA6 mutations were significantly more frequent in those not treated with FOLFOX, while PDGFRB alterations were enriched in EO H/L patients receiving FOLFOX compared with EO NHW counterparts. In late-onset H/L disease, NTRK2 and PDGFRB mutations were more prevalent in non-FOLFOX-treated tumors. Distinct MAPK-related alterations were also observed among NHW patients, including enrichment of AKT3, FGF4, RRAS2, CRKL, DUSP4, JUN, MAPK1, RRAS, and SOS1 mutations in non–FOLFOX-treated groups, as well as age-dependent differences in BRAF and TP53 mutation frequencies. Survival analyses revealed borderline evidence that MAPK pathway alterations were associated with improved overall survival in select NHW subgroups, including EOCRC patients treated with FOLFOX. Conversational AI enabled rapid generation and validation of multi-parameter queries.

**Conclusions:** Although MAPK pathway disruption is a near-universal molecular feature in CRC, the specific genes, effectors, and regulatory nodes altered vary by ancestry, age of onset, and chemotherapy exposure, revealing biologically and clinically meaningful substructure that is not captured by pathway-level status. NF1, MAPK3, RPS6KA4, and PDGFRB emerge as candidate genomic biomarkers in EOCRC and in H/L populations, while BRAF and adaptor-mediated signaling alterations are enriched in late-onset CRC. Conversational AI significantly accelerated analytic throughput and supported equity-driven biomarker discovery. Future studies incorporating multi-omic data and diverse prospective cohorts will be essential to refine MAPK-based precision strategies and address population-level disproportions in CRC.

## 1. Introduction

Colorectal cancer (CRC) remains a leading cause of cancer morbidity and mortality worldwide, yet its clinical course is increasingly heterogeneous across age groups, tumor biology, and population backgrounds (1,2). A particularly concerning shift is the continued rise of early-onset colorectal cancer (EOCRC), which reinforces the need for biomarker strategies that are both precision-oriented and equity-aware, especially for populations that have been historically underrepresented in genomic research and clinical trial datasets (1–3). Despite expanding access to next-generation sequencing and targeted therapies, the molecular determinants that shape treatment response and outcomes across clinically relevant subgroups remain incompletely defined (1,3,4).

A central axis in CRC pathobiology is the EGFR-RAS-RAF-MEK-ERK (MAPK) signaling cascade, which regulates proliferation, survival, invasion, and therapy resistance (5, 6). MAPK biology is tightly linked to the evolution of personalized CRC care: anti-EGFR monoclonal antibodies (cetuximab and panitumumab) represented an early breakthrough in targeted therapy, and biomarker-driven selection, particularly expanded RAS testing and the recognition of BRAF-driven disease biology, has improved patient stratification and reduced ineffective treatment exposure (4, 7–11). However, even within biomarker-selected populations, response to anti-EGFR therapy remains variable, highlighting the need for additional predictive biomarkers and a deeper mechanistic understanding of resistance beyond canonical RAS and BRAF alterations (7, 9, 11, 12).

The MAPK pathway is also implicated in chemoresistance to CRC standard-of-care regimens, including oxaliplatin- and fluoropyrimidine-based therapies such as FOLFOX (13–15). Mechanistic studies have shown that MAPK/ERK activation can promote epithelial-mesenchymal transition (EMT) and oxaliplatin resistance (16), while p38 MAPK signaling can facilitate survival programs in oxaliplatin-resistant CRC models and contribute to escape from apoptosis (13,17). These findings underscore that MAPK activity is not merely a downstream readout of oncogenic drivers, it is dynamically shaped by therapeutic pressures and pathway cross-talk (14, 15). For example, inhibition of mTORC1 can trigger compensatory MAPK activation through a PI3K-dependent feedback loop, supporting the rationale for combination strategies that anticipate adaptive signaling (18). Complementing these mechanistic insights, clinical correlative studies have linked MAPK-associated epigenetic regulation and signaling states to outcomes in patients treated with FOLFOX-based therapy, suggesting potential biomarker roles that are context dependent (19).

Meanwhile, therapeutic targeting of the MAPK pathway in CRC has advanced but remains challenging (20–24). Although RAF and MEK inhibitors have transformed outcomes in several RAF-driven malignancies, CRC displays complex intrinsic and adaptive resistance mechanisms that can limit durability of response (25–30). Contemporary CRC precision oncology emphasizes that effective MAPK-directed strategies will require improved molecular stratification, rational combinations, and biomarker discovery beyond single-gene predictors (7,11,31,32). These challenges are further complicated in real-world settings, where treatment patterns vary (e.g., FOLFOX, CAPEOX, FOLFOXIRI) and where subgroup-specific evidence, particularly in underrepresented populations, remains limited (31,33–35).

A practical barrier to progress is not only biological complexity, but also analytic complexity: actionable insights often require integrating age of onset, treatment exposure, clinicopathologic features, and ancestry or population descriptors with high-dimensional genomic features (1,2,3). Traditional analytic workflows are rigorous but can be slow to iterate when investigators need to test multi-parameter hypotheses across stratified cohorts. This limitation is especially relevant for disparity-focused precision oncology, where subgroup-aware analyses are essential and sample sizes can be uneven across strata.

To address these gaps, we apply a conversational artificial intelligence (AI) framework, AI-HOPE (36) and its pathway-specific module, AI-HOPE-MAPK (37), to accelerate clinical-genomic integration and hypothesis testing through natural language-driven queries. In this study, we characterize MAPK pathway alterations across 2,515 CRC cases, stratified by ancestry (H/L vs NHW), age of onset (early vs late), and FOLFOX exposure, and we assess associations with survival outcomes (38–40). By combining conventional statistical testing with AI-enabled cohort interrogation, we aim to clarify how MAPK alterations vary by clinical context and treatment status, and to establish an efficient, reproducible approach for biomarker discovery that supports precision medicine for populations historically underrepresented in cancer genomics research.

## 2. Materials and Methods

### 2.1 Study design and data provenance

This study employed a retrospective, integrative analysis of de-identified colorectal cancer (CRC) cases aggregated from large-scale, publicly available cancer genomics resources. Clinical, genomic, demographic, and treatment-level data were obtained from harmonized datasets hosted on cBioPortal, including The Cancer Genome Atlas (TCGA) Colorectal Adenocarcinoma (PanCancer Atlas), MSK-CHORD (2024), and the AACR Project GENIE Biopharma Collaborative (BPC) CRC cohort. These datasets were selected for their depth of somatic variant annotation, availability of longitudinal clinical metadata, and representation of real-world treatment exposure.

Eligible cases included patients with histopathologically confirmed colorectal adenocarcinoma and available tumor-based next-generation sequencing data. When multiple sequencing records were present for a single individual, one representative sample was retained to prevent duplicate inclusion.

### 2.2 Population stratification and clinical subgroup definitions

Patients were stratified by ancestry, age at diagnosis, and chemotherapy exposure. Hispanic/Latino (H/L) ancestry was determined using self-reported ethnicity fields when available. In datasets lacking explicit ethnicity annotation, probabilistic surname-based inference methods were applied to identify likely H/L ancestry. The comparator population consisted of non-Hispanic White (NHW) patients meeting identical inclusion criteria.

EOCRC was defined as diagnosis prior to 50 years of age, while late-onset colorectal cancer (LOCRC) was defined as diagnosis at age 50 years or older. These definitions were applied uniformly across all datasets to ensure consistency.

### 2.3 Treatment exposure annotation and FOLFOX classification

Chemotherapy exposure was abstracted from structured treatment variables and curated medication records. Patients were classified as FOLFOX-treated if documented exposure included all three regimen components (5-fluorouracil, leucovorin, and oxaliplatin) administered within the same therapeutic line or within an overlapping treatment window consistent with standard FOLFOX protocols.

Patients lacking documentation of all three agents, or who received partial or alternative regimens (e.g., single-agent fluoropyrimidine, oxaliplatin alone, or non-overlapping administration), were categorized as non-FOLFOX. Treatment timing relative to sequencing was reviewed to confirm relevance to tumor molecular profiles.

### 2.4 MAPK pathway curation and genomic alteration assessment

A curated MAPK signaling gene set was assembled by integrating canonical pathway definitions from KEGG and Reactome with contemporary CRC-focused literature describing MAPK-driven tumorigenesis, therapy resistance, and pathway cross-talk. This gene set encompassed upstream receptor tyrosine kinases, core RAS/RAF/MEK/ERK components, MAPK effector kinases, and regulatory feedback nodes.

Somatic mutation data were extracted from mutation annotation format (MAF) files and equivalent variant tables. Only protein-altering variants, including missense, nonsense, frameshift, splice-site, and start-loss mutations, were retained for analysis. MAPK pathway alteration status was defined at the patient level as the presence of at least one qualifying somatic alteration in any MAPK-associated gene.

### 2.5 Statistical analyses

Comparisons of MAPK alteration frequencies across ancestry, age-of-onset, and treatment-defined subgroups were performed using Fisher’s exact test or chi-square test, as appropriate based on cell counts. Continuous clinical variables were evaluated using non-parametric tests.

Overall survival (OS) was defined as the interval from diagnosis or sequencing (depending on dataset availability) to death or last follow-up. Survival distributions were estimated using Kaplan-Meier methods, and group-level differences were assessed using the log-rank test. Exploratory Cox proportional hazards models were constructed to estimate hazard ratios with 95% confidence intervals, adjusting for available covariates including age, sex, tumor site, microsatellite instability (MSI) status, and chemotherapy exposure. All statistical analyses were conducted using R (version 4.3), with two-sided p-values <0.05 considered statistically significant.

### 2.6 Conversational AI-enabled integration and analytic workflow

To complement traditional statistical workflows, we implemented AI-HOPE and its pathway-specific module AI-HOPE-MAPK, conversational artificial intelligence platforms designed to integrate clinical, genomic, demographic, and treatment variables within a unified analytic environment. These systems enable natural language-driven cohort construction, pathway-specific mutation querying, and rapid subgroup stratification across high-dimensional datasets.

AI-HOPE-MAPK was used to Identify patients meeting compound criteria combining ancestry, age of onset, treatment exposure, and MAPK alteration status; generate pathway-level mutation frequency summaries stratified by clinical and demographic variables; perform iterative subgroup exploration to prioritize candidate ancestry- and treatment-associated MAPK alterations; and facilitate reproducible analytic logic through transparent, query-based workflows.

All AI-assisted outputs were cross-validated against conventional data extracts and independently confirmed using standard statistical analyses. The AI platform was used as an analytic accelerator rather than a replacement for statistical inference, ensuring reproducibility, interpretability, and methodological transparency.

## 3. Results

### 3.1 Cohort composition and baseline clinical characteristics

Baseline demographic, clinical, and molecular features of the study population are detailed in Table 1. The final analytic cohort comprised 2,515 colorectal cancer (CRC) cases, including 266 H/L (10.6%) and 2,249 NHW (89.4%) patients. Distinct distributions were observed across age-of-onset and treatment-defined subgroups, underscoring population-specific differences in clinical presentation and therapeutic exposure.

**Table 1.** Baseline Demographic, Clinical, and Molecular Characteristics of the Study Cohort.

| Category | Variable | Hispanic/Latino (H/L) n = 266 | Non-Hispanic White (NHW) n = 2,249 |
| --- | --- | --- | --- |
| **Age at Diagnosis & FOLFOX Exposure** | EOCRC, FOLFOX-treated | 73 (27.4%) | 375 (16.7%) |
|  | EOCRC, not FOLFOX-treated | 52 (19.5%) | 302 (13.4%) |
|  | LOCRC, FOLFOX-treated | 91 (34.2%) | 919 (40.9%) |
|  | LOCRC, not FOLFOX-treated | 50 (18.8%) | 653 (29.0%) |
| **Tumor Site** | Colon adenocarcinoma | 164 (61.7%) | 1,328 (59.0%) |
|  | Rectal adenocarcinoma | 64 (24.1%) | 646 (28.7%) |
|  | Colorectal adenocarcinoma (NOS) | 38 (14.3%) | 275 (12.2%) |
| **Sex** | Male | 158 (59.4%) | 1,267 (56.3%) |
|  | Female | 108 (40.6%) | 982 (43.7%) |
| **Tumor Sample Source** | Primary tumor | 266 (100%) | 2,249 (100%) |
| **Clinical Stage at Diagnosis** | Stage I-III | 156 (58.6%) | 1,236 (55.0%) |
|  | Stage IV | 108 (40.6%) | 1,005 (44.7%) |
|  | Missing | 2 (0.8%) | 8 (0.4%) |
| **Microsatellite Status** | MSS | 200 (75.2%) | 1,940 (86.3%) |
|  | MSI-H | 21 (7.9%) | 209 (9.3%) |
|  | Indeterminate | 10 (3.8%) | 57 (2.5%) |
|  | Missing | 35 (13.2%) | 43 (1.9%) |
| **Ethnicity Annotation** | Spanish/Hispanic NOS | 230 (86.5%) | — |
|  | Mexican / Chicano | 30 (11.3%) | — |
|  | Other Hispanic categories* | 6 (2.3%) | — |
|  | Non-Hispanic White | — | 2,249 (100%) |

\*Includes “Hispanic or Latino,” “Other Spanish/Hispanic,” and surname-inferred Hispanic ancestry.

When stratified by age at diagnosis and FOLFOX exposure, H/L patients showed a greater proportional burden of early-onset disease. Specifically, EOCRC treated with FOLFOX accounted for 27.4% of H/L cases compared with 16.7% among NHW patients, while EOCRC not exposed to FOLFOX was also more frequent in the H/L cohort (19.5% vs. 13.4%). In contrast, NHW patients were more commonly represented among late-onset CRC (LOCRC) receiving FOLFOX (40.9% vs. 34.2%) and LOCRC not treated with FOLFOX (29.0% vs. 18.8%), reflecting differential treatment patterns by age and ancestry.

Tumor anatomic distributions were similar across groups. Colon adenocarcinoma was the predominant diagnosis in both populations (61.7% H/L; 59.0% NHW), followed by rectal adenocarcinoma and colorectal adenocarcinoma not otherwise specified, with only modest proportional variation between cohorts. Sex distributions were likewise comparable, with males representing 59.4% of H/L and 56.3% of NHW cases. All tumors included in the analysis were derived from primary tumor specimens.

Clinical stage at diagnosis demonstrated broadly aligned patterns, although NHW patients exhibited a slightly higher proportion of stage IV disease at presentation (44.7%) compared with H/L patients (40.6%). Microsatellite status differed between groups, with microsatellite-stable (MSS) tumors more prevalent in the NHW cohort (86.3%) than in the H/L cohort (75.2%), while MSI-high tumors comprised a smaller fraction of both populations.

Ethnicity annotations reflected the cohort construction strategy. The H/L group was primarily composed of patients categorized as Spanish/Hispanic NOS (86.5%), with additional representation from Mexican/Chicano (11.3%) and other Hispanic subcategories. As expected, all individuals in the NHW cohort were annotated as non-Hispanic White. Together, these baseline characteristics provide the clinical context for subsequent analyses of ancestry-, age-, and treatment-associated MAPK pathway alterations.

### 3.2 Clinical and Genomic Stratification

#### 3.2.1 Hispanic/Latino CRC by Age of Onset and FOLFOX Exposure

To further dissect ancestry-specific heterogeneity within the H/L cohort, we performed a stratified analysis comparing EOCRC and LOCRC patients according to FOLFOX treatment status (Table 2a). This analysis integrated clinical metrics with MAPK-related genomic features to evaluate how age and chemotherapy exposure shape tumor biology within this disproportionately affected population.

**Table 2.**
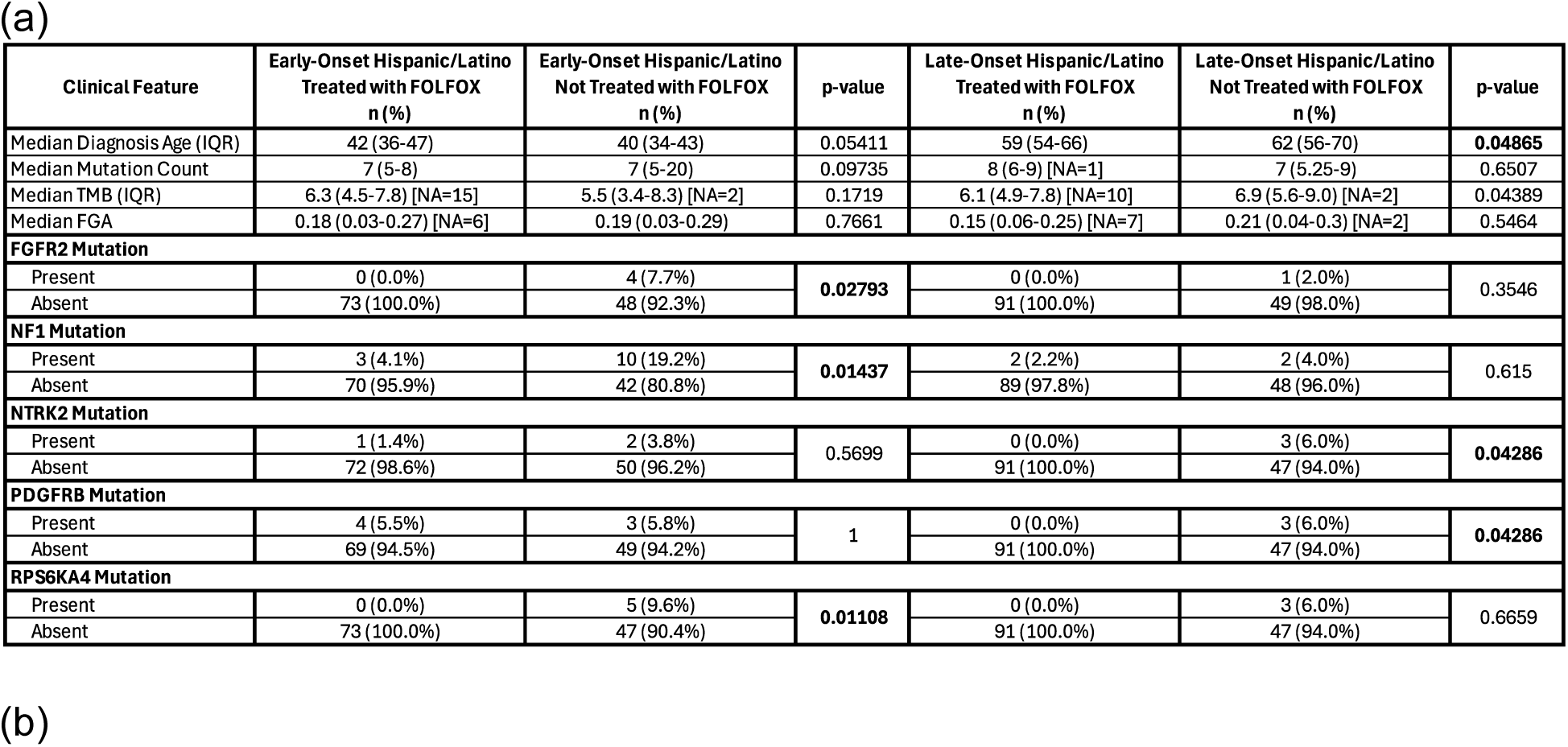
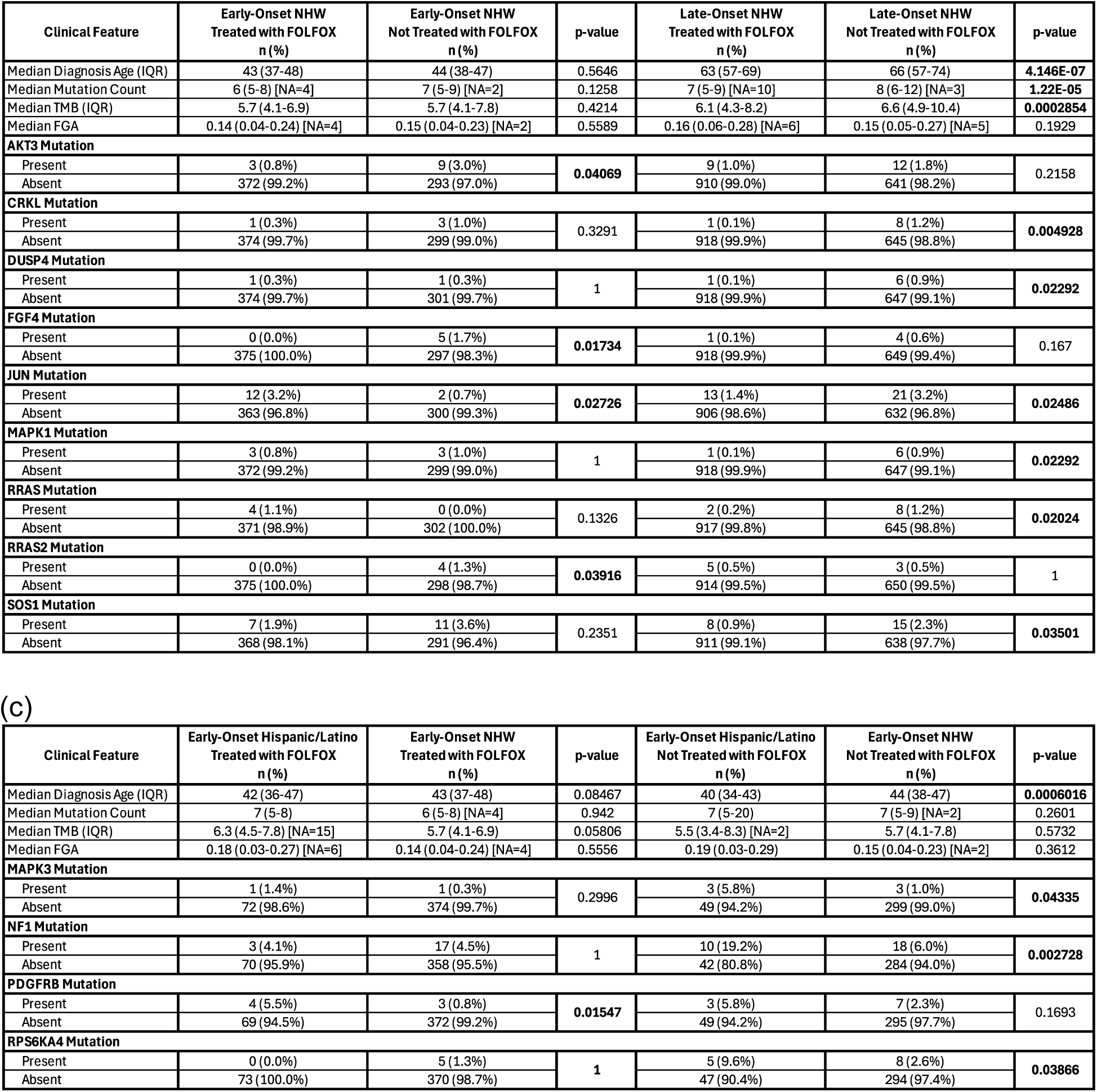
Age-, Ancestry-, and Treatment-Stratified Clinical and MAPK Genomic Features in Colorectal Cancer. This table summarizes key clinical parameters and MAPK pathway-related genomic characteristics across colorectal cancer subgroups defined by age at diagnosis, ancestry, and FOLFOX treatment exposure. Analyses are presented across four complementary comparisons: (a) early-onset versus late-onset disease within Hispanic/Latino (H/L) patients; (b) early-onset versus late-onset disease within non-Hispanic White (NHW) patients; and (c) ancestry-based comparisons among early-onset colorectal cancer (EOCRC) cases. Reported variables include median age at diagnosis, overall mutation burden, TMB, fraction of genome altered, and the frequency of selected MAPK signaling pathway gene alterations.

| Clinical Feature | Early-Onset Hispanic/Latino<br>Treated with FOLFOX<br>n (%) | Early-Onset Hispanic/Latino<br>Not Treated with FOLFOX<br>n (%) | p-value | Late-Onset Hispanic/Latino<br>Treated with FOLFOX<br>n (%) | Late-Onset Hispanic/Latino<br>Not Treated with FOLFOX<br>n (%) | p-value |
| --- | --- | --- | --- | --- | --- | --- |
| Median Diagnosis Age (IQR) | 42 (36-47) | 40 (34-43) | 0.05411 | 59 (54-66) | 62 (56-70) | 0.04865 |
| Median Mutation Count | 7 (5-8) | 7 (5-20) | 0.09735 | 8 (6-9) [NA=1] | 7 (5.25-9) | 0.6507 |
| Median TMB (IQR) | 6.3 (4.5-7.8) [NA=15] | 5.5 (3.4-8.3) [NA=2] | 0.1719 | 6.1 (4.9-7.8) [NA=10] | 6.9 (5.6-9.0) [NA=2] | 0.04389 |
| Median FGA | 0.18 (0.03-0.27) [NA=6] | 0.19 (0.03-0.29) | 0.7661 | 0.15 (0.06-0.25) [NA=7] | 0.21 (0.04-0.3) [NA=2] | 0.5464 |
| FGFR2 Mutation |  |  |  |  |  |  |
| Present | 0 (0.0%) | 4 (7.7%) | 0.02793 | 0 (0.0%) | 1 (2.0%) | 0.3546 |
| Absent | 73 (100.0%) | 48 (92.3%) |  | 91 (100.0%) | 49 (98.0%) |  |
| NF1 Mutation |  |  |  |  |  |  |
| Present | 3 (4.1%) | 10 (19.2%) | 0.01437 | 2 (2.2%) | 2 (4.0%) | 0.615 |
| Absent | 70 (95.9%) | 42 (80.8%) |  | 89 (97.8%) | 48 (96.0%) |  |
| NTRK2 Mutation |  |  |  |  |  |  |
| Present | 1 (1.4%) | 2 (3.8%) | 0.5699 | 0 (0.0%) | 3 (6.0%) | 0.04286 |
| Absent | 72 (98.6%) | 50 (96.2%) |  | 91 (100.0%) | 47 (94.0%) |  |
| PDGFRB Mutation |  |  |  |  |  |  |
| Present | 4 (5.5%) | 3 (5.8%) | 1 | 0 (0.0%) | 3 (6.0%) | 0.04286 |
| Absent | 69 (94.5%) | 49 (94.2%) |  | 91 (100.0%) | 47 (94.0%) |  |
| RPS6KA4 Mutation |  |  |  |  |  |  |
| Present | 0 (0.0%) | 5 (9.6%) | 0.01108 | 0 (0.0%) | 3 (6.0%) | 0.6659 |
| Absent | 73 (100.0%) | 47 (90.4%) |  | 91 (100.0%) | 47 (94.0%) |  |

(b)
| Clinical Feature | Early-Onset NHW<br>Treated with FOLFOX<br>n (%) | Early-Onset NHW<br>Not Treated with FOLFOX<br>n (%) | p-value | Late-Onset NHW<br>Treated with FOLFOX<br>n (%) | Late-Onset NHW<br>Not Treated with FOLFOX<br>n (%) | p-value |
| --- | --- | --- | --- | --- | --- | --- |
| Median Diagnosis Age (IQR) | 43 (37-48) | 44 (38-47) | 0.5646 | 63 (57-69) | 66 (57-74) | 4.146E-07 |
| Median Mutation Count | 6 (5-8) [NA=4] | 7 (5-9) [NA=2] | 0.1258 | 7 (5-9) [NA=10] | 8 (6-12) [NA=3] | 1.22E-05 |
| Median TMB (IQR) | 5.7 (4.1-6.9) | 5.7 (4.1-7.8) | 0.4214 | 6.1 (4.3-8.2) | 6.6 (4.9-10.4) | 0.0002854 |
| Median FGA | 0.14 (0.04-0.24) [NA=4] | 0.15 (0.04-0.23) [NA=2] | 0.5589 | 0.16 (0.06-0.28) [NA=6] | 0.15 (0.05-0.27) [NA=5] | 0.1929 |
| AKT3 Mutation |  |  |  |  |  |  |
| Present | 3 (0.8%) | 9 (3.0%) | 0.04069 | 9 (1.0%) | 12 (1.8%) | 0.2158 |
| Absent | 372 (99.2%) | 293 (97.0%) |  | 910 (99.0%) | 641 (98.2%) |  |
| CRKL Mutation |  |  |  |  |  |  |
| Present | 1 (0.3%) | 3 (1.0%) | 0.3291 | 1 (0.1%) | 8 (1.2%) | 0.004928 |
| Absent | 374 (99.7%) | 299 (99.0%) |  | 918 (99.9%) | 645 (98.8%) |  |
| DUSP4 Mutation |  |  |  |  |  |  |
| Present | 1 (0.3%) | 1 (0.3%) | 1 | 1 (0.1%) | 6 (0.9%) | 0.02292 |
| Absent | 374 (99.7%) | 301 (99.7%) |  | 918 (99.9%) | 647 (99.1%) |  |
| FGF4 Mutation |  |  |  |  |  |  |
| Present | 0 (0.0%) | 5 (1.7%) | 0.01734 | 1 (0.1%) | 4 (0.6%) | 0.167 |
| Absent | 375 (100.0%) | 297 (98.3%) |  | 918 (99.9%) | 649 (99.4%) |  |
| JUN Mutation |  |  |  |  |  |  |
| Present | 12 (3.2%) | 2 (0.7%) | 0.02726 | 13 (1.4%) | 21 (3.2%) | 0.02486 |
| Absent | 363 (96.8%) | 300 (99.3%) |  | 906 (98.6%) | 632 (96.8%) |  |
| MAPK1 Mutation |  |  |  |  |  |  |
| Present | 3 (0.8%) | 3 (1.0%) | 1 | 1 (0.1%) | 6 (0.9%) | 0.02292 |
| Absent | 372 (99.2%) | 299 (99.0%) |  | 918 (99.9%) | 647 (99.1%) |  |
| RRAS Mutation |  |  |  |  |  |  |
| Present | 4 (1.1%) | 0 (0.0%) | 0.1326 | 2 (0.2%) | 8 (1.2%) | 0.02024 |
| Absent | 371 (98.9%) | 302 (100.0%) |  | 917 (99.8%) | 645 (98.8%) |  |
| RRAS2 Mutation |  |  |  |  |  |  |
| Present | 0 (0.0%) | 4 (1.3%) | 0.03916 | 5 (0.5%) | 3 (0.5%) | 1 |
| Absent | 375 (100.0%) | 298 (98.7%) |  | 914 (99.5%) | 650 (99.5%) |  |
| SOS1 Mutation |  |  |  |  |  |  |
| Present | 7 (1.9%) | 11 (3.6%) | 0.2351 | 8 (0.9%) | 15 (2.3%) | 0.03501 |
| Absent | 368 (98.1%) | 291 (96.4%) |  | 911 (99.1%) | 638 (97.7%) |  |

(c)
| Clinical Feature | Early-Onset Hispanic/Latino<br>Treated with FOLFOX<br>n (%) | Early-Onset NHW<br>Treated with FOLFOX<br>n (%) | p-value | Early-Onset Hispanic/Latino<br>Not Treated with FOLFOX<br>n (%) | Early-Onset NHW<br>Not Treated with FOLFOX<br>n (%) | p-value |
| --- | --- | --- | --- | --- | --- | --- |
| Median Diagnosis Age (IQR) | 42 (36-47) | 43 (37-48) | 0.08467 | 40 (34-43) | 44 (38-47) | 0.0006016 |
| Median Mutation Count | 7 (5-8) | 6 (5-8) [NA=4] | 0.942 | 7 (5-20) | 7 (5-9) [NA=2] | 0.2601 |
| Median TMB (IQR) | 6.3 (4.5-7.8) [NA=15] | 5.7 (4.1-6.9) | 0.05806 | 5.5 (3.4-8.3) [NA=2] | 5.7 (4.1-7.8) | 0.5732 |
| Median FGA | 0.18 (0.03-0.27) [NA=6] | 0.14 (0.04-0.24) [NA=4] | 0.5556 | 0.19 (0.03-0.29) | 0.15 (0.04-0.23) [NA=2] | 0.3612 |
| MAPK3 Mutation |  |  |  |  |  |  |
| Present | 1 (1.4%) | 1 (0.3%) | 0.2996 | 3 (5.8%) | 3 (1.0%) | 0.04335 |
| Absent | 72 (98.6%) | 374 (99.7%) |  | 49 (94.2%) | 299 (99.0%) |  |
| NF1 Mutation |  |  |  |  |  |  |
| Present | 3 (4.1%) | 17 (4.5%) | 1 | 10 (19.2%) | 18 (6.0%) | 0.002728 |
| Absent | 70 (95.9%) | 358 (95.5%) |  | 42 (80.8%) | 284 (94.0%) |  |
| PDGFRB Mutation |  |  |  |  |  |  |
| Present | 4 (5.5%) | 3 (0.8%) | 0.01547 | 3 (5.8%) | 7 (2.3%) | 0.1693 |
| Absent | 69 (94.5%) | 372 (99.2%) |  | 49 (94.2%) | 295 (97.7%) |  |
| RPS6KA4 Mutation |  |  |  |  |  |  |
| Present | 0 (0.0%) | 5 (1.3%) | 1 | 5 (9.6%) | 8 (2.6%) | 0.03866 |
| Absent | 73 (100.0%) | 370 (98.7%) |  | 47 (90.4%) | 294 (97.4%) |  |

Across both age strata, median age at diagnosis differed modestly by treatment status. Among EO H/L patients, those treated with FOLFOX were diagnosed slightly later than their non-FOLFOX-treated counterparts (median 42 vs. 40 years), approaching statistical significance (p = 0.054). In contrast, a significant age difference was observed in LO H/L patients, with FOLFOX-treated individuals diagnosed at a younger median age compared with those not receiving FOLFOX (59 vs. 62 years; p = 0.049), suggesting potential age-related treatment selection effects in older patients.

Global measures of TMB showed nuanced differences by age and treatment. Median mutation counts were comparable between EO H/L patients regardless of FOLFOX exposure, while LO H/L patients exhibited similar mutation burdens across treatment groups. TMB demonstrated greater variability in LOCRC: LO H/L patients not treated with FOLFOX had a significantly higher median TMB compared with those receiving FOLFOX (6.9 vs. 6.1 mutations/Mb; p = 0.044). In contrast, TMB differences among EO H/L patients did not reach statistical significance. Fraction of genome altered (FGA) did not differ meaningfully by treatment status in either age group.

At the gene-specific level, several MAPK pathway alterations exhibited strong age- and treatment-dependent patterns. In EO H/L patients, FGFR2 mutations were observed exclusively in non-FOLFOX-treated tumors (7.7% vs. 0.0%; p = 0.028). Similarly, NF1 alterations were significantly enriched among EO H/L patients not treated with FOLFOX (19.2%) compared with those receiving FOLFOX (4.1%; p = 0.014). RPS6KA4 mutations, another MAPK effector, followed a similar pattern, occurring only in EO H/L tumors not exposed to FOLFOX (9.6% vs. 0.0%; p = 0.011).

Distinct trends emerged in late-onset disease. While FGFR2 and NF1 mutations were infrequent and not significantly different by treatment status in LO H/L patients, NTRK2 and PDGFRB alterations were significantly enriched in LO H/L tumors not treated with FOLFOX (each 6.0% vs. 0.0%; p = 0.043). These findings suggest divergent MAPK pathway dependencies between EOCRC and LOCRC within the same ancestral group.

These results demonstrate that MAPK pathway alterations in H/L CRC are strongly modulated by both age of onset and chemotherapy exposure, with distinct gene-level enrichment patterns in early- versus late-onset disease. This stratified genomic landscape underscores the importance of age- and treatment-aware analyses when evaluating precision oncology biomarkers in underrepresented populations and provides a foundation for subsequent ancestry-comparative and survival analyses.

#### 3.2.2 Age-, Treatment-, and MAPK-Specific Genomic Patterns in Non-Hispanic White Patients

To delineate MAPK-associated clinical-genomic heterogeneity within the NHW cohort, we examined EOCRC and LOCRC cases stratified by FOLFOX exposure (Table 2b). This analysis integrated age at diagnosis, global mutational metrics, and gene-level MAPK alterations to assess how treatment context intersects with tumor biology in NHW patients.

Among EOCRC cases, median age at diagnosis was comparable between FOLFOX-treated and non-FOLFOX-treated patients (43 vs. 44 years; p = 0.56), indicating minimal age-based selection for chemotherapy in younger NHW individuals. In contrast, LOCRC patients demonstrated a pronounced age difference by treatment status: those receiving FOLFOX were diagnosed significantly earlier than their non-treated counterparts (median 63 vs. 66 years; p = 4.1 × 10⁻⁷), suggesting that age may influence chemotherapy utilization in older NHW populations.

Global mutational measures revealed limited differences in EOCRC but clear treatment-associated divergence in LOCRC. Median mutation counts and TMB were similar between treatment groups in EOCRC. However, among LOCRC patients, non-FOLFOX-treated tumors exhibited significantly higher median mutation counts (8 vs. 7; p = 1.2 × 10⁻⁵) and elevated TMB (6.6 vs. 6.1 mutations/Mb; p = 2.9 × 10⁻⁴) compared with FOLFOX-treated tumors. Fraction of genome altered (FGA) did not differ significantly by treatment status in either age group.

At the gene-specific level, multiple MAPK pathway components showed age- and treatment-dependent enrichment patterns. In EOCRC, AKT3, FGF4, JUN, and RRAS2 mutations were significantly more frequent in tumors not treated with FOLFOX, whereas JUN alterations were enriched in FOLFOX-treated EOCRC tumors (3.2% vs. 0.7%; p = 0.027), suggesting distinct MAPK signaling contexts associated with chemotherapy exposure in younger patients.

In LOCRC, treatment-associated differences were more prominent across several MAPK-related genes. Tumors not exposed to FOLFOX demonstrated significantly higher frequencies of CRKL, DUSP4, MAPK1, RRAS, and SOS1 mutations, while JUN alterations were again enriched in FOLFOX-treated tumors (1.4% vs. 3.2% in non-treated; p = 0.025). These patterns indicate that MAPK pathway perturbations in NHW CRC are not uniform but vary substantially with age and chemotherapy exposure.

These findings reveal that, within the NHW population, MAPK pathway alterations are strongly modulated by age of onset and FOLFOX treatment status, with relatively modest differences in EOCRC and more pronounced genomic divergence in LOCRC. This stratified landscape highlights the importance of incorporating treatment context into MAPK-focused biomarker analyses and provides a framework for subsequent ancestry-comparative and outcome-oriented investigations.

#### 3.2.4 Ancestry-Specific Genomic Features in Early-Onset Colorectal Cancer by Treatment Context

To directly compare ancestry-associated molecular differences in EOCRC, we evaluated H/L and NHW patients stratified by FOLFOX treatment status (Table 2c). This focused analysis allowed assessment of whether clinical and MAPK-related genomic features diverge by ethnicity within the same age-of-onset category.

Among FOLFOX-treated EOCRC cases, median age at diagnosis was similar between H/L and NHW patients (42 vs. 43 years; p = 0.085), indicating comparable age distributions at treatment initiation. Measures of global tumor complexity, including mutation count, TMB, and fraction of genome altered (FGA), were also broadly comparable between ancestries in this treatment group. However, gene-level differences within the MAPK pathway were evident. PDGFRB mutations were significantly more frequent in FOLFOX-treated H/L EOCRC tumors compared with NHW counterparts (5.5% vs. 0.8%; p = 0.015), suggesting ancestry-associated differences in receptor tyrosine kinase signaling among treated early-onset cases.

In contrast, more pronounced ancestry-related divergence was observed among EOCRC patients not treated with FOLFOX. H/L patients in this group were diagnosed at a significantly younger median age than NHW patients (40 vs. 44 years; p = 6.0 × 10⁻⁴), highlighting a substantial age disparity within untreated early-onset disease. Despite similar overall mutation counts, specific MAPK pathway alterations were markedly enriched in H/L tumors. MAPK3 mutations were significantly more common in H/L EOCRC not exposed to FOLFOX compared with NHW tumors (5.8% vs. 1.0%; p = 0.043). Likewise, NF1 alterations were strongly enriched in H/L patients relative to NHW patients (19.2% vs. 6.0%; p = 0.0027), underscoring ancestry-associated differences in negative regulation of MAPK signaling. RPS6KA4 mutations, a downstream MAPK effector, were also more frequent in H/L EOCRC not treated with FOLFOX (9.6% vs. 2.6%; p = 0.039).

These results demonstrate that ancestry-related differences in MAPK pathway alterations are most pronounced in EOCRC patients not exposed to FOLFOX, whereas treated EOCRC shows more limited divergence. The enrichment of MAPK3, NF1, RPS6KA4, and PDGFRB alterations in H/L patients suggests distinct MAPK signaling dependencies in early-onset disease that may be masked or modified by chemotherapy exposure. These results further support the need for ancestry- and treatment-aware molecular stratification in EOCRC precision oncology.

### 3.3 Distribution of MAPK Pathway Alterations Across Age, Ancestry, and Chemotherapy Context

To determine whether MAPK pathway alteration prevalence varies by age of onset, ancestry, or FOLFOX exposure, we compared pathway-level alteration frequencies across stratified colorectal cancer subgroups (Tables 3a-3d). In contrast to the gene-specific heterogeneity observed in earlier analyses, MAPK pathway alterations were highly prevalent across nearly all clinical strata, with only modest variation by treatment or ancestry.

#### 3.3.1 Age- and treatment-specific patterns within ancestry groups

Within the H/L cohort, MAPK pathway alterations were detected in the vast majority of tumors regardless of age or treatment status (Table 3a). Among early-onset cases, alterations were present in 95.9% of FOLFOX-treated tumors and all of the tumors not exposed to FOLFOX, with no statistically significant difference between groups. A similar pattern was observed in late-onset H/L disease, where MAPK alterations were identified in 97.8% of FOLFOX-treated tumors and all of the non-FOLFOX-treated tumors. These findings indicate that MAPK pathway involvement is nearly ubiquitous in H/L CRC, independent of age at diagnosis or chemotherapy exposure.

**Table 3.**
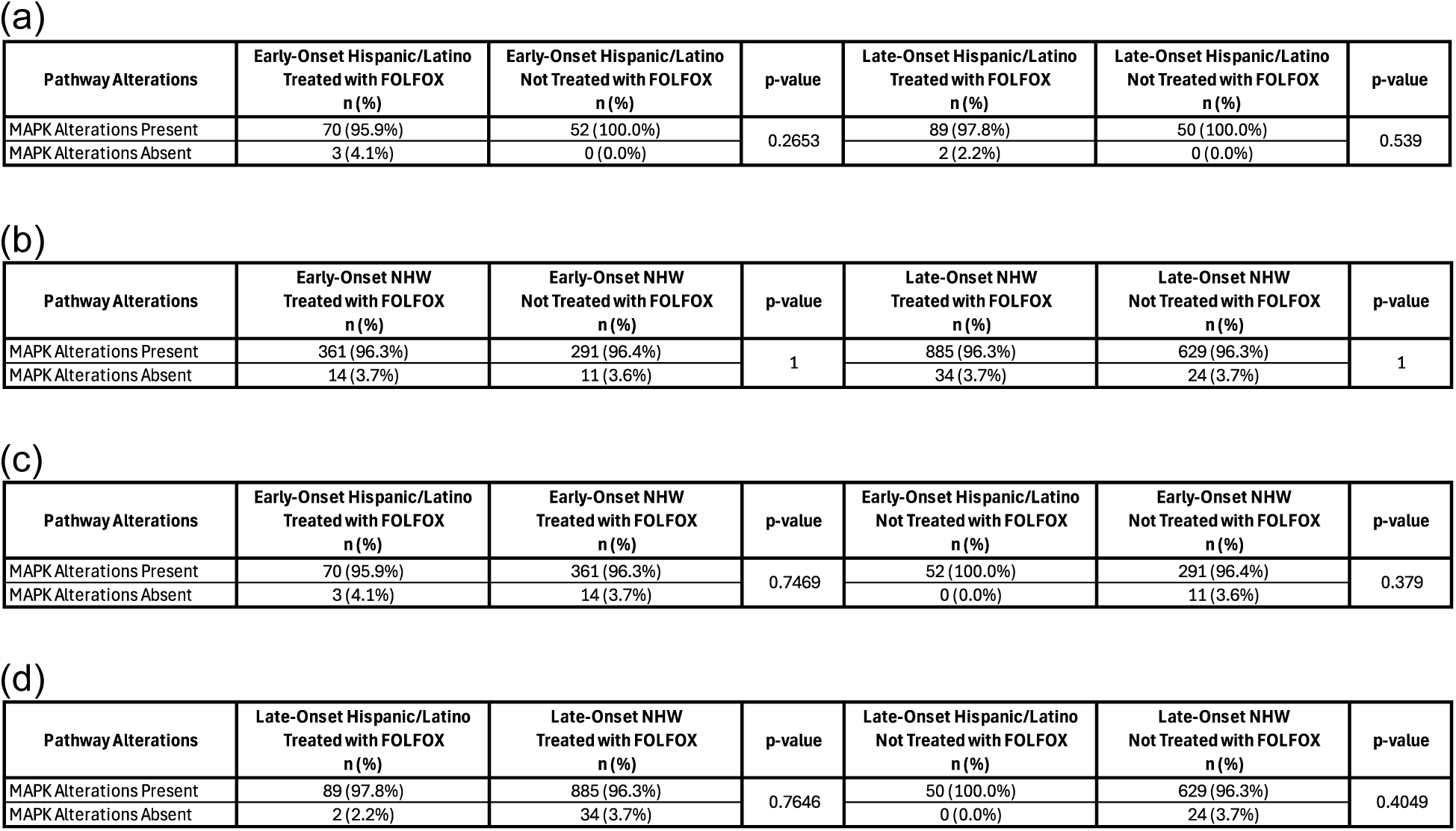
Prevalence of MAPK Pathway Alterations Across Age-, Ancestry-, and Treatment-Defined Colorectal Cancer Subgroups. This table summarizes the frequency of MAPK pathway alterations across colorectal cancer cohorts stratified by age at diagnosis (early-onset vs. late-onset), ancestry (Hispanic/Latino vs. non-Hispanic White), and FOLFOX chemotherapy exposure. Data are organized into four analytical panels: (a) within-ancestry comparisons of early- and late-onset disease among Hispanic/Latino patients by treatment status; (b) parallel age- and treatment-based comparisons within the non-Hispanic White cohort; (c) ancestry-based comparisons restricted to early-onset colorectal cancer, stratified by FOLFOX exposure; and (d) ancestry-based comparisons among late-onset colorectal cancer cases, also stratified by treatment status.

| Pathway Alterations | Early-Onset Hispanic/Latino<br>Treated with FOLFOX<br>n (%) | Early-Onset Hispanic/Latino<br>Not Treated with FOLFOX<br>n (%) | p-value | Late-Onset Hispanic/Latino<br>Treated with FOLFOX<br>n (%) | Late-Onset Hispanic/Latino<br>Not Treated with FOLFOX<br>n (%) | p-value |
| --- | --- | --- | --- | --- | --- | --- |
| MAPK Alterations Present | 70 (95.9%) | 52 (100.0%) | 0.2653 | 89 (97.8%) | 50 (100.0%) | 0.539 |
| MAPK Alterations Absent | 3 (4.1%) | 0 (0.0%) |  | 2 (2.2%) | 0 (0.0%) |  |

(b)
| Pathway Alterations | Early-Onset NHW<br>Treated with FOLFOX<br>n (%) | Early-Onset NHW<br>Not Treated with FOLFOX<br>n (%) | p-value | Late-Onset NHW<br>Treated with FOLFOX<br>n (%) | Late-Onset NHW<br>Not Treated with FOLFOX<br>n (%) | p-value |
| --- | --- | --- | --- | --- | --- | --- |
| MAPK Alterations Present | 361 (96.3%) | 291 (96.4%) | 1 | 885 (96.3%) | 629 (96.3%) | 1 |
| MAPK Alterations Absent | 14 (3.7%) | 11 (3.6%) |  | 34 (3.7%) | 24 (3.7%) |  |

(c)
| Pathway Alterations | Early-Onset Hispanic/Latino<br>Treated with FOLFOX<br>n (%) | Early-Onset NHW<br>Treated with FOLFOX<br>n (%) | p-value | Early-Onset Hispanic/Latino<br>Not Treated with FOLFOX<br>n (%) | Early-Onset NHW<br>Not Treated with FOLFOX<br>n (%) | p-value |
| --- | --- | --- | --- | --- | --- | --- |
| MAPK Alterations Present | 70 (95.9%) | 361 (96.3%) | 0.7469 | 52 (100.0%) | 291 (96.4%) | 0.379 |
| MAPK Alterations Absent | 3 (4.1%) | 14 (3.7%) |  | 0 (0.0%) | 11 (3.6%) |  |

(d)
| Pathway Alterations | Late-Onset Hispanic/Latino<br>Treated with FOLFOX<br>n (%) | Late-Onset NHW<br>Treated with FOLFOX<br>n (%) | p-value | Late-Onset Hispanic/Latino<br>Not Treated with FOLFOX<br>n (%) | Late-Onset NHW<br>Not Treated with FOLFOX<br>n (%) | p-value |
| --- | --- | --- | --- | --- | --- | --- |
| MAPK Alterations Present | 89 (97.8%) | 885 (96.3%) | 0.7646 | 50 (100.0%) | 629 (96.3%) | 0.4049 |
| MAPK Alterations Absent | 2 (2.2%) | 34 (3.7%) |  | 0 (0.0%) | 24 (3.7%) |  |

Comparable results were observed in the NHW cohort (Table 3b). MAPK alterations were present in approximately 96% of tumors across all NHW subgroups, including early- and late-onset disease and regardless of FOLFOX treatment status. No significant differences in pathway alteration prevalence were detected between treated and untreated patients within either age category, suggesting that MAPK pathway engagement is a common feature of NHW CRC tumors across the disease spectrum.

#### 3.3.2 Ancestry-based comparisons within age and treatment strata

Direct comparisons between H/L and NHW patients further underscored the consistency of MAPK pathway alteration prevalence across populations. Among early-onset CRC patients treated with FOLFOX, MAPK alterations were observed at nearly identical frequencies in H/L and NHW tumors (95.9% vs. 96.3%, respectively; Table 3c). Similarly, in early-onset patients not treated with FOLFOX, pathway alterations were detected in all of the H/L tumors and 96.4% of NHW tumors, without a statistically significant difference.

Ancestry-based comparisons in late-onset CRC followed the same trend (Table 3d). MAPK alterations were present in 97.8% of FOLFOX-treated H/L tumors and 96.3% of treated NHW tumors, while non-FOLFOX-treated late-onset tumors showed alteration frequencies of 100% in H/L patients and 96.3% in NHW patients. Across all comparisons, differences in MAPK pathway alteration prevalence by ancestry did not reach statistical significance.

#### 3.3.3 Interpretation and integration with gene-level findings

Taken together, these results demonstrate that MAPK pathway alterations are nearly universal in colorectal cancer, irrespective of age at onset, ancestry, or FOLFOX exposure. While pathway-level alteration prevalence does not discriminate between clinical subgroups, this apparent uniformity contrasts with the substantial gene-level heterogeneity identified earlier, where specific MAPK components (e.g., NF1, PDGFRB, MAPK3, RPS6KA4, JUN) showed strong age-, ancestry-, and treatment-dependent enrichment. These findings suggest that, although MAPK signaling is broadly implicated across CRC, the specific molecular mechanisms driving pathway dysregulation differ by clinical context, reinforcing the importance of gene-level and subgroup-aware analyses in precision oncology.

### 3.4 Gene-Level Landscape of MAPK Pathway Alterations Across Age, Ancestry, and FOLFOX Exposure

Gene-level interrogation of the MAPK pathway (Tables S1-S12) showed that, despite near-universal pathway-level alteration positivity across strata, most individual genes occurred at low-to-moderate frequencies, with a limited set exhibiting statistically significant enrichment by age, ancestry, and/or treatment status.

Within early-onset H/L tumors (Table S1), most MAPK genes were infrequently mutated and did not differ by FOLFOX exposure. However, three loci showed treatment-associated enrichment in the non-FOLFOX subgroup: FGFR2 (7.7% in non-FOLFOX vs. 0% in FOLFOX; p = 0.02793), NF1 (19.2% vs. 4.1%; p = 0.01437), and RPS6KA4 (9.6% vs. 0%; p = 0.01108). Core drivers remained common in both groups, including KRAS (41.1% FOLFOX vs. 34.6% non-FOLFOX) and TP53 (∼78-81%). In late-onset H/L tumors (Table S2), treatment-associated differences were generally absent, with the exception of increased NTRK2 (6.0% vs. 0%; p = 0.04286) and PDGFRB (6.0% vs. 0%; p = 0.04286) in the non-FOLFOX group; BRAF was notably higher in late-onset H/L overall (16.0-16.5%) than in early-onset H/L (4.1-7.7%), independent of therapy.

Among early-onset NHW tumors (Table S3), the MAPK gene spectrum was similarly stable across FOLFOX strata, but significant differences emerged for select genes: AKT3 mutations were more frequent in non-FOLFOX cases (3.0% vs. 0.8%; p = 0.04069), FGF4 was detected only in non-FOLFOX tumors (1.7% vs. 0%; p = 0.01734), JUN was enriched in FOLFOX-treated tumors (3.2% vs. 0.7%; p = 0.02726), and RRAS2 occurred exclusively in non-FOLFOX tumors (1.3% vs. 0%; p = 0.03916). In late-onset NHW tumors (Table S4), multiple genes differed by treatment status, with higher prevalence in non-FOLFOX cases for CRKL (1.2% vs. 0.1%; p = 0.004928), DUSP4 (0.9% vs. 0.1%; p = 0.02292), JUN (3.2% vs. 1.4%; p = 0.02486), MAPK1 (0.9% vs. 0.1%; p = 0.02292), RAF1 (2.9% vs. 0.9%; p = 0.004111), RPS6KA4 (5.2% vs. 1.8%; p = 0.0003744), RRAS (1.2% vs. 0.2%; p = 0.02024), and SOS1 (2.3% vs. 0.9%; p = 0.03501). Additionally, TP53 was modestly higher in FOLFOX-treated late-onset NHW tumors (73.4% vs. 68.1%; p = 0.02559), and TGFBR2 was higher in non-FOLFOX cases (7.0% vs. 4.1%; p = 0.01578).

Age-stratified analyses reinforced age-associated biology within treatment strata. Among FOLFOX-treated H/L cases (Table S5), BRAF was significantly higher in late-onset compared with early-onset tumors (16.5% vs. 4.1%; p = 0.01229), and PDGFRB was observed only in early-onset tumors (5.5% vs. 0%; p = 0.03747). Among non-FOLFOX H/L cases (Table S6), NF1 remained enriched in early-onset disease (19.2% vs. 4.0%; p = 0.02834), consistent with a stronger NF1 signal in younger untreated H/L tumors. In the NHW cohort, age comparisons in both treated (Table S7) and untreated (Table S8) strata also supported increased BRAF burden with later onset: BRAF was higher in late-onset versus early-onset non-FOLFOX NHW tumors (14.2% vs. 7.9%; p = 0.007983), whereas gene-level differences were otherwise limited.

Direct ancestry comparisons within EOCRC showed that most MAPK genes were broadly similar between H/L and NHW within each treatment group (Tables S9-S11). Nonetheless, ancestry-specific signals were evident in non-FOLFOX EOCRC, where H/L tumors showed higher frequencies of MAPK3 (5.8% vs. 1.0%; p = 0.04335), NF1 (19.2% vs. 6.0%; p = 0.002728), and RPS6KA4 (9.6% vs. 2.6%; p = 0.03866) (Table S10). In contrast, in FOLFOX-treated EOCRC, PDGFRB was higher in H/L than NHW tumors (5.5% vs. 0.8%; p = 0.01547) (Table S9). In late-onset non-FOLFOX comparisons (Table S12), NTRK2 was more frequent in H/L than NHW (6.0% vs. 1.4%; p = 0.0472), and RRAS2 was enriched in H/L (4.0% vs. 0.5%; p = 0.04318), while most other loci did not differ by ancestry.

These stratified gene-level results indicate that MAPK pathway disruption in CRC is ubiquitous at the pathway level, but specific nodes, particularly NF1, FGFR2, RPS6KA4, BRAF, and select RTK/adaptor genes (PDGFRB, NTRK2, RAF1, SOS1, CRKL), show context-dependent enrichment by age, ancestry, and chemotherapy exposure.

### 3.5 MAPK Pathway Mutation Profile

#### 3.5.1 Early-Onset Hispanic/Latino Colorectal Cancer

Figure 1a presents an integrated overview of somatic alterations affecting MAPK pathway genes in EOCRC H/L patients (EOCRC; n = 122), combining mutation type, TMB, and FOLFOX treatment annotation. Unlike signaling pathways that are altered only in a subset of tumors, MAPK pathway involvement was universal in this cohort, with every tumor (122/122; 100%) harboring at least one alteration within the curated MAPK gene set.

**Figure 1.**
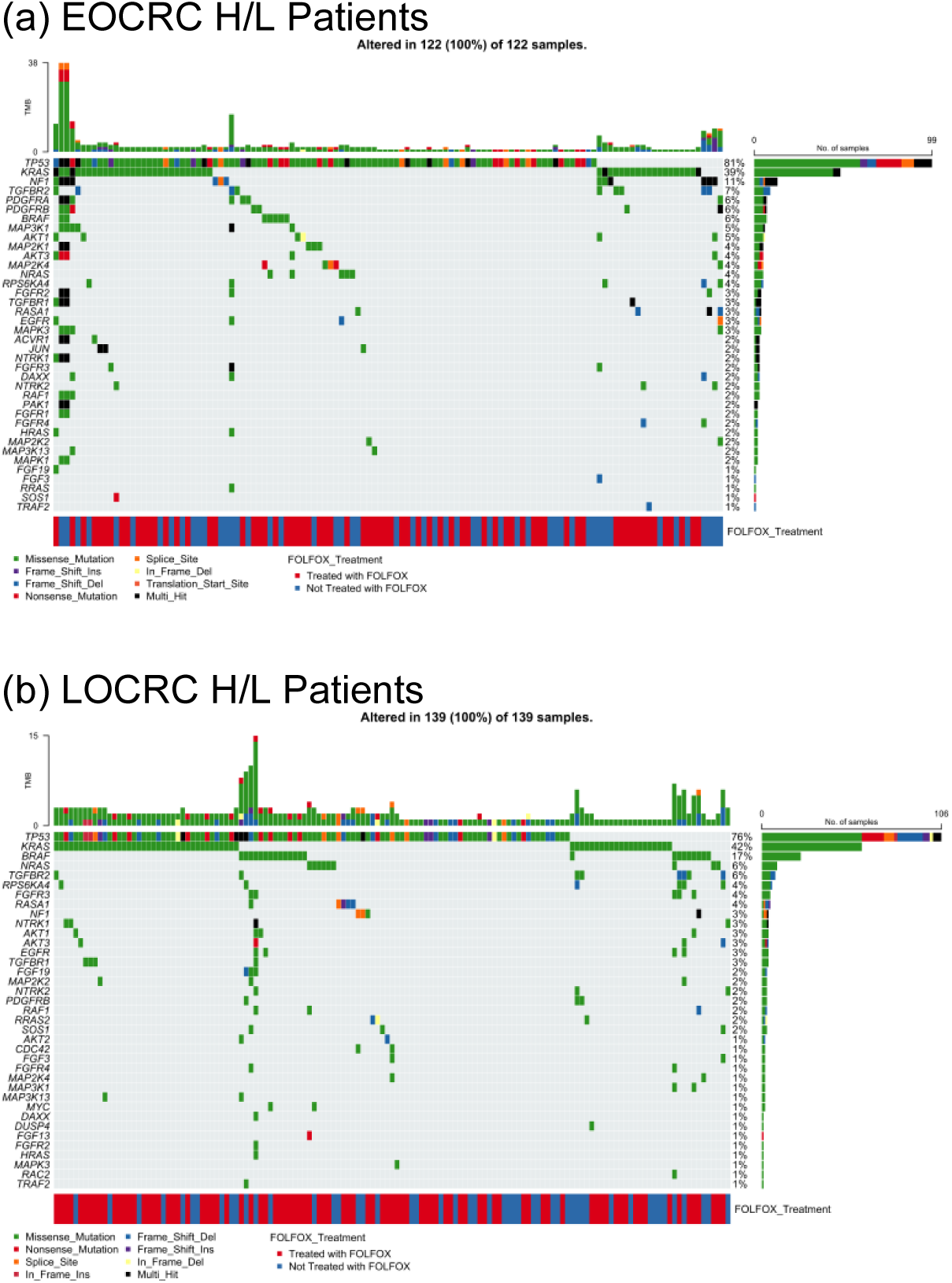
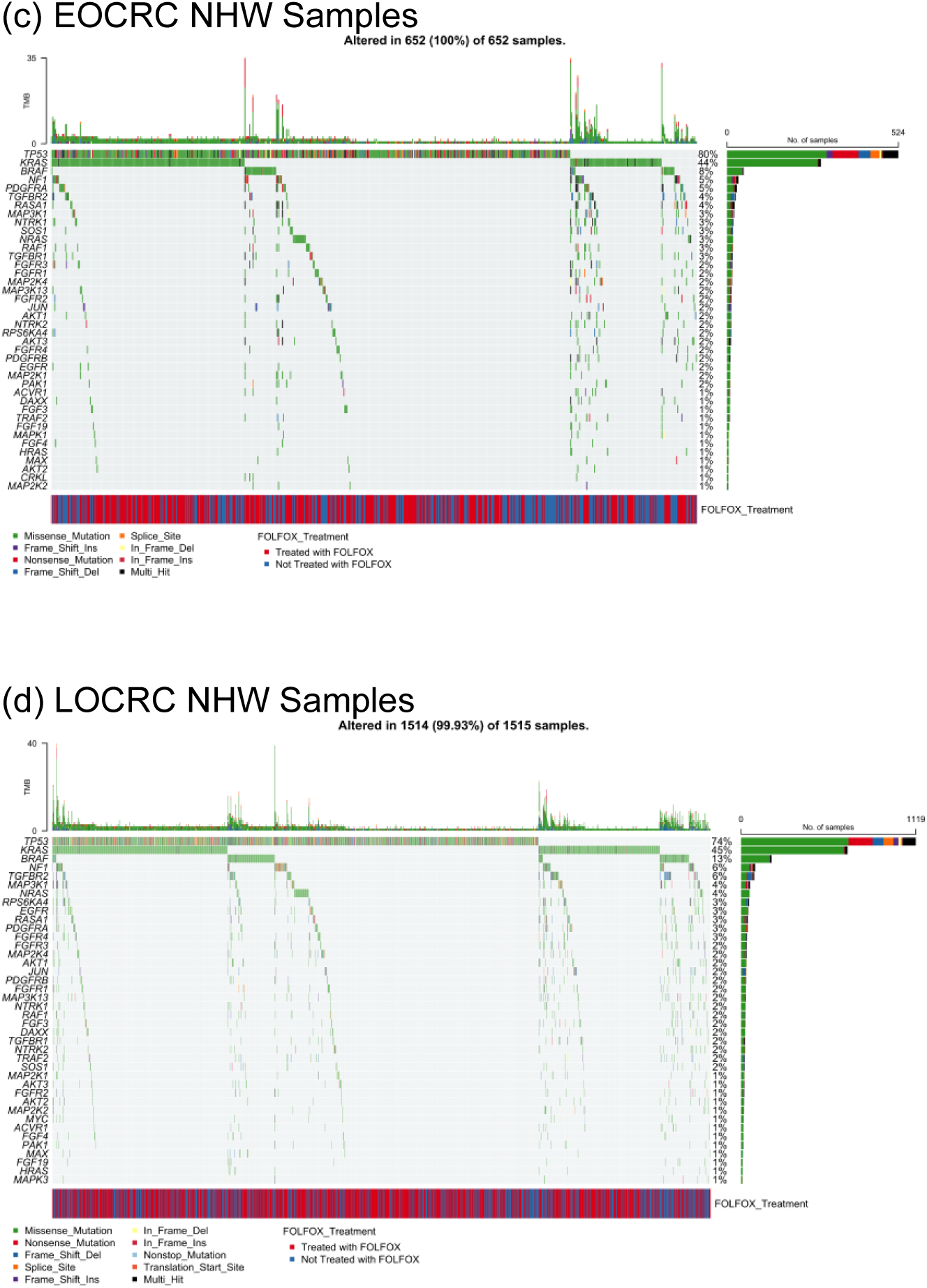
MAPK pathway mutational landscapes across colorectal cancer subgroups defined by age of onset and ancestry. This figure displays oncoplot visualizations summarizing somatic mutations in genes involved in MAPK signaling across colorectal cancer (CRC) cohorts stratified by diagnostic age and ancestral background. Separate panels illustrate mutation patterns for early-onset and late-onset CRC within Hispanic/Latino (H/L) and non-Hispanic White (NHW) populations. Each oncoplot integrates variant class, TMB, and FOLFOX chemotherapy exposure, enabling direct comparison of MAPK pathway disruption across clinically and demographically distinct groups. Panels correspond to: (a) early-onset H/L CRC, (b) late-onset H/L CRC, (c) early-onset NHW CRC, and (d) late-onset NHW CRC.

The mutational landscape was anchored by well-established colorectal cancer drivers. TP53 was altered most frequently (81%), followed by KRAS (39%). Additional recurrent events involved regulators and modulators of MAPK signaling, including NF1 and TGFBR2 (each 11%). A broader distribution of less frequent alterations spanned both upstream receptor tyrosine kinases and downstream signaling intermediates, such as PDGFRA (∼7%) and a group of genes altered in approximately 5-6% of tumors (PDGFRB, BRAF, MAP3K1, AKT1, MAP2K1). Numerous other pathway components, including NRAS, MAP2K4, RPS6KA4, FGFR2, TGFBR1, RASA1, EGFR, and MAPK3, were affected at lower frequencies (typically 2-4%), reflecting substantial inter-tumor heterogeneity within a broadly perturbed pathway.

At the variant level, missense substitutions constituted the predominant mutation class, with fewer loss-of-function events such as frame-shift insertions or deletions, nonsense mutations, and sporadic splice-site variants. Multi-hit alterations were rare and restricted to individual cases. TMB values spanned a wide range, including a subset of tumors with elevated mutational burden; however, these hypermutated samples did not show preferential association with any specific MAPK gene, indicating that increased TMB reflects global genomic instability rather than selective MAPK pathway hypermutation.

Assessment of chemotherapy exposure revealed no overt segregation of MAPK alterations by FOLFOX treatment status. Treated and untreated tumors were evenly distributed across the mutational spectrum, suggesting that, in EOCRC H/L patients, MAPK pathway disruption is widespread, diverse, and not visually driven by chemotherapy exposure at the pathway-wide level.

#### 3.5.2 Late-Onset Hispanic/Latino CRC

Figure 1b depicts the MAPK pathway mutational profile of LOCRC H/L tumors (n = 139), integrating somatic variant classes, TMB, and FOLFOX treatment status. Consistent with findings in early-onset disease, MAPK pathway alterations were observed in all tumors (139/139; 100%), underscoring the pervasive involvement of MAPK signaling in H/L CRC across age groups.

The mutational spectrum in late-onset H/L CRC was dominated by canonical CRC drivers and core MAPK-associated genes. TP53 was the most frequently altered gene (76%), followed by KRAS (42%) and BRAF (17%), reflecting a higher prevalence of BRAF alterations compared with early-onset H/L tumors. Additional recurrent alterations were observed in NRAS (∼7%) and TGFBR2 (∼6%), with a broader distribution of lower-frequency mutations across multiple pathway regulators and effectors, including RPS6KA4, FGFR2, RASA1, NF1, NTRK1, AKT3, EGFR, TGFBR1, MAP2K2, and NTRK2 (generally ∼2-4% each). This pattern illustrates a more diversified MAPK mutational landscape in late-onset disease, characterized by contributions from both upstream receptor tyrosine kinases and downstream signaling intermediates.

As in early-onset tumors, missense mutations (green) constituted the majority of events, with fewer truncating alterations such as frame-shift insertions or deletions (blue shades) and nonsense mutations (red). Multi-hit events were uncommon and did not cluster within specific genes. TMB values were generally modest across the cohort, with a subset of tumors exhibiting elevated mutational burden; however, these higher-TMB cases were not preferentially associated with any single MAPK gene, suggesting that increased TMB reflects global genomic instability rather than focal MAPK hypermutation.

The FOLFOX treatment annotation (blue = treated; red = not treated) was distributed across the cohort without an apparent segregation of MAPK alterations by chemotherapy exposure. Both treated and untreated tumors displayed comparable patterns of pathway involvement, indicating that, at the pathway-wide level, MAPK alterations in late-onset H/L CRC are age-associated but not visually driven by FOLFOX status.

The LOCRC H/L MAPK mutational landscape reveals a broader and more BRAF-enriched profile compared with early-onset disease, reinforcing age-related differences in MAPK pathway architecture while highlighting the ubiquitous role of MAPK signaling in colorectal tumorigenesis within H/L populations.

#### 3.5.3 Early-Onset Non-Hispanic White CRC

Figure 1c illustrates the MAPK pathway mutational landscape in EOCRC NHW tumors (n = 652), integrating gene-level alterations, TMB, and FOLFOX treatment status. In line with pathway-level analyses, MAPK pathway alterations were present in all tumors (652/652; 100%), confirming widespread disruption of MAPK signaling in early-onset disease within the NHW population.

The mutational profile was dominated by well-established CRC driver genes. TP53 alterations were observed in approximately 80% of tumors, followed by KRAS (48%) and BRAF (8%), reflecting a lower BRAF frequency compared with late-onset NHW disease and late-onset H/L tumors. Additional recurrent alterations were distributed across multiple MAPK-associated genes, including NF1 (∼5%), PDGFRA (∼5%), RASGRF1, RASA1, MAP3K1, NTRK1, NRAS, SOS1, and TGFBR2 (each ∼3-4%). A broad tail of lower-frequency mutations (<3%) affected both upstream receptor tyrosine kinases (e.g., EGFR, FGFR1/2) and downstream signaling components (e.g., MAP2K1, MAP3K3, AKT3, RPS6KA4), underscoring substantial inter-tumor heterogeneity.

Across the cohort, missense mutations (green) represented the predominant alteration type, with fewer nonsense mutations, frame-shift insertions/deletions, and occasional splice-site events. Multi-hit alterations were infrequent and did not cluster within specific genes. TMB values spanned a wide range, including a subset of hypermutated tumors characterized by prominent TMB peaks; however, these high-TMB cases did not preferentially harbor mutations in any single MAPK gene, suggesting that elevated mutational load reflects generalized genomic instability rather than focal MAPK pathway hypermutation.

The FOLFOX treatment annotation track (blue = treated; red = not treated) was evenly interspersed across samples, with no clear segregation of MAPK alteration patterns by chemotherapy exposure at the pathway-wide level. Both treated and untreated tumors exhibited similar distributions of driver and non-driver MAPK alterations, indicating that early-onset NHW MAPK genomic architecture is largely independent of FOLFOX status when assessed globally.

Early-onset NHW CRC demonstrates a KRAS- and TP53-dominated MAPK mutational profile, with lower BRAF involvement and extensive low-frequency alterations across the pathway. When considered alongside ancestry-comparative analyses, these findings highlight both shared MAPK pathway dependence across populations and subtle differences in the relative contribution of specific MAPK nodes to EOCRC tumorigenesis.

#### 3.5.4 Late-Onset Non-Hispanic White CRC

Figure 1d depicts the MAPK pathway mutational architecture of LOCRC NHW tumors (n = 1,515), integrating somatic variant types, TMB, and FOLFOX treatment annotation. Consistent with pathway-level analyses, MAPK pathway alterations were detected in nearly all tumors (1,514/1,515; 99.9%), reinforcing the central role of MAPK signaling in late-onset CRC within the NHW population.

The mutational spectrum was dominated by canonical CRC drivers. TP53 alterations were the most prevalent (∼74%), followed by KRAS (∼43%) and BRAF (∼15%), reflecting a marked enrichment of BRAF mutations relative to early-onset NHW disease. Additional recurrent MAPK-associated alterations were observed in NF1 (∼6%), TGFBR2 (∼5%), MAP3K1 (∼4%), NRAS (∼4%), and RPS6KA4 (∼3%), with a wide distribution of lower-frequency events affecting receptor tyrosine kinases and signaling intermediates such as PDGFRA, FGFR4, MAP2K1, MAPK3, AKT1, JUN, PDGFRB, EGFR, MAP3K3, and NTRK1 (generally ∼1-3%). This pattern indicates a broad and heterogeneous disruption of MAPK signaling nodes in late-onset NHW tumors.

As observed in other cohorts, missense mutations (green) accounted for the majority of alterations, accompanied by less frequent truncating events, including frame-shift insertions/deletions, nonsense mutations, and occasional splice-site variants. Multi-hit alterations were rare and did not cluster within specific MAPK genes. TMB values ranged from low to moderately elevated, with a subset of hypermutated tumors visible as prominent TMB peaks. However, these high-TMB samples were not preferentially associated with mutations in any single MAPK pathway component, suggesting that increased mutational load reflects global genomic instability rather than selective MAPK hypermutation.

The FOLFOX treatment annotation track (blue = treated; red = not treated) was evenly interspersed across the cohort, with no clear segregation of MAPK alteration patterns by chemotherapy exposure at the pathway-wide level. Both treated and untreated tumors demonstrated comparable distributions of high-frequency drivers and lower-frequency MAPK alterations.

LOCRC NHW exhibits a TP53-KRAS-BRAF-enriched MAPK mutational profile with extensive low-frequency pathway perturbations. Compared with early-onset NHW disease, late-onset tumors show a higher contribution of BRAF and NF1 alterations, consistent with age-associated shifts in MAPK pathway biology, while maintaining broad pathway involvement independent of FOLFOX treatment status.

### 3.6 Survival Associations of MAPK Pathway Alterations

To evaluate the prognostic relevance of MAPK pathway alterations in colorectal cancer, we conducted Kaplan-Meier analyses stratified by age of onset, ancestry, and FOLFOX treatment exposure.

#### 3.6.1 Early-Onset Hispanic/Latino Patients Receiving FOLFOX

In EOCRC H/L patients treated with FOLFOX (Figure 2a), overall survival did not differ significantly according to MAPK pathway mutation status (p = 0.39). Survival trajectories for tumors harboring MAPK alterations and those without detectable pathway mutations followed largely overlapping courses across the duration of follow-up, indicating no clear separation between groups.

**Figure 2.**
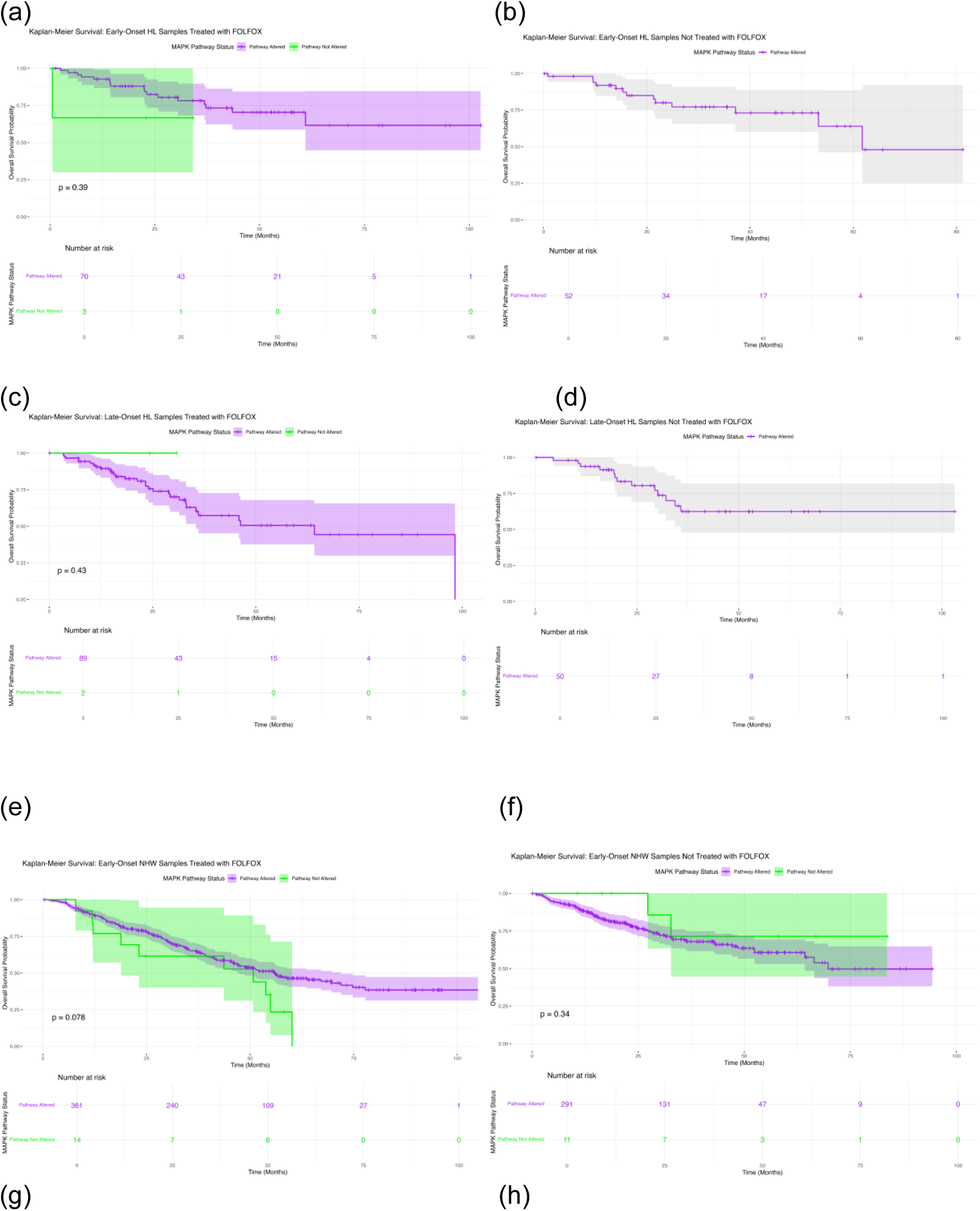
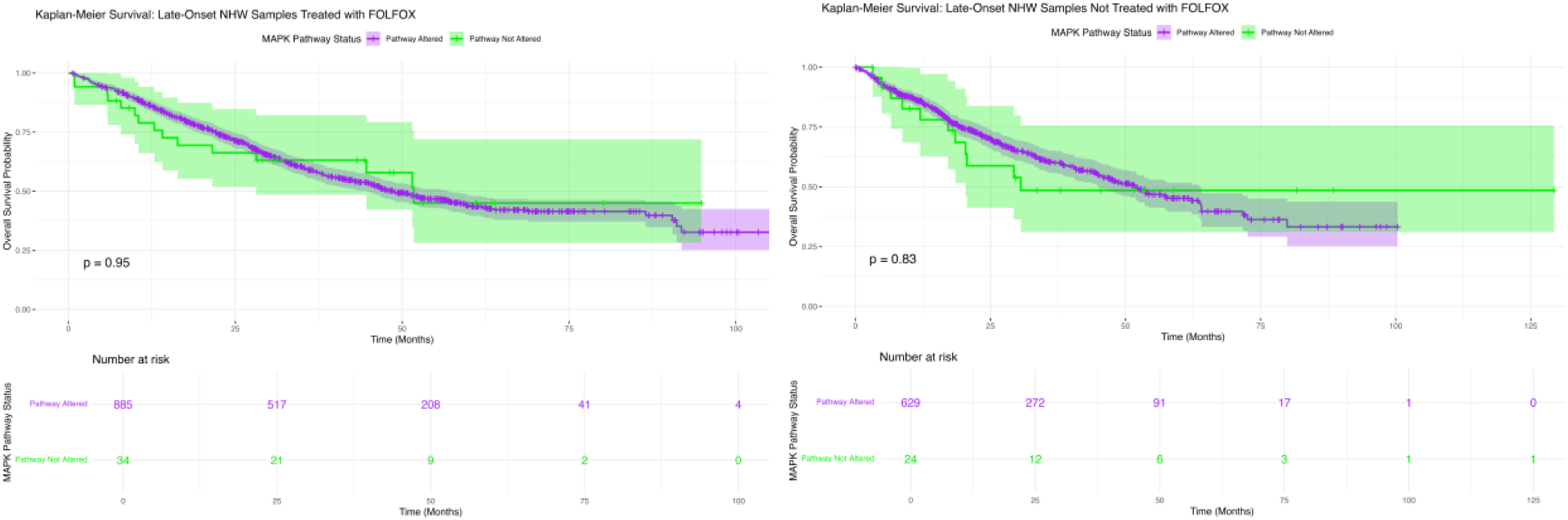
Overall survival by MAPK pathway alteration status across colorectal cancer subgroups defined by age, ancestry, and chemotherapy exposure. This figure presents Kaplan-Meier survival analyses comparing overall survival between colorectal cancer (CRC) patients with and without MAPK pathway alterations, stratified by age at diagnosis, ancestral background, and FOLFOX treatment status. Survival curves are shown for the following subgroups: (a) early-onset Hispanic/Latino patients receiving FOLFOX; (b) early-onset Hispanic/Latino patients not exposed to FOLFOX; (c) late-onset Hispanic/Latino patients treated with FOLFOX; (d) late-onset Hispanic/Latino patients not treated with FOLFOX; (e) early-onset non-Hispanic White patients receiving FOLFOX; (f) early-onset non-Hispanic White patients not receiving FOLFOX; (g) late-onset non-Hispanic White patients treated with FOLFOX; and (h) late-onset non-Hispanic White patients not treated with FOLFOX. Within each panel, outcomes for tumors harboring MAPK pathway mutations are contrasted with those lacking detectable alterations. Shaded regions denote 95% confidence intervals, and tabulated counts beneath each plot indicate the number of individuals at risk over time, enabling assessment of survival dynamics within each clinically and demographically defined subgroup.

Patients with MAPK-altered tumors demonstrated broader confidence intervals, reflecting the smaller size of the non-altered comparator group and increased uncertainty at later time points. Despite this variability, long-term survival probabilities remained comparable between cohorts, with no consistent trend toward improved or worsened outcomes associated with MAPK pathway disruption. The non-altered group showed a modest decline in survival beyond approximately 25-30 months, though overall survival remained high.

These results suggest that, among early-onset H/L patients receiving FOLFOX, MAPK pathway alteration status does not confer a statistically significant prognostic effect on overall survival. These findings should be interpreted in the context of limited sample sizes in the non-altered subgroup but indicate that, within this treatment-defined population, MAPK pathway involvement alone is insufficient to stratify survival risk.

#### 3.6.2 Early-Onset Hispanic/Latino Patients Without FOLFOX Exposure

Among EOCRC H/L patients who did not undergo FOLFOX treatment (Figure 2b), overall survival patterns were predominantly uniform across MAPK pathway subgroups. All tumors within this cohort harbored MAPK pathway alterations, preventing pathway-based stratification and precluding a direct comparison between altered and non-altered outcomes.

The survival curve for the MAPK-altered cohort showed a gradual decline over time, with the majority of patients maintaining high survival probability during the initial follow-up period. Confidence intervals widened notably at later time points, reflecting attrition in the number of individuals remaining under observation. While survival probability decreased after approximately 50-60 months, no distinct inflection point or abrupt change in trajectory was observed.

Although these findings reinforce the pervasive presence of MAPK alterations in early-onset H/L CRC, they also indicate that, within this untreated subgroup, pathway alteration status does not meaningfully differentiate survival risk. The absence of non-altered comparators, combined with broad confidence intervals and declining sample size over time, limits definitive interpretation but supports the conclusion that MAPK disruption, at a pathway-wide level, is not a primary determinant of survival outcomes in early-onset H/L tumors lacking FOLFOX exposure.

#### 3.6.3 Late-Onset Hispanic/Latino Patients Receiving FOLFOX

Among LOCRC H/L patients treated with FOLFOX (Figure 2c), survival outcomes appeared broadly similar regardless of MAPK pathway alteration status. No statistically significant difference in overall survival was observed between MAPK-altered and non-altered groups (p = 0.43), indicating an absence of clear prognostic discrimination within this subgroup.

The survival curve for MAPK-altered tumors demonstrated a steady decline across the follow-up period, while the curve for non-altered tumors, represented by a small comparator subset, remained relatively stable early on before converging toward the altered trajectory at later time points. Wide confidence intervals, particularly for the non-altered samples, highlighted the small number of individuals without MAPK pathway alterations and contributed to uncertainty in the survival estimates.

Despite visible variability in curve shape, the overlapping confidence bands and similar long-term survival probabilities suggest that MAPK mutation status does not meaningfully influence prognosis among late-onset H/L patients undergoing FOLFOX therapy. As in other subgroups, sample size imbalance limits the precision of comparative interpretation; however, the overall pattern supports the conclusion that MAPK pathway disruption, at the pathway level, is not a dominant survival determinant in this clinical context.

#### 3.6.4 Late-Onset Hispanic/Latino Patients Without FOLFOX Treatment

In the subgroup of LOCRC H/L patients who did not receive FOLFOX (Figure 2d), overall survival outcomes were largely uniform, with no apparent variation attributable to MAPK pathway mutation status. Because all tumors in this subgroup carried MAPK pathway alterations, a non-altered comparator group was not present, preventing pathway-based stratification within the cohort.

The survival curve displayed a gradual early decline followed by stabilization across later follow-up intervals. Confidence intervals widened over time, reflecting fewer individuals remaining under observation at later survival points, but overall survival probabilities stayed relatively consistent throughout the duration assessed.

These findings highlight the widespread presence of MAPK alterations in LOCRC H/L patients and indicate that, in the absence of FOLFOX chemotherapy, MAPK pathway involvement does not visibly distinguish survival trajectories within this subgroup. However, the absence of a non-altered comparison cohort limits the ability to draw conclusions regarding the prognostic value of these alterations in untreated late-onset H/L tumors.

#### 3.6.5 Early-Onset Non-Hispanic White Patients Treated With FOLFOX

Among EOCRC NHW patients receiving FOLFOX chemotherapy (Figure 2e), overall survival patterns showed modest divergence based on MAPK pathway status. Although the comparison did not reach statistical significance (p = 0.078), visual inspection of the Kaplan-Meier curves revealed trends worth noting.

Patients with MAPK-altered tumors demonstrated a gradual but steady decline in survival over the full follow-up period, maintaining intermediate survival probabilities throughout. In contrast, the non-altered group, although considerably smaller in size, displayed a steeper drop in survival within the first 40 months, followed by stabilization at lower survival levels. By later follow-up points, confidence intervals widened substantially for both curves, particularly for the non-altered subgroup, reflecting diminishing sample size and reduced precision of estimates.

Despite the absence of a statistically significant result, the widening separation between curves over time suggests a potential biological distinction between altered and non-altered MAPK groups in this setting. However, given the small number of MAPK-non-altered cases and wide variability at later time points, these observations should be interpreted cautiously. Taken together, the data indicate that while MAPK mutation status does not definitively stratify survival outcomes in FOLFOX-treated EOCRC NHW patients, pathway differences may contribute to subtle survival variation that warrants further investigation in larger cohorts.

#### 3.6.6 Early-Onset NHW Patients Not Treated With FOLFOX

For EOCRC NHW patients who did not receive FOLFOX chemotherapy (Figure 2f), overall survival outcomes did not show a statistically meaningful association with MAPK pathway alteration status (p = 0.34). Both MAPK-altered and non-altered groups began with similarly high survival probabilities and followed closely aligned trajectories across the observation period.

A slight divergence appeared midway through follow-up, during which the non-altered subgroup showed somewhat higher survival probabilities than the altered cohort. However, this separation remained modest and transient, and confidence intervals surrounding both curves were wide and largely overlapping, particularly for the smaller non-altered group, underscoring notable variability and limited statistical power.

By late follow-up, survival estimates converged again between groups, further supporting the absence of a strong survival distinction driven by MAPK pathway status in this clinical context. Overall, these findings suggest that, among EOCRC NHW patients who did not receive FOLFOX, MAPK alterations do not exert a clear or measurable influence on overall survival, though small subgroup sizes warrant cautious interpretation.

#### 3.6.7 Late-Onset NHW Patients Treated With FOLFOX

Among LOCRC NHW patients treated with FOLFOX chemotherapy (Figure 2g), overall survival showed no meaningful distinction based on MAPK pathway mutation status. Survival curves for MAPK-altered and non-altered tumors were nearly superimposed throughout the observation window, with both groups demonstrating a steady decline over time and comparable long-term outcomes (p = 0.95).

Confidence intervals for the two trajectories overlapped extensively, reinforcing the lack of separation between groups. Although minor variations were visible in early and mid-follow-up, these fluctuations did not persist and did not suggest any consistent advantage or disadvantage tied to pathway alteration.

These findings indicate that, within this large LOCRC NHW cohort receiving FOLFOX, MAPK pathway mutation status does not appear to influence overall survival, highlighting a largely uniform response pattern to chemotherapy regardless of underlying MAPK alteration profile.

#### 3.6.8 Late-Onset NHW Patients Not Treated With FOLFOX

In LOCRC NHW patients who did not receive FOLFOX chemotherapy (Figure 2h), overall survival outcomes were broadly similar between tumors with and without MAPK pathway alterations (p = 0.83). Both trajectories began with near-complete survival and exhibited a steady decline over time, with no durable separation emerging between altered and non-altered groups.

Although the MAPK-altered cohort showed a slightly steeper drop in survival during mid-follow-up, confidence intervals for both groups overlapped extensively across the entire observation period, indicating substantial uncertainty and no reliable evidence of prognostic divergence. The non-altered subgroup, though smaller in size, demonstrated wide confidence shading, reflecting reduced precision in later time points but still aligning closely with the altered group’s survival estimate.

These results suggest that MAPK pathway mutation status does not impart a measurable survival difference in untreated LOCRC NHW patients, reinforcing the broader observation that MAPK alterations, while frequent, may not independently predict outcome in the absence of FOLFOX exposure.

### 3.7 Conversational Artificial Intelligence

#### 3.7.1 AI-enabled exploratory cohort stratification and survival assessment

To generate preliminary clinical insight prior to pathway-specific modeling, we used the conversational AI framework to rapidly construct ancestry-matched survival cohorts and perform exploratory outcome comparisons. This approach allowed real-time interrogation of integrated clinical-genomic CRC data while preserving strict user-defined eligibility rules.

Figure S1 provides one such example, focusing on early-onset CRC patients treated with FOLFOX chemotherapy. The AI platform automatically identified: 73 early-onset H/L FOLFOX-treated cases (panel a), representing 2.9% of available samples in this subgroup, and 375 early-onset NHW FOLFOX-treated controls (panel b), accounting for 14.9% of records within the corresponding ancestry group. Kaplan-Meier analysis performed through the platform revealed a clear separation in overall survival trajectories between the two ancestry groups (panel c), with significantly improved outcomes observed in the H/L cohort (log-rank p = 0.0310). The widening confidence bands over longer follow-up reflect decreasing patient numbers at risk but do not obscure the durable survival advantage.

This exploratory result demonstrates how real-time AI-directed cohort refinement can uncover clinically meaningful ancestry-based differences within treatment-defined strata. These findings provided the rationale for subsequent MAPK-focused analyses and highlight the platform’s utility in rapidly identifying outcome patterns that warrant deeper mechanistic and statistical investigation.

#### 3.7.2 AI-assisted comparison of MSI stability in early-onset Hispanic/Latino CRC stratified by treatment exposure

To further evaluate molecular differences within ancestry-defined subgroups, we applied the conversational AI framework to examine whether microsatellite stability patterns varied by chemotherapy exposure among early-onset H/L colorectal cancer patients. Using natural language-directed cohort construction, the system identified two treatment-defined populations: those who received FOLFOX (n = 73) and those who did not (n = 52).

Panels (a) and (b) in Figure S2 illustrate the proportions of cases that met inclusion criteria relative to all available samples, confirming that both subgroups represented a small and comparable fraction of the dataset.

A contingency comparison was then conducted to assess whether MSI-stable tumor status differed between treated and untreated groups. As shown in panel (c), MSI-stable tumors were distributed similarly across cohorts, and Fisher’s exact testing demonstrated no statistically significant association between MSI category and treatment status (p = 0.785). Odds ratio outputs likewise indicated no directional trend favoring either subgroup.

These findings suggest that, within early-onset H/L CRC, MSI stability is not influenced by FOLFOX exposure, reinforcing the interpretation that treatment alone does not drive detectable variation in MSI-related tumor biology in this population. This analysis also highlights the practical value of the AI-guided workflow in rapidly generating cohort-specific molecular comparisons and validating null relationships within highly granular clinical strata.

#### 3.7.3 AI-assisted exploratory comparison of AKT3 mutation frequency in early-onset NHW CRC by treatment exposure

As part of our exploratory computational workflow, we applied the conversational AI framework to interrogate whether AKT3 mutations varied according to chemotherapy exposure within early-onset NHW colorectal cancer patients. Using AI-directed cohort filtering, the system automatically identified two treatment-defined groups: individuals who received FOLFOX (n = 375) and those who did not (n = 302) (Figure S3).

Panels (a) and (b) illustrate the proportion of samples in each cohort that met the AKT3 mutation criterion, confirming that only a small minority of tumors were classified as AKT3-altered. Notably, mutation prevalence was slightly lower in the FOLFOX-treated cohort (14.9%) compared with untreated patients (12.0%), although both groups showed similar overall distributions.

A stacked bar chart (panel c) summarizes results from an odds ratio comparison using Fisher’s exact test. Statistical evaluation revealed no significant association between treatment exposure and AKT3 alteration status (p = 0.065), indicating that AKT3 mutation patterns remain largely stable regardless of chemotherapy history. While the numerical signal suggested marginally reduced alteration rates in treated cases, confidence interval width and p-value trends supported the interpretation that differences were not meaningful.

Taken together, this AI-mediated analysis demonstrates that AKT3 mutation frequency does not appear to differ substantially between treated and untreated early-onset NHW CRC patients, and further highlights the utility of rapid, query-driven cohort generation for validating molecular equivalence or divergence across clinically relevant patient subgroups.

#### 3.7.4 AI-guided exploratory comparison of PDGFRB alterations in early-onset FOLFOX-treated patients across ancestry groups

As part of the exploratory AI interrogation, we evaluated whether PDGFRB mutation prevalence differed between early-onset H/L and NHW colorectal cancer patients treated with FOLFOX (Figure S4). Using conversational filtering, the AI platform automatically assembled a case cohort of H/L patients (n = 73) and a control cohort of NHW patients (n = 375) meeting identical clinical conditions, ensuring that treatment status and age of onset were matched across ancestry groups. Visual inspection of the pie charts in panels (a) and (b) confirmed accurate extraction of the relevant comparison subsets, with only a small fraction of individuals in either cohort exhibiting PDGFRB alterations.

Subsequent odds ratio analysis is summarized in panel (c), demonstrating that PDGFRB mutations were uncommon in both ancestry-defined cohorts. Mutation frequency was numerically lower among H/L patients, but Fisher’s exact testing indicated no statistically significant difference between groups (p = 0.502), and the odds ratio suggested a minimal effect size consistent with rare-event variability.

Taken together, these exploratory results indicate that PDGFRB mutation patterns appear largely equivalent across ancestry in early-onset FOLFOX-treated CRC, reinforcing that this gene is unlikely to contribute meaningfully to ancestry-specific molecular divergence within this clinical setting. Importantly, this analysis illustrates the capability of the conversational AI system to rapidly generate ancestry-matched cohorts and validate mutation frequency comparisons, even in the context of sparsely mutated genes.

#### 3.7.5 AI-driven identification of mutation-level disparities within MAPK-altered early-onset tumors across ancestry groups

To further characterize molecular variation linked to MAPK pathway dysregulation, we used the conversational AI system to compare mutation attributes between MAPK-altered early-onset H/L and NHW colorectal tumors (Figure S5). The platform autonomously generated ancestry-stratified cohorts based on integrated clinical-genomic criteria, resulting in 122 MAPK-altered H/L tumors and 652 MAPK-altered NHW tumors. Panels (a) and (b) illustrate cohort distribution patterns relative to the full data space, demonstrating a markedly larger representation of MAPK-altered cases among NHW patients.

The AI then performed exploratory, attribute-level prioritization to identify statistically significant differences between groups (panel c). The mutation profile comparison revealed broad ancestry-associated heterogeneity, with several canonical colorectal cancer drivers enriched at different rates across the two populations. Highly significant differences (p < 0.001) were observed for TP53, APC, KRAS, TCF7L2, and PIK3CA, followed by additional signals involving SMAD4, BRAF, PIK3R1, and NF1. Many of these genes intersect biologically with MAPK signaling or represent upstream regulators of pathway activation, underscoring their mechanistic relevance.

These exploratory findings suggest that MAPK-altered early-onset CRC is not molecularly uniform across ancestry groups, but instead reflects distinct co-mutation landscapes. The rapid attribute extraction made possible by conversational AI enabled the identification of multiple genomic features with potential relevance to ancestry-specific CRC biology. These results informed subsequent focused analyses and highlight the utility of AI-assisted interrogation for uncovering context-dependent molecular variation within MAPK-driven disease.

## 4. Discussion

This study represents, to our knowledge, one of the first conversational AI-enabled, pathway-focused analyses of the MAPK signaling axis in colorectal cancer systematically stratified by age at diagnosis, ancestry, and FOLFOX exposure, with an explicit emphasis on H/L populations. Using the AI-HOPE framework and its pathway-specific module AI-HOPE-MAPK, we harmonized multi-cohort clinical and genomic data from 2,515 CRC cases and interrogated MAPK alterations through natural language-driven analytics combined with conventional biostatistical testing. This integrative approach allowed us to move beyond single-gene biomarkers and to examine how MAPK pathway disruption and co-mutation patterns vary across clinically and demographically relevant contexts.

### Summary of key findings

First, MAPK pathway involvement was nearly universal across CRC, independent of ancestry, age of onset, or FOLFOX treatment. Pathway-level alterations were detected in ∼96-100% of tumors in every subgroup examined, including EOCRC and LOCRC, H/L and NHW patients, and both FOLFOX-treated and non-FOLFOX-treated cohorts. This ubiquity underscores the central role of MAPK signaling in colorectal tumorigenesis but also indicates that simple “MAPK-altered vs. not altered” status is not a useful discriminator for patient stratification.

Second, substantial heterogeneity emerged at the gene level, revealing context-dependent enrichment of specific MAPK components. In EO H/L tumors not exposed to FOLFOX, NF1, FGFR2, and the downstream effector RPS6KA4 were significantly enriched compared with treated EO H/L cases. LO H/L tumors not treated with FOLFOX showed higher frequencies of NTRK2 and PDGFRB mutations than their treated counterparts. Within the NHW population, multiple MAPK-related genes, including AKT3, FGF4, JUN, RRAS2 in EOCRC and CRKL, DUSP4, MAPK1, RAF1, RPS6KA4, RRAS, and SOS1 in LOCRC, displayed higher prevalence in non-FOLFOX-treated tumors, while JUN showed the opposite pattern in some strata. Age-stratified comparisons further highlighted BRAF enrichment in late-onset disease across ancestries and reinforced NF1 enrichment in early-onset tumors, particularly in untreated H/L cases.

Third, ancestry-specific differences were most pronounced in early-onset, chemo-naïve disease. Among EOCRC patients not treated with FOLFOX, H/L tumors showed higher frequencies of MAPK3, NF1, and RPS6KA4 mutations than NHW tumors, whereas among FOLFOX-treated EOCRC cases, PDGFRB alterations were more common in H/L than NHW tumors. These findings indicate that, although MAPK pathway disruption is pervasive in both populations, the specific nodes through which the pathway is perturbed differ by ancestry and treatment context.

Fourth, pathway-level MAPK mutation status was generally not prognostic for overall survival. Across eight Kaplan-Meier analyses stratified by ancestry, age, and FOLFOX exposure, we observed no statistically significant survival differences between MAPK-altered and non-altered groups. Small non-altered subgroups and broad confidence intervals limited power for some comparisons, and EOCRC NHW patients treated with FOLFOX exhibited a modest, non-significant trend toward worse survival in the MAPK-non-altered group. Overall, however, MAPK alteration status did not emerge as a robust survival biomarker in any stratum.

Finally, the conversational AI framework proved valuable as an exploratory engine. AI-guided analyses rapidly identified an ancestry-based survival advantage for FOLFOX-treated EO H/L patients compared with ancestry-matched NHW controls, confirmed that MSI stability and AKT3 mutation frequency did not differ meaningfully by treatment in select subgroups, and surfaced mutation-level disparities within MAPK-altered early-onset tumors (e.g., differential TP53, APC, KRAS, TCF7L2, PIK3CA, and NF1 frequencies between H/L and NHW cases). These AI-derived signals shaped our subsequent hypothesis-driven analyses.

### Biological implications of MAPK pathway alterations

The near-universal MAPK pathway alteration burden across CRC reinforces the concept that MAPK signaling represents a core dependency of colorectal tumorigenesis, spanning both canonical oncogenic drivers (KRAS, BRAF, NRAS) and a network of upstream receptors, adaptors, negative regulators, and effector kinases. However, the non-uniform distribution of individual gene alterations suggests that MAPK dysregulation is achieved through distinct molecular routes in different clinical contexts.

The enrichment of NF1 mutations in EO H/L tumors, especially in chemo-naïve cases, highlights the potential importance of disrupted negative regulation of RAS signaling in early-onset disease within this population. NF1 loss is known to augment RAS activity, and its co-occurrence with other MAPK drivers could amplify pathway signaling or modify sensitivity to targeted agents. Similarly, FGFR2, NTRK2, PDGFRB, and CRKL/SOS1 alterations implicate diverse receptor tyrosine kinases and adaptor proteins as alternative entry points into MAPK activation, particularly in older or untreated patients.

The age-associated increase in BRAF mutations, most prominent in LOCRC across ancestries, aligns with prior observations that BRAF-mutant CRC often presents at older ages and carries a distinct clinical phenotype. In our data, BRAF enrichment with age occurred against a background of stable KRAS and TP53 frequencies, suggesting that BRAF-driven MAPK activation represents one of several age-linked evolutionary trajectories rather than a universal feature of late-onset disease.

The context-specific enrichment of RPS6KA4 and MAPK3, both downstream effectors of ERK signaling, points toward variation not only in initiating events but also in terminal pathway branches. These genes may modulate transcriptional programs related to proliferation, stress responses, and chemoresistance. In EO H/L non-FOLFOX-treated tumors, co-occurrence of NF1, RPS6KA4, and MAPK3 alterations suggests a unique MAPK architecture characterized by dysregulated negative feedback and enhanced effector signaling.

These patterns support a model in which MAPK activation is a shared requirement across CRC, but the wiring diagram of the pathway, where and how it is perturbed, varies with age, ancestry, and treatment history. Such variation may have implications for response to targeted inhibitors and for combination strategies that seek to exploit specific vulnerabilities within MAPK signaling networks.

### Ancestry-specific genomic patterns and treatment context

Our analyses underscore the importance of considering ancestry and treatment context together when interrogating MAPK biology. The most striking ancestry-associated differences emerged in early-onset, chemo-naïve disease, where H/L tumors showed higher frequencies of NF1, MAPK3, and RPS6KA4 alterations than NHW tumors despite similar global mutation burdens. These findings suggest that early tumorigenic events in H/L individuals may preferentially involve deregulation of MAPK negative regulators and distal effectors, potentially reflecting differences in germline backgrounds, environmental exposures, or microenvironmental pressures.

In contrast, ancestry differences were attenuated among FOLFOX-treated EOCRC, where only PDGFRB exhibited significantly higher prevalence in H/L tumors. This pattern may indicate that chemotherapy exposure homogenizes observable MAPK architectures by applying selective pressure across ancestries or, alternatively, that treatment decisions introduce clinical selection biases that obscure underlying biological differences.

Among LOCRC, treatment context appeared to play a larger role than ancestry itself. Numerous MAPK-related genes showed higher mutation frequencies in non-FOLFOX-treated NHW tumors compared with treated counterparts, suggesting that either tumors with more complex MAPK architectures are less likely to receive FOLFOX or that such tumors arise in clinical contexts (e.g., comorbidities, performance status) that preclude intensive chemotherapy. H/L LOCRC tumors also exhibited treatment-linked enrichment of NTRK2 and PDGFRB in non-FOLFOX cases, consistent with a broader theme that chemo-naïve tumors carry a richer diversity of MAPK entry points and modulators.

Taken together, these results support the view that ancestry-specific MAPK patterns are most apparent when examined within narrowly defined clinical strata, particularly early-onset, untreated disease. They also emphasize that real-world treatment patterns and selection effects must be carefully considered when interpreting apparent genomic differences between populations.

### Implications for FOLFOX response and prognostic stratification

Contrary to expectations based on preclinical data linking MAPK activation to chemoresistance and poor outcomes, we found no consistent association between MAPK pathway mutation status and overall survival in any age-, ancestry-, or treatment-defined subgroup. The lack of prognostic impact at the pathway level likely reflects several factors.

First, because MAPK pathway alterations are almost universal, a simple binary classification (altered vs. not altered) compresses the diversity of MAPK architectures into a coarse metric with limited discriminatory power. Second, the functional impact of individual mutations varies widely: many alterations may be passenger events or may even attenuate signaling, while others drive strong activation. Third, survival outcomes are shaped by interactions between MAPK and other oncogenic pathways (WNT, PI3K, TGF-β), tumor microenvironment features, and treatment regimens beyond FOLFOX, none of which are fully captured by a single pathway-level variable.

The modest, non-significant trend toward poorer outcomes in MAPK-non-altered EOCRC NHW patients treated with FOLFOX hints that tumors lacking canonical MAPK alterations may rely on alternative resistance mechanisms, but our data are underpowered to draw firm conclusions. In aggregate, these findings suggest that MAPK mutation status alone is unlikely to serve as a stand-alone prognostic marker for FOLFOX response. Instead, future work should explore multi-gene or co-mutation signatures, potentially integrating NF1, BRAF, PDGFRB, RPS6KA4, and other nodes with MSI, TMB, and clinical features to construct more nuanced models of chemotherapy sensitivity.

Importantly, our conversational AI analyses also showed that ancestry itself, independent of MAPK status, can be associated with survival differences in specific contexts, as illustrated by the better outcomes observed in EO H/L vs. EO NHW patients treated with FOLFOX. These findings reinforce that pathway-focused biomarkers must ultimately be interpreted alongside social, environmental, and systemic determinants of health to avoid oversimplified conclusions about biology-driven disparities.

### AI-HOPE-MAPK as an enabling technology

The AI-HOPE and AI-HOPE-MAPK platforms were central to this study. By embedding harmonized clinical, genomic, ancestry, and treatment data within a conversational analytic environment, the system allowed investigators to pose complex questions, such as “compare MAPK-altered early-onset H/L tumors not treated with FOLFOX to MAPK-altered NHW counterparts and identify genes that differ significantly”, using natural language rather than bespoke code.

This capability yielded several advantages: a) Rapid hypothesis generation: AI-guided queries quickly surfaced context-specific signals (e.g., NF1 and RPS6KA4 enrichment in EO H/L non-FOLFOX tumors, BRAF enrichment in LO disease, ancestry-linked co-mutation differences in MAPK-altered EOCRC) that would have been laborious to identify manually. b) Efficient validation of null results: The platform confirmed that MSI stability, AKT3 mutation frequency, and PDGFRB mutation prevalence did not differ meaningfully between select treatment or ancestry strata, allowing us to de-prioritize these hypotheses early. c) Transparent cohort construction: AI-generated cohort definitions and 2×2 tables were cross-checked against conventional R scripts and statistical tests, reducing manual data wrangling and minimizing transcription errors. d) Equity-focused analytics: By design, the system facilitated ancestry-aware and age-aware stratification, ensuring that H/L populations, often underrepresented in genomic datasets, were interrogated with the same granularity as NHW populations.

Crucially, we treated AI-HOPE-MAPK as an analytic accelerator rather than a replacement for traditional statistics. All AI-suggested findings were validated using standard methods, and several AI-flagged trends did not withstand formal testing, highlighting the importance of coupling exploratory AI with rigorous biostatistical oversight.

### Limitations and future directions

This study has several limitations. Despite the large overall cohort, many clinically important subgroups, especially H/L patients lacking FOLFOX exposure or carrying specific MAPK alterations, remained relatively small, leading to wide confidence intervals and limited power for survival and gene-frequency comparisons. Our analyses were restricted to coding single-nucleotide variants and small indels; copy-number alterations, structural variants, gene fusions (particularly relevant for NTRK genes), and expression or phosphoproteomic readouts of MAPK activation were not systematically incorporated.

Treatment annotation, while sufficient to classify FOLFOX exposure, lacked granular detail on dose intensity, treatment duration, combination partners, and subsequent lines of therapy, all of which can modulate outcomes and interact with MAPK biology. The retrospective, multi-cohort design introduces potential confounding from unmeasured variables such as comorbidities, performance status, socioeconomic factors, and center-specific treatment practices, factors that are particularly salient when interpreting ancestry-linked differences.

From a methodological standpoint, ancestry assignment relied on a combination of self-reported ethnicity and surname-based inference, which may misclassify some individuals and does not capture intra-group diversity (e.g., Caribbean vs. Mexican heritage). Likewise, AI-HOPE-MAPK depends on the quality and completeness of source annotations; systematic biases or errors in the underlying datasets can propagate through AI-assisted analyses. Future iterations of the platform should incorporate data provenance tracking, automated uncertainty flagging, and fairness auditing to ensure that AI workflows do not inadvertently perpetuate existing inequities.

Future research should pursue several directions. First, prospective, ancestrally diverse cohorts with standardized treatment and biospecimen collection will be crucial to validate the gene-level MAPK signals identified here, particularly NF1, PDGFRB, MAPK3, RPS6KA4, NTRK2, and RAF1/SOS1/CRKL. Second, integrating multi-omic measures of pathway activity, including RNA expression signatures, phospho-proteomics, and spatial profiling, with mutational data may yield more predictive MAPK activation scores than genomic status alone. Third, mechanistic studies using in vitro and in vivo models representative of H/L and NHW genetic backgrounds are needed to elucidate how specific MAPK alterations interact with chemotherapy and with other oncogenic pathways. Finally, extension of AI-HOPE-MAPK to model crosstalk between MAPK and WNT, PI3K, TGF-β, and immune pathways could uncover composite biomarkers that better capture the complexity of CRC biology.

## 5. Conclusion

In summary, this AI-enabled precision oncology study reveals that MAPK pathway disruption is a near-universal feature of colorectal cancer across age, ancestry, and FOLFOX treatment strata, but that the manner in which the pathway is altered, through specific genes, receptors, adaptors, and effectors, varies in a context-dependent manner. Early-onset, chemo-naïve H/L tumors are enriched for NF1, MAPK3, and RPS6KA4 alterations, while late-onset and non-FOLFOX-treated NHW tumors harbor distinct constellations of MAPK modulators, and BRAF burden increases with age in both populations. Despite this rich gene-level heterogeneity, MAPK pathway mutation status alone does not consistently predict overall survival in any age-ancestry-treatment subgroup, highlighting the limitations of binary pathway metrics and the need for more nuanced, multi-gene biomarkers.

Beyond these biological insights, our work demonstrates how conversational AI systems such as AI-HOPE-MAPK can harmonize complex clinical-genomic datasets, accelerate hypothesis generation, and support equity-focused, ancestry-aware biomarker discovery. By enabling natural language-driven cohort construction and rapid exploratory analyses, AI-HOPE-MAPK functioned as a catalyst for traditional analytics rather than a replacement, helping to prioritize meaningful signals and efficiently dismiss non-informative hypotheses.

As precision oncology moves toward increasingly granular, population-conscious models of care, frameworks that integrate pathway biology, treatment context, and ancestry within transparent AI workflows will be essential. Continued development of AI-HOPE-MAPK, coupled with mechanistic experimentation and prospective validation in diverse patient populations, will be critical for translating MAPK-focused insights into clinically actionable strategies, and for ensuring that these advances benefit patients historically underrepresented in cancer genomics research.

## Data Availability

All data used in the present study is publicly available at https://www.cbioportal.org/ and https://genie.cbioportal.org. The datasets used in our study were aggregated/summary data, and no individual-level data were used. Additional data can be provided upon reasonable request to the authors.

## Supplementary Materials

**Table S1.**
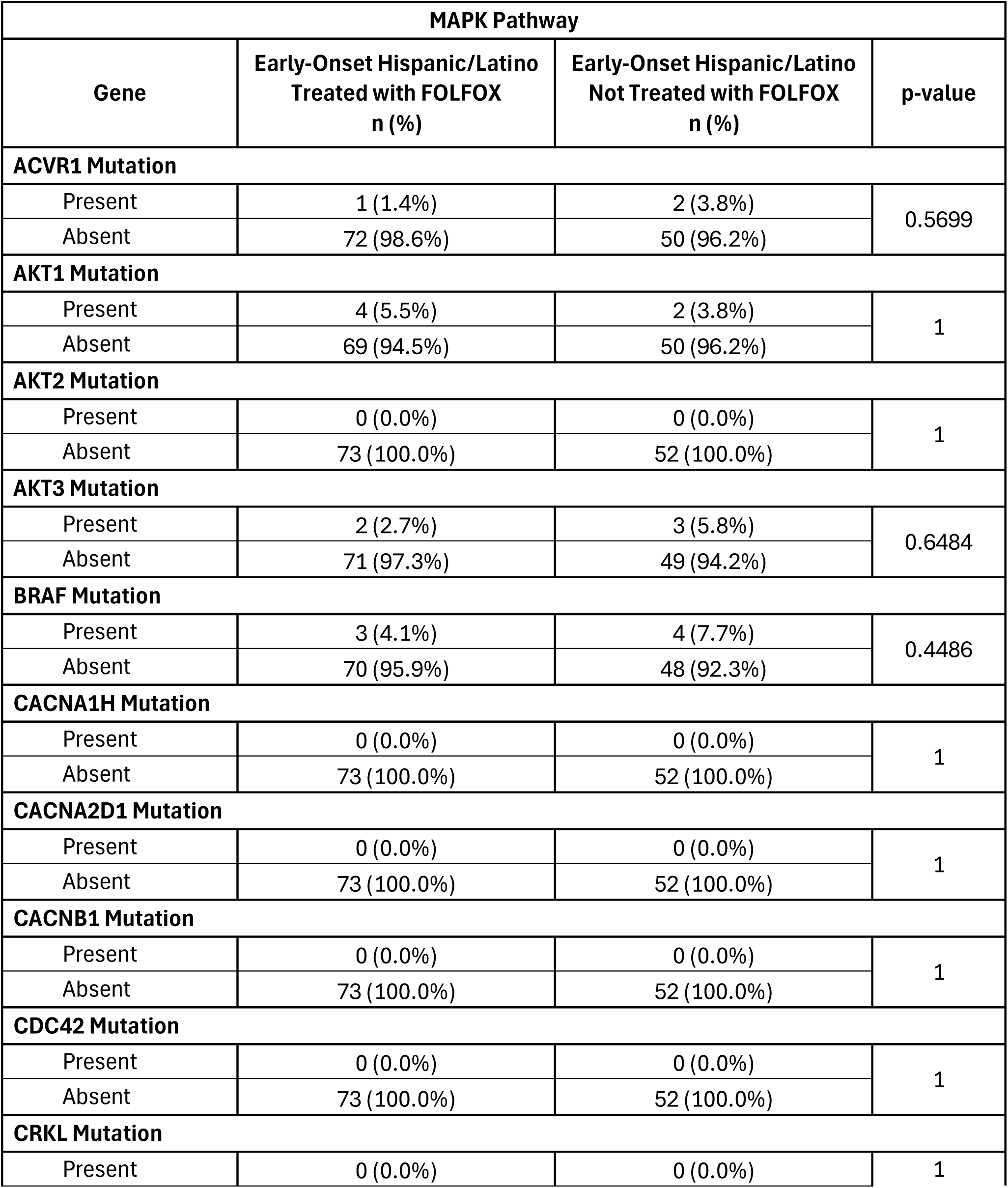
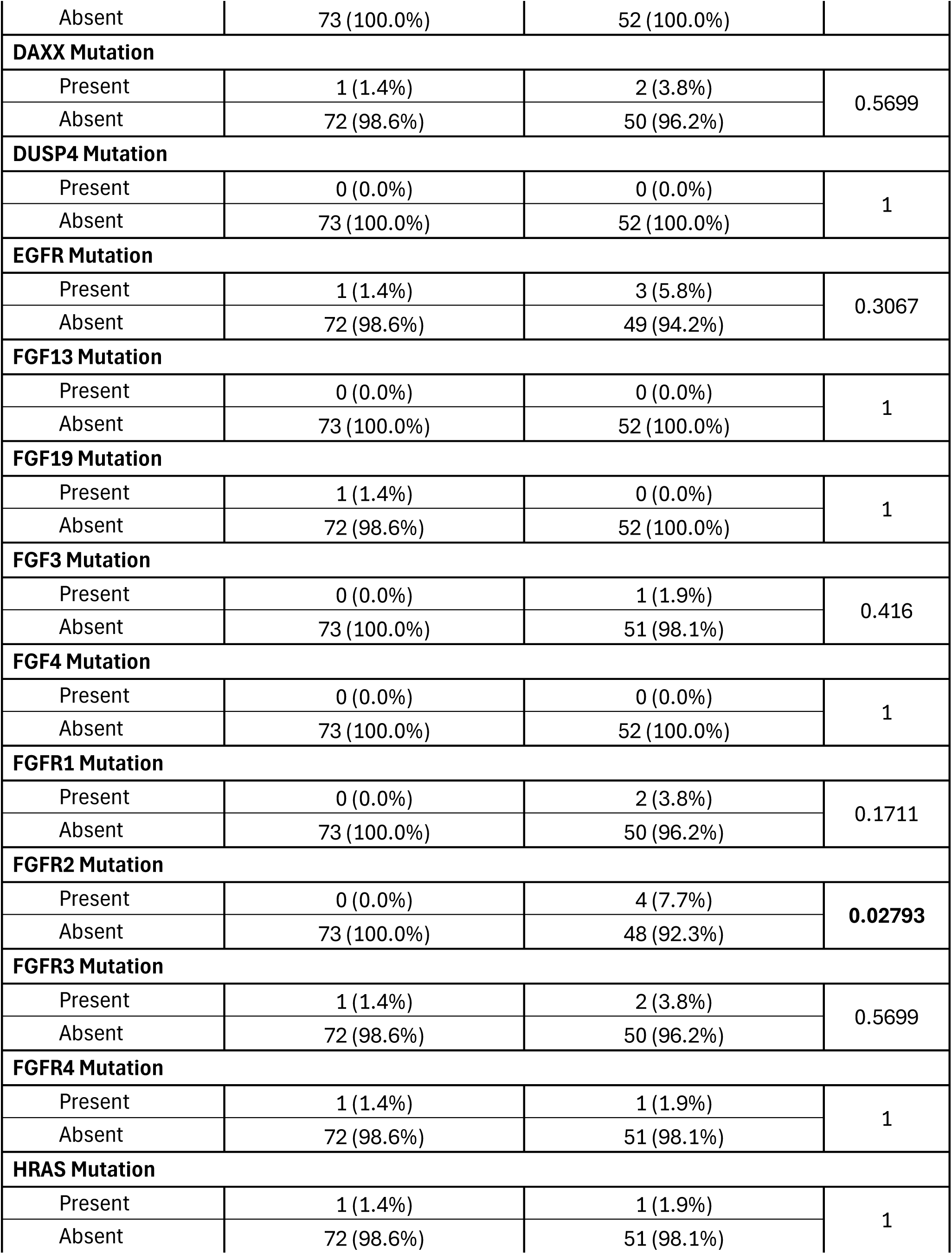
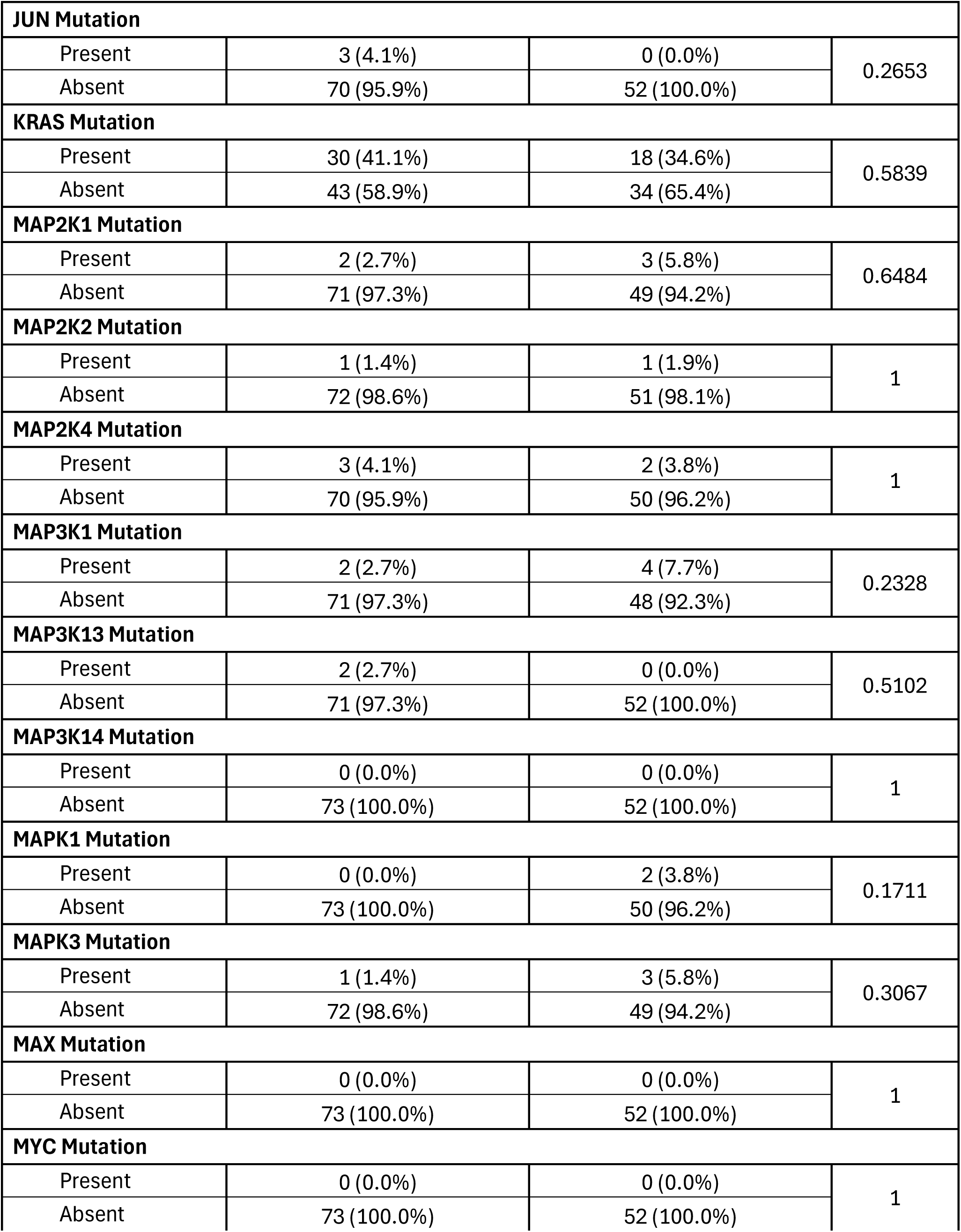
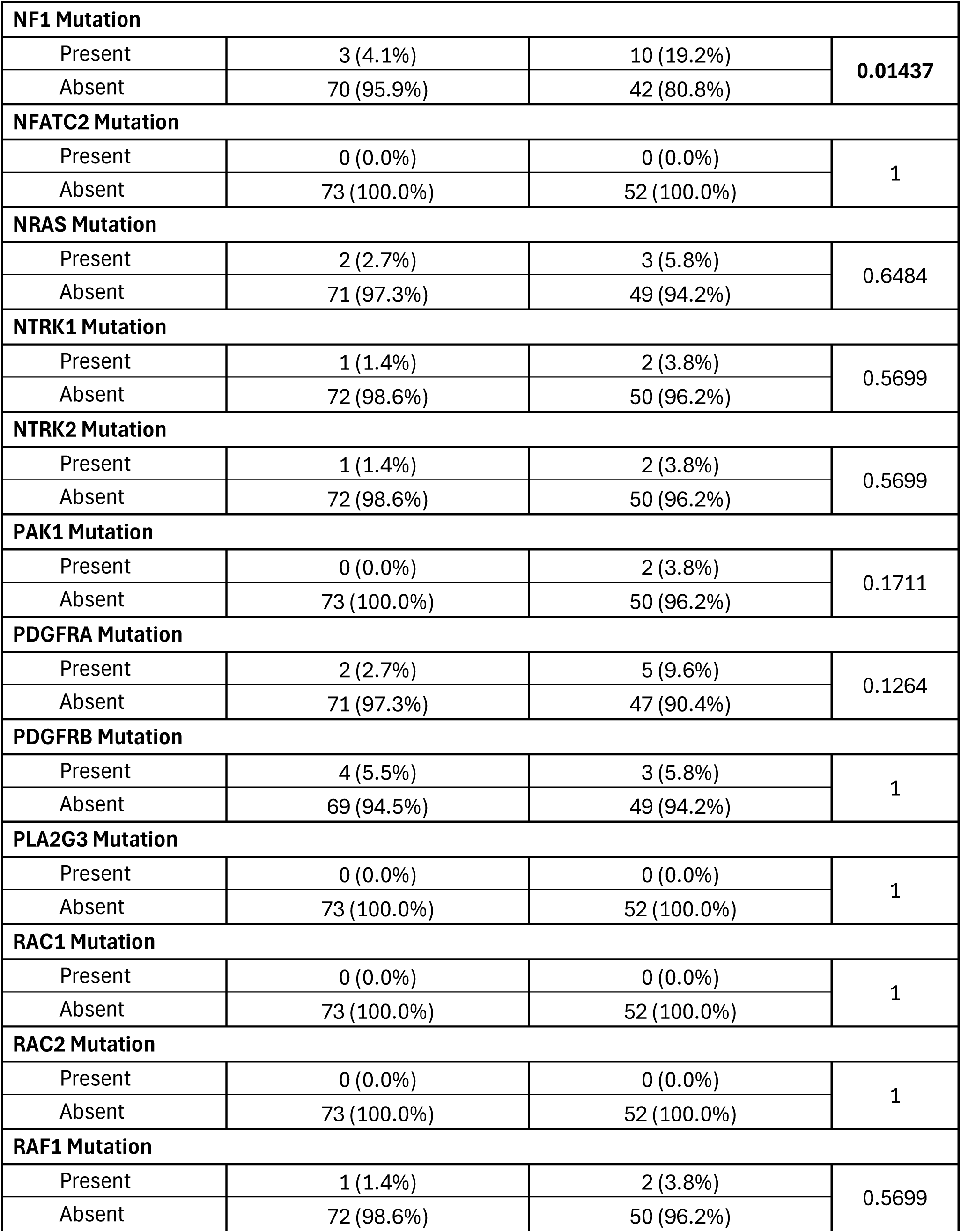
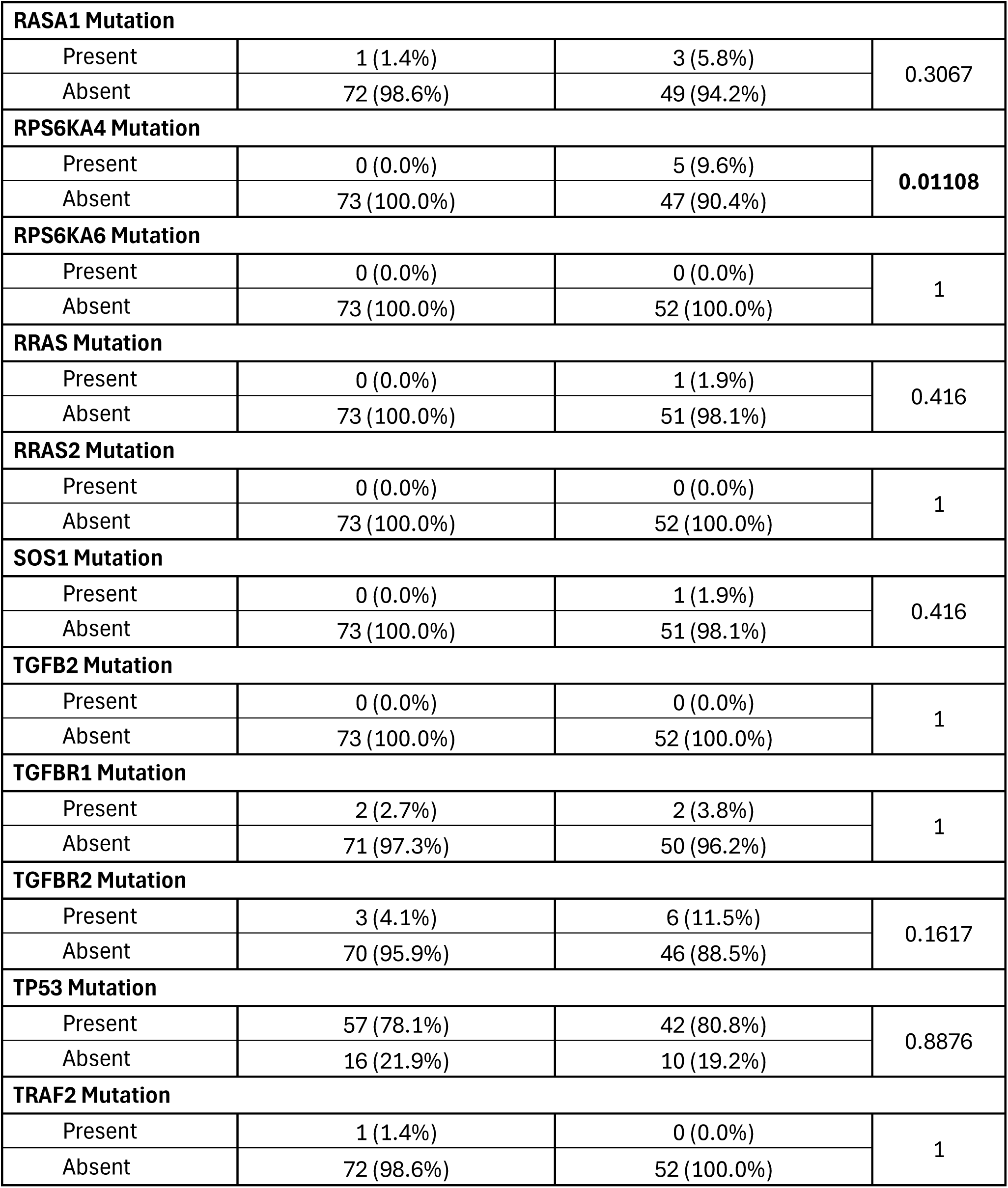
Comparison of Early-Onset Hispanic/Latino (H/L) Patients Treated with FOLFOX versus Not Treated with FOLFOX.

**Table S2.**
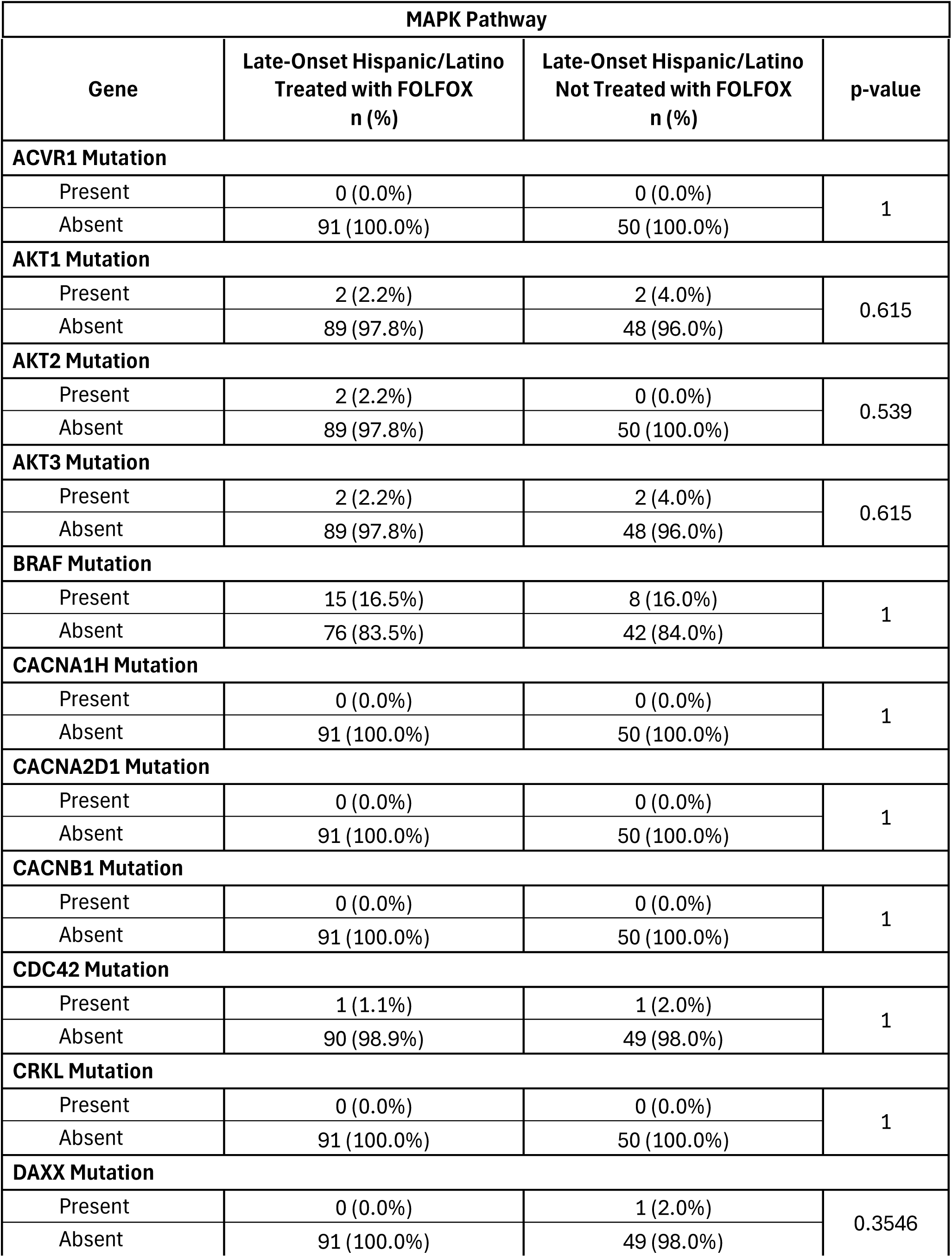
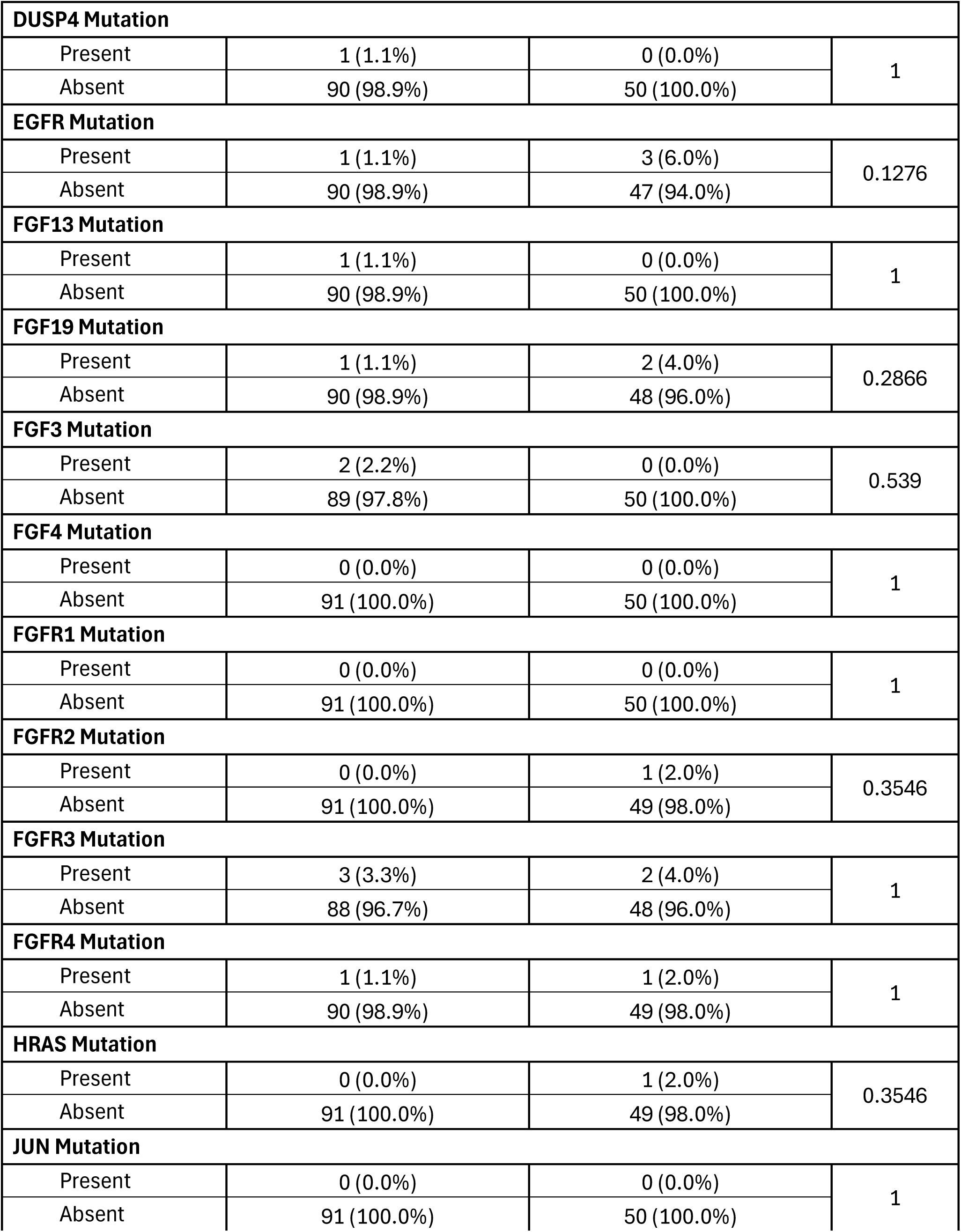
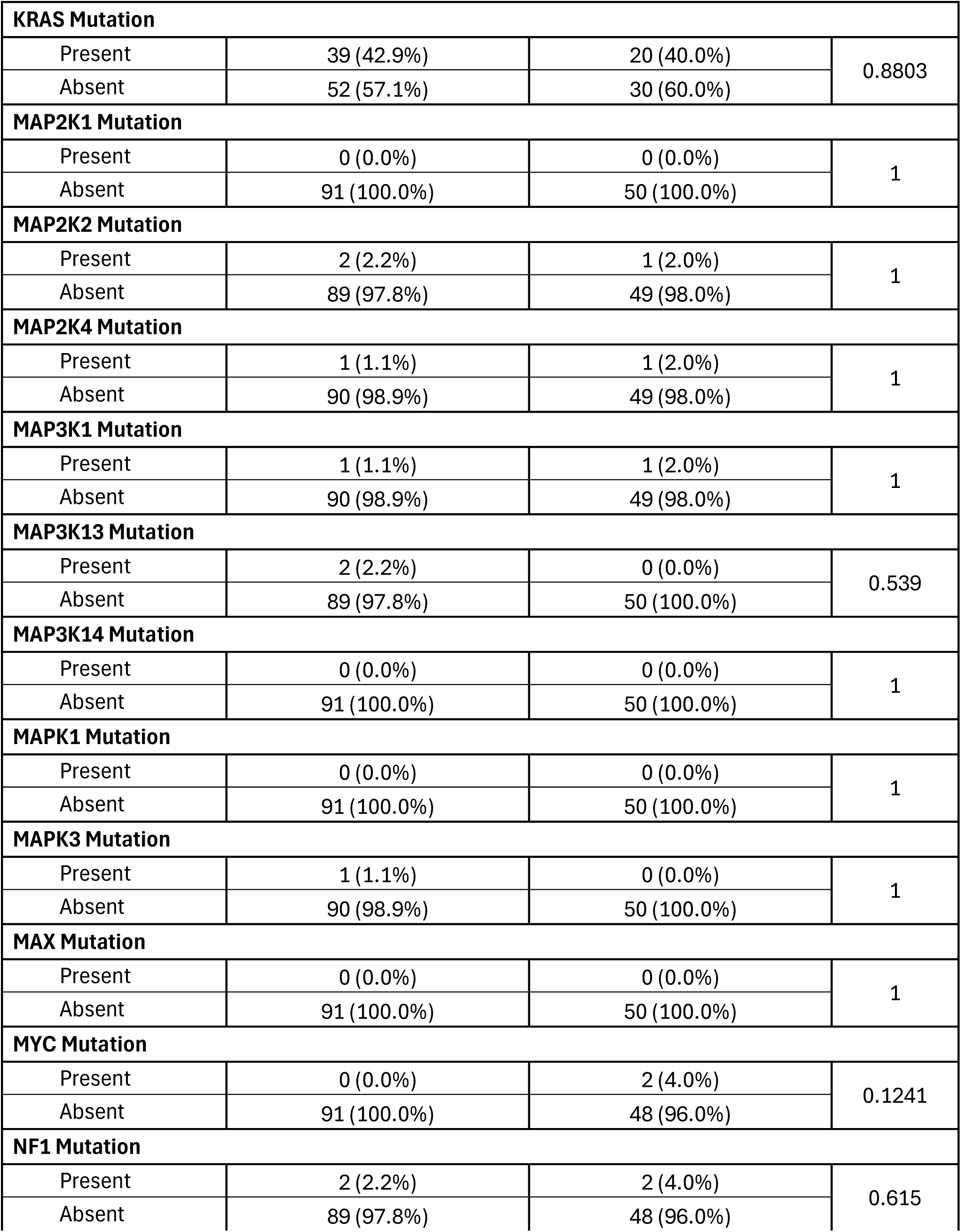
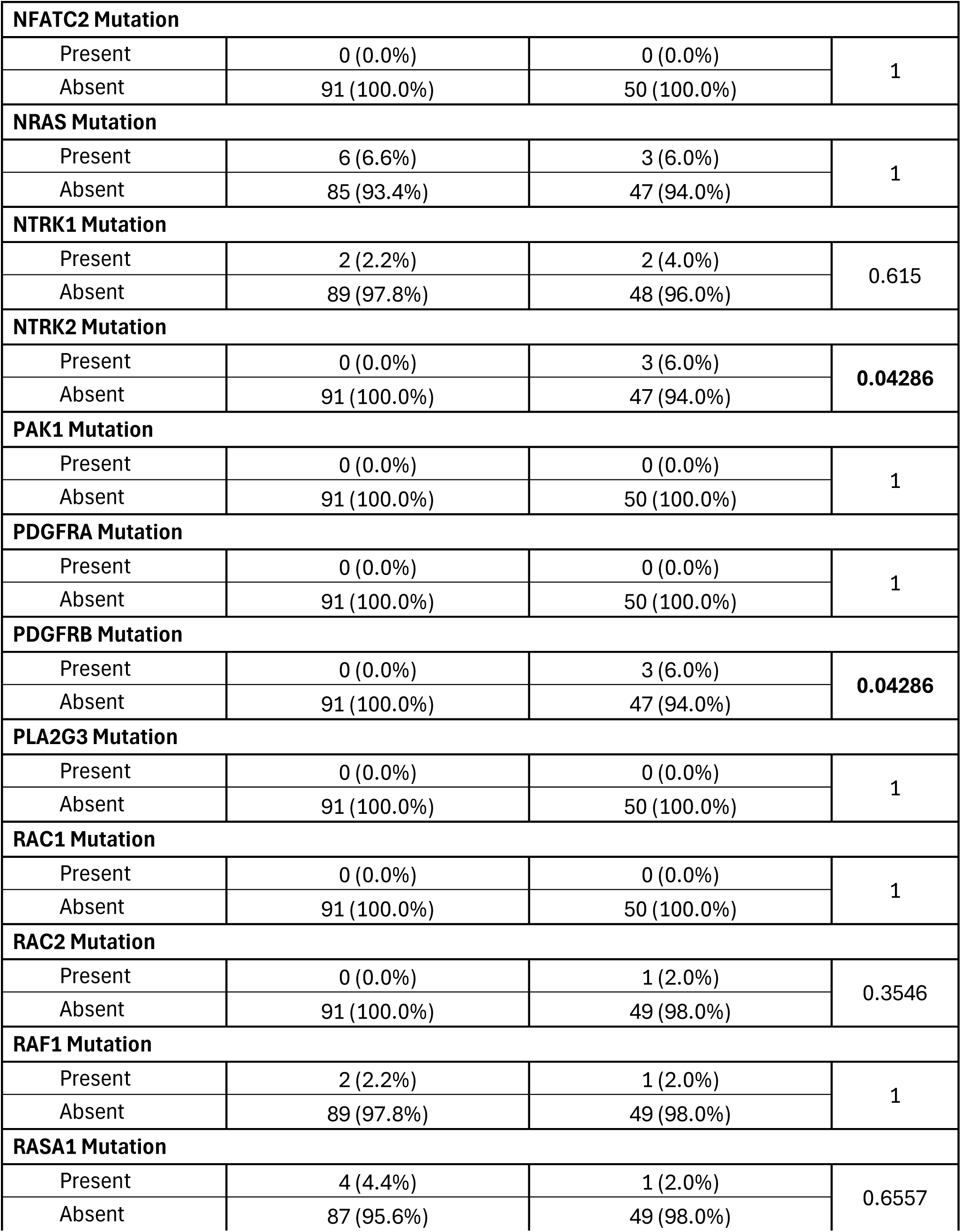
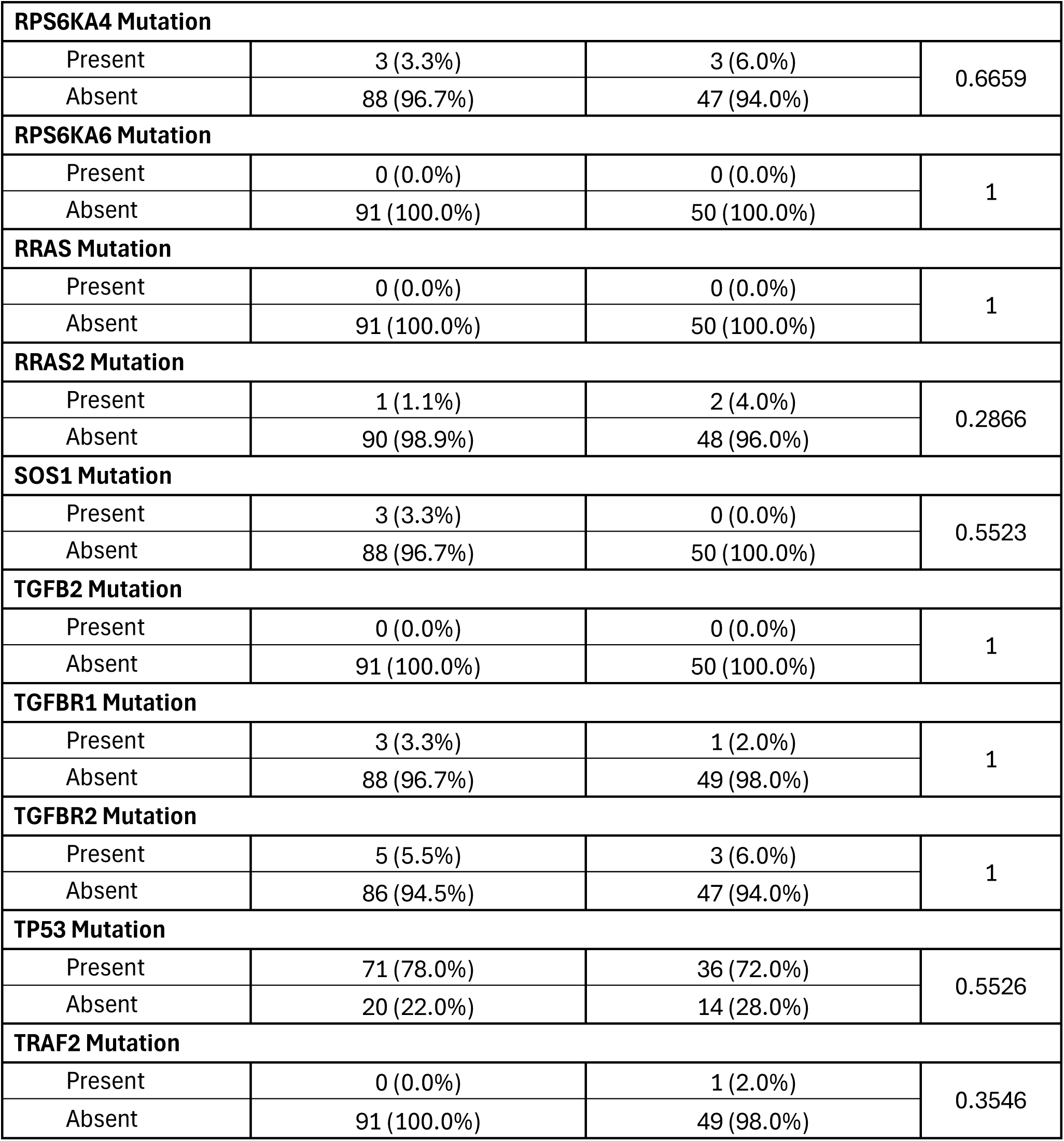
Comparison of Late-Onset Hispanic/Latino (H/L) Patients Treated with FOLFOX versus Not Treated with FOLFOX.

**Table S3.** Comparison of Early-Onset Non-Hispanic White (NHW) Patients Treated with FOLFOX versus Not Treated with FOLFOX.

| MAPK Pathway |  |  |  |
| --- | --- | --- | --- |
| Gene | Early-Onset NHW<br>Treated with FOLFOX<br>n (%) | Early-Onset NHW<br>Not Treated with FOLFOX<br>n (%) | p-value |
| ACVR1 Mutation |  |  |  |
| Present | 4 (1.1%) | 5 (1.7%) | 0.522 |
| Absent | 371 (98.9%) | 297 (98.3%) |  |
| AKT1 Mutation |  |  |  |
| Present | 9 (2.4%) | 4 (1.3%) | 0.4034 |
| Absent | 366 (97.6%) | 298 (98.7%) |  |
| AKT2 Mutation |  |  |  |
| Present | 2 (0.5%) | 2 (0.7%) | 1 |
| Absent | 373 (99.5%) | 300 (99.3%) |  |
| AKT3 Mutation |  |  |  |
| Present | 3 (0.8%) | 9 (3.0%) | 0.04069 |
| Absent | 372 (99.2%) | 293 (97.0%) |  |
| BRAF Mutation |  |  |  |
| Present | 27 (7.2%) | 24 (7.9%) | 0.8262 |
| Absent | 348 (92.8%) | 278 (92.1%) |  |
| CACNA1H Mutation |  |  |  |
| Present | 0 (0.0%) | 0 (0.0%) | 1 |
| Absent | 375 (100.0%) | 302 (100.0%) |  |
| CACNA2D1 Mutation |  |  |  |
| Present | 0 (0.0%) | 0 (0.0%) | 1 |
| Absent | 375 (100.0%) | 302 (100.0%) |  |
| CACNB1 Mutation |  |  |  |
| Present | 0 (0.0%) | 0 (0.0%) | 1 |
| Absent | 375 (100.0%) | 302 (100.0%) |  |
| CDC42 Mutation |  |  |  |
| Present | 0 (0.0%) | 2 (0.7%) | 0.1986 |
| Absent | 375 (100.0%) | 300 (99.3%) |  |
| CRKL Mutation |  |  |  |
| Present | 1 (0.3%) | 3 (1.0%) | 0.3291 |
| Absent | 374 (99.7%) | 299 (99.0%) |  |
| DAXX Mutation |  |  |  |
| Present | 3 (0.8%) | 6 (2.0%) | 0.1977 |
| Absent | 372 (99.2%) | 296 (98.0%) |  |
| DUSP4 Mutation |  |  |  |
| Present | 1 (0.3%) | 1 (0.3%) | 1 |
| Absent | 374 (99.7%) | 301 (99.7%) |  |
| EGFR Mutation |  |  |  |
| Present | 5 (1.3%) | 5 (1.7%) | 0.98 |
| Absent | 370 (98.7%) | 297 (98.3%) |  |
| FGF13 Mutation |  |  |  |
| Present | 0 (0.0%) | 0 (0.0%) | 1 |
| Absent | 375 (100.0%) | 302 (100.0%) |  |
| FGF19 Mutation |  |  |  |
| Present | 3 (0.8%) | 4 (1.3%) | 0.706 |
| Absent | 372 (99.2%) | 298 (98.7%) |  |
| FGF3 Mutation |  |  |  |
| Present | 4 (1.1%) | 5 (1.7%) | 0.522 |
| Absent | 371 (98.9%) | 297 (98.3%) |  |
| FGF4 Mutation |  |  |  |
| Present | 0 (0.0%) | 5 (1.7%) | 0.01734 |
| Absent | 375 (100.0%) | 297 (98.3%) |  |
| FGFR1 Mutation |  |  |  |
| Present | 10 (2.7%) | 6 (2.0%) | 0.7456 |
| Absent | 365 (97.3%) | 296 (98.0%) |  |
| FGFR2 Mutation |  |  |  |
| Present | 5 (1.3%) | 9 (3.0%) | 0.2206 |
| Absent | 370 (98.7%) | 293 (97.0%) |  |
| FGFR3 Mutation |  |  |  |
| Present | 5 (1.3%) | 11 (3.6%) | 0.08699 |
| Absent | 370 (98.7%) | 291 (96.4%) |  |
| FGFR4 Mutation |  |  |  |
| Present | 6 (1.6%) | 5 (1.7%) | 1 |
| Absent | 369 (98.4%) | 297 (98.3%) |  |
| HRAS Mutation |  |  |  |
| Present | 3 (0.8%) | 2 (0.7%) | 1 |
| Absent | 372 (99.2%) | 300 (99.3%) |  |
| JUN Mutation |  |  |  |
| Present | 12 (3.2%) | 2 (0.7%) | 0.02726 |
| Absent | 363 (96.8%) | 300 (99.3%) |  |
| KRAS Mutation |  |  |  |
| Present | 163 (43.5%) | 124 (41.1%) | 0.5811 |
| Absent | 212 (56.5%) | 178 (58.9%) |  |
| MAP2K1 Mutation |  |  |  |
| Present | 6 (1.6%) | 4 (1.3%) | 1 |
| Absent | 369 (98.4%) | 298 (98.7%) |  |
| MAP2K2 Mutation |  |  |  |
| Present | 3 (0.8%) | 1 (0.3%) | 0.6326 |
| Absent | 372 (99.2%) | 301 (99.7%) |  |
| MAP2K4 Mutation |  |  |  |
| Present | 6 (1.6%) | 9 (3.0%) | 0.3421 |
| Absent | 369 (98.4%) | 293 (97.0%) |  |
| MAP3K1 Mutation |  |  |  |
| Present | 10 (2.7%) | 12 (4.0%) | 0.4622 |
| Absent | 365 (97.3%) | 290 (96.0%) |  |
| MAP3K13 Mutation |  |  |  |
| Present | 5 (1.3%) | 9 (3.0%) | 0.2206 |
| Absent | 370 (98.7%) | 293 (97.0%) |  |
| MAP3K14 Mutation |  |  |  |
| Present | 1 (0.3%) | 0 (0.0%) | 1 |
| Absent | 374 (99.7%) | 302 (100.0%) |  |
| MAPK1 Mutation |  |  |  |
| Present | 3 (0.8%) | 3 (1.0%) | 1 |
| Absent | 372 (99.2%) | 299 (99.0%) |  |
| MAPK3 Mutation |  |  |  |
| Present | 1 (0.3%) | 3 (1.0%) | 0.3291 |
| Absent | 374 (99.7%) | 299 (99.0%) |  |
| MAX Mutation |  |  |  |
| Present | 4 (1.1%) | 1 (0.3%) | 0.3877 |
| Absent | 371 (98.9%) | 301 (99.7%) |  |
| MYC Mutation |  |  |  |
| Present | 2 (0.5%) | 1 (0.3%) | 1 |
| Absent | 373 (99.5%) | 301 (99.7%) |  |
| NF1 Mutation |  |  |  |
| Present | 17 (4.5%) | 18 (6.0%) | 0.5099 |
| Absent | 358 (95.5%) | 284 (94.0%) |  |
| NFATC2 Mutation |  |  |  |
| Present | 0 (0.0%) | 0 (0.0%) | 1 |
| Absent | 375 (100.0%) | 302 (100.0%) |  |
| NRAS Mutation |  |  |  |
| Present | 12 (3.2%) | 6 (2.0%) | 0.358 |
| Absent | 363 (96.8%) | 296 (98.0%) |  |
| NTRK1 Mutation |  |  |  |
| Present | 10 (2.7%) | 9 (3.0%) | 0.9909 |
| Absent | 365 (97.3%) | 293 (97.0%) |  |
| NTRK2 Mutation |  |  |  |
| Present | 5 (1.3%) | 8 (2.6%) | 0.3379 |
| Absent | 370 (98.7%) | 294 (97.4%) |  |
| PAK1 Mutation |  |  |  |
| Present | 7 (1.9%) | 3 (1.0%) | 0.5245 |
| Absent | 368 (98.1%) | 299 (99.0%) |  |
| PDGFRA Mutation |  |  |  |
| Present | 14 (3.7%) | 16 (5.3%) | 0.4263 |
| Absent | 361 (96.3%) | 286 (94.7%) |  |
| PDGFRB Mutation |  |  |  |
| Present | 3 (0.8%) | 7 (2.3%) | 0.1197 |
| Absent | 372 (99.2%) | 295 (97.7%) |  |
| PLA2G3 Mutation |  |  |  |
| Present | 0 (0.0%) | 0 (0.0%) | 1 |
| Absent | 375 (100.0%) | 302 (100.0%) |  |
| RAC1 Mutation |  |  |  |
| Present | 1 (0.3%) | 3 (1.0%) | 0.3291 |
| Absent | 374 (99.7%) | 299 (99.0%) |  |
| RAC2 Mutation |  |  |  |
| Present | 2 (0.5%) | 2 (0.7%) | 1 |
| Absent | 373 (99.5%) | 300 (99.3%) |  |
| RAF1 Mutation |  |  |  |
| Present | 9 (2.4%) | 9 (3.0%) | 0.8211 |
| Absent | 366 (97.6%) | 293 (97.0%) |  |
| RASA1 Mutation |  |  |  |
| Present | 9 (2.4%) | 14 (4.6%) | 0.1667 |
| Absent | 366 (97.6%) | 288 (95.4%) |  |
| RPS6KA4 Mutation |  |  |  |
| Present | 5 (1.3%) | 8 (2.6%) | 0.3379 |
| Absent | 370 (98.7%) | 294 (97.4%) |  |
| RPS6KA6 Mutation |  |  |  |
| Present | 0 (0.0%) | 1 (0.3%) | 0.4461 |
| Absent | 375 (100.0%) | 301 (99.7%) |  |
| RRAS Mutation |  |  |  |
| Present | 4 (1.1%) | 0 (0.0%) | 0.1326 |
| Absent | 371 (98.9%) | 302 (100.0%) |  |
| RRAS2 Mutation |  |  |  |
| Present | 0 (0.0%) | 4 (1.3%) | 0.03916 |
| Absent | 375 (100.0%) | 298 (98.7%) |  |
| SOS1 Mutation |  |  |  |
| Present | 7 (1.9%) | 11 (3.6%) | 0.2351 |
| Absent | 368 (98.1%) | 291 (96.4%) |  |
| TGFB2 Mutation |  |  |  |
| Present | 0 (0.0%) | 0 (0.0%) | 1 |
| Absent | 375 (100.0%) | 302 (100.0%) |  |
| TGFBR1 Mutation |  |  |  |
| Present | 8 (2.1%) | 9 (3.0%) | 0.6506 |
| Absent | 367 (97.9%) | 293 (97.0%) |  |
| TGFBR2 Mutation |  |  |  |
| Present | 11 (2.9%) | 16 (5.3%) | 0.1721 |
| Absent | 364 (97.1%) | 286 (94.7%) |  |
| TP53 Mutation |  |  |  |
| Present | 296 (78.9%) | 228 (75.5%) | 0.3319 |
| Absent | 79 (21.1%) | 74 (24.5%) |  |
| TRAF2 Mutation |  |  |  |
| Present | 5 (1.3%) | 4 (1.3%) | 1 |
| Absent | 370 (98.7%) | 298 (98.7%) |  |

**Table S4.**
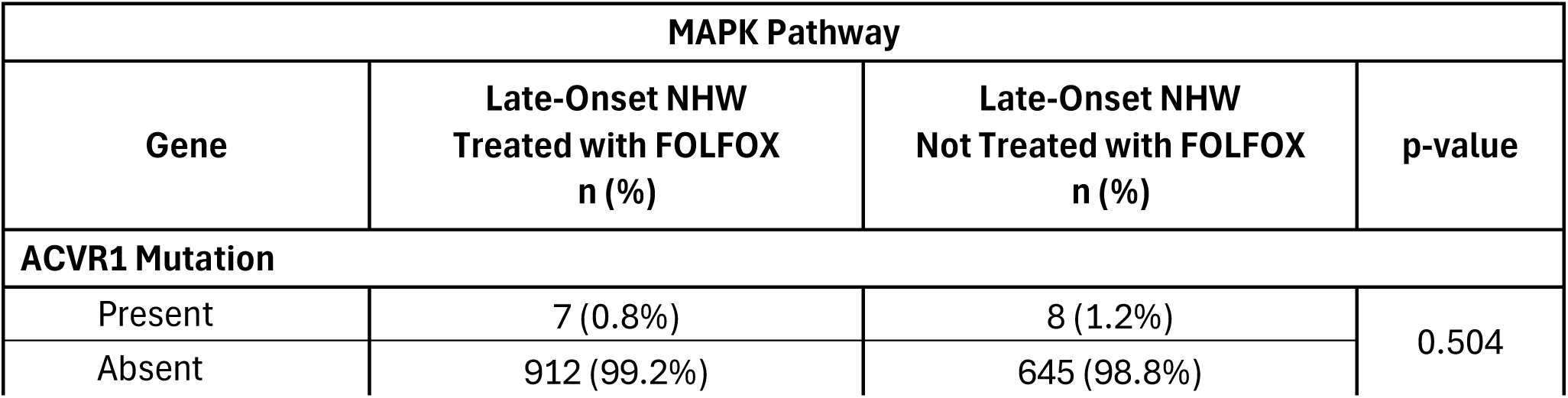
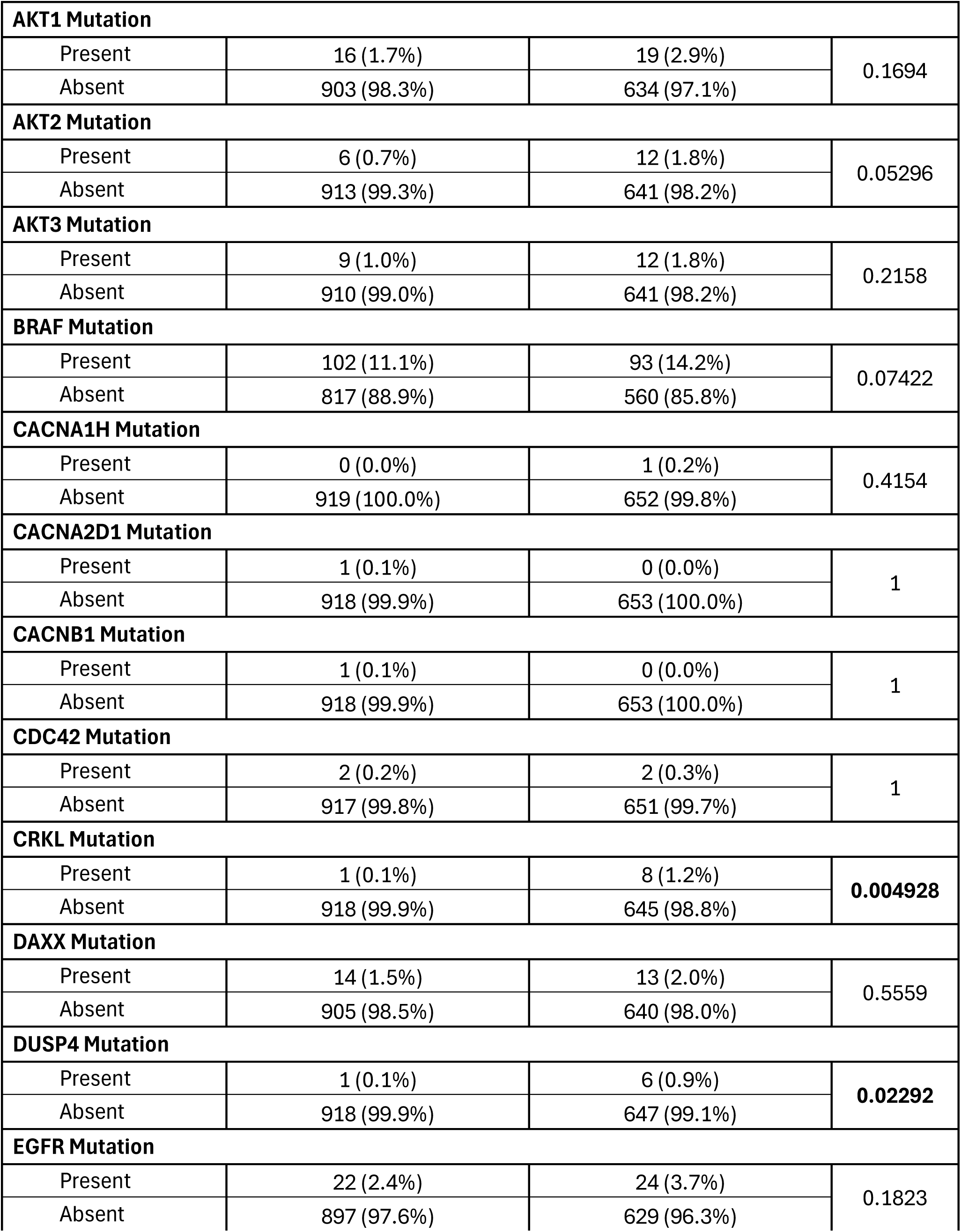
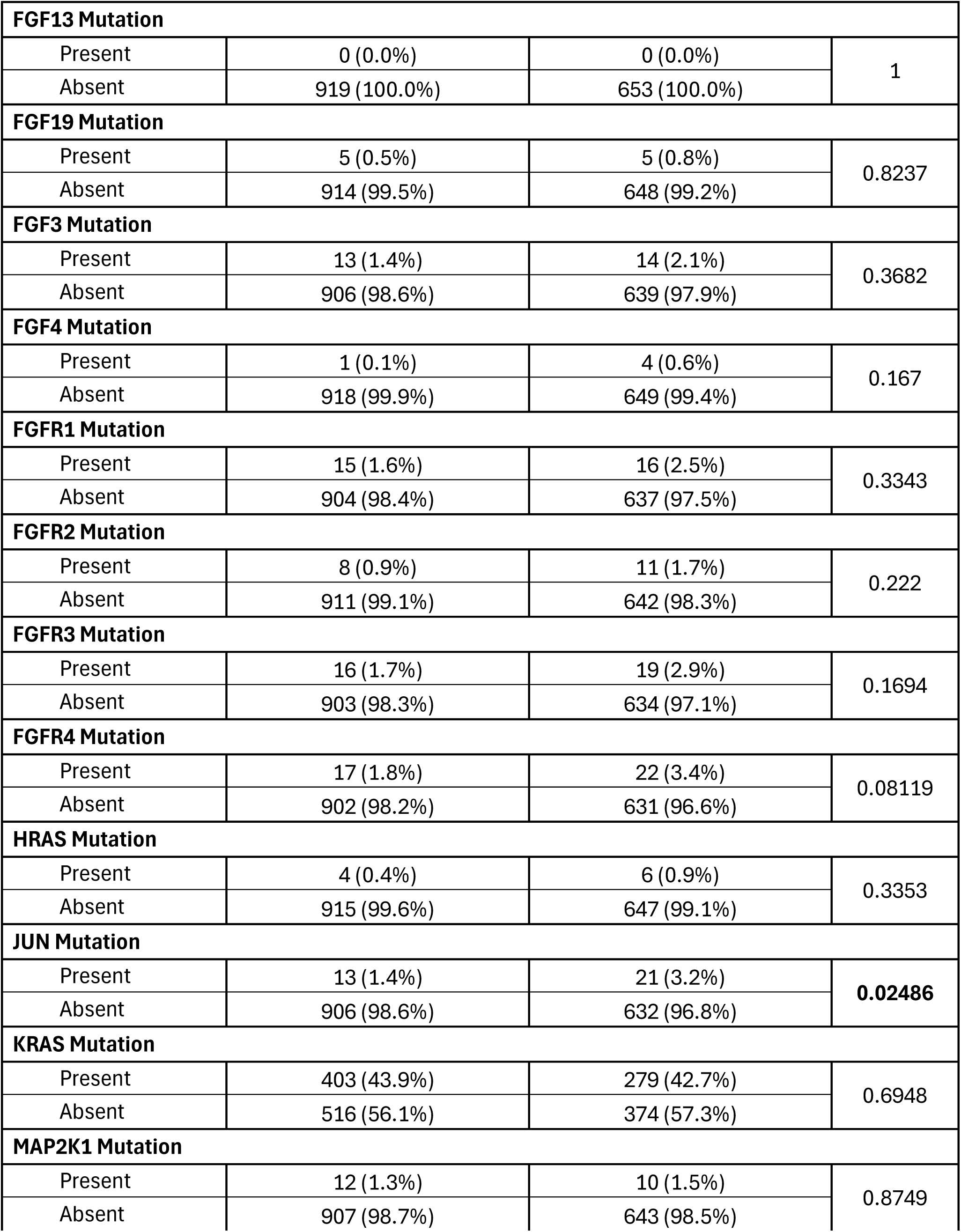
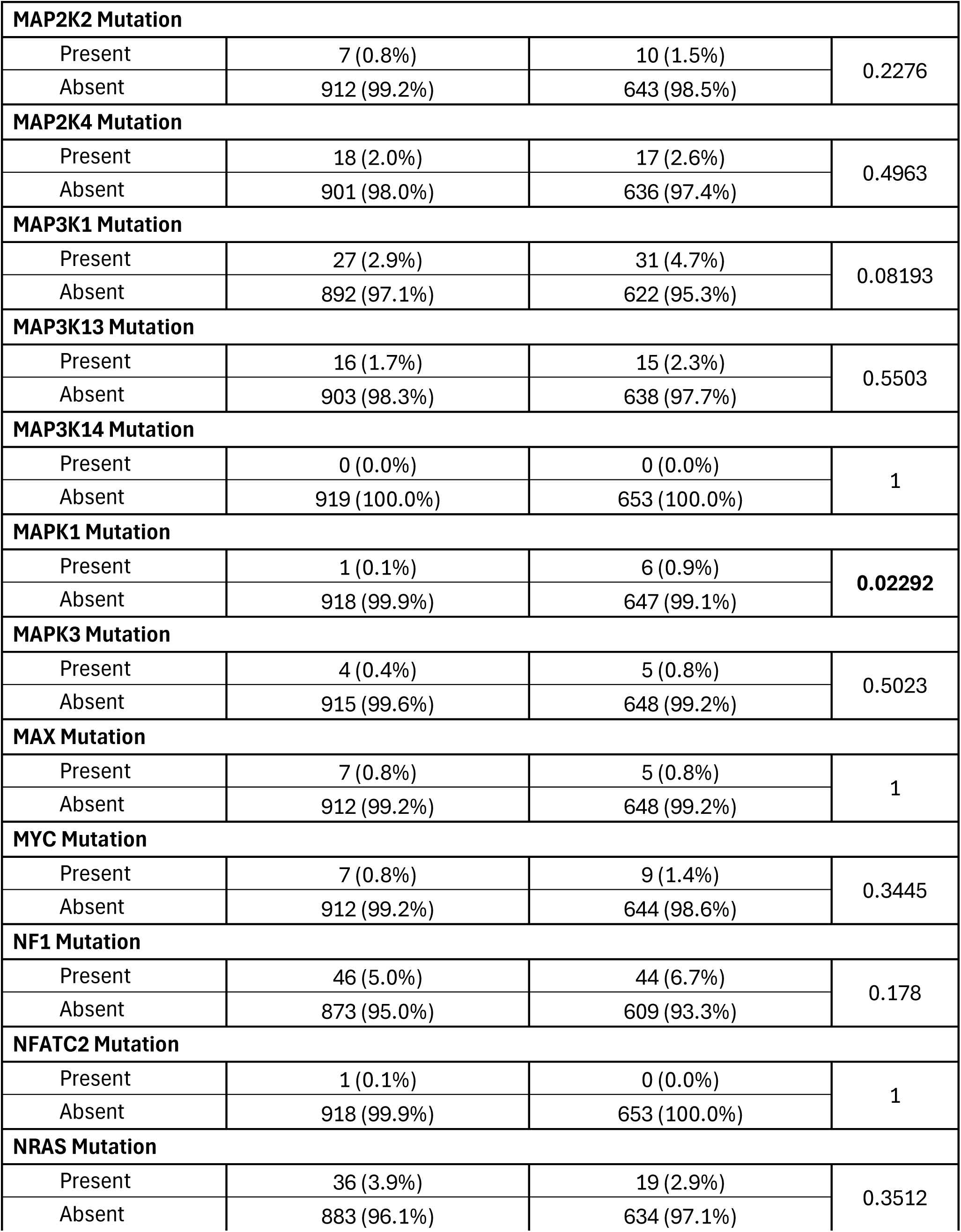
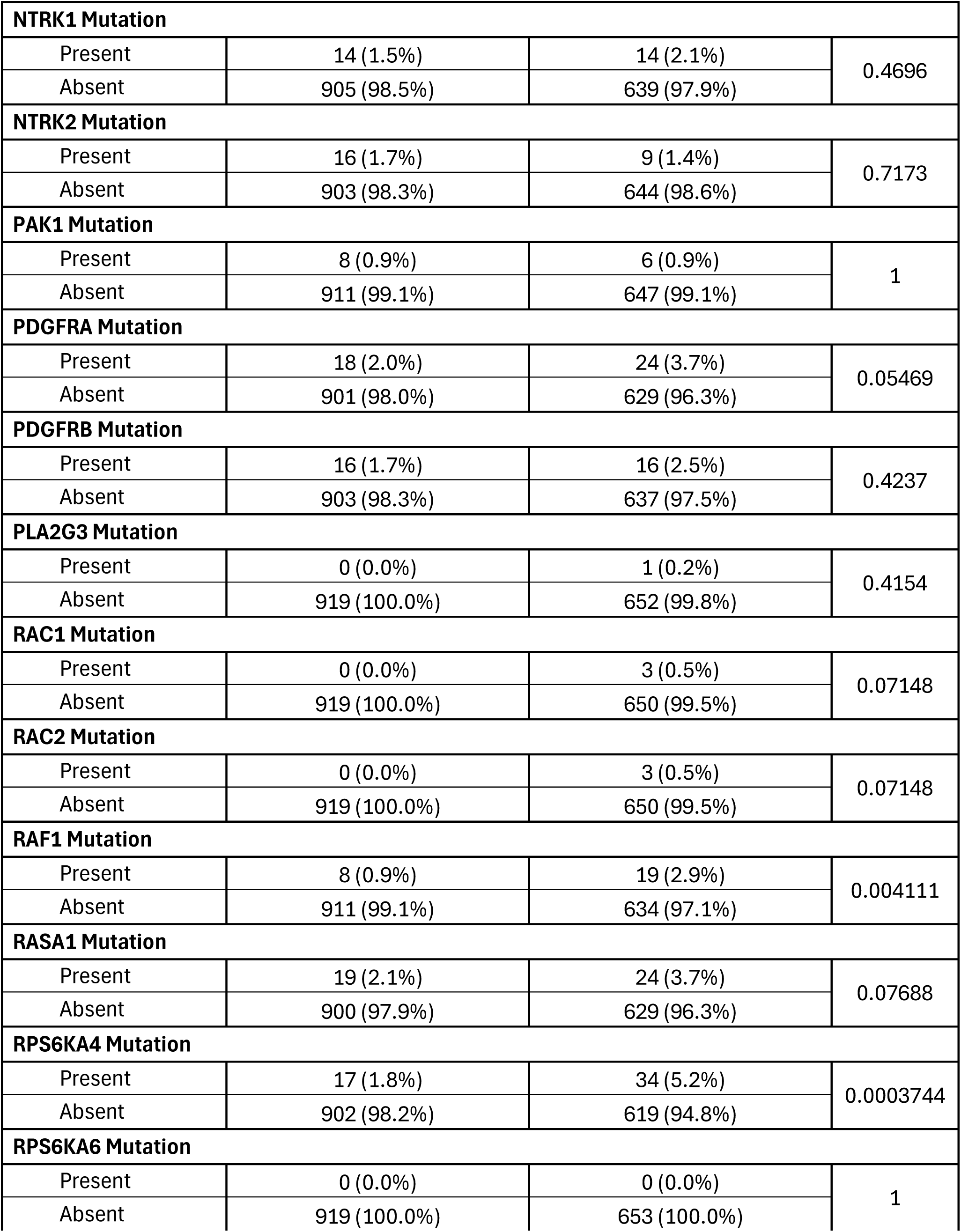
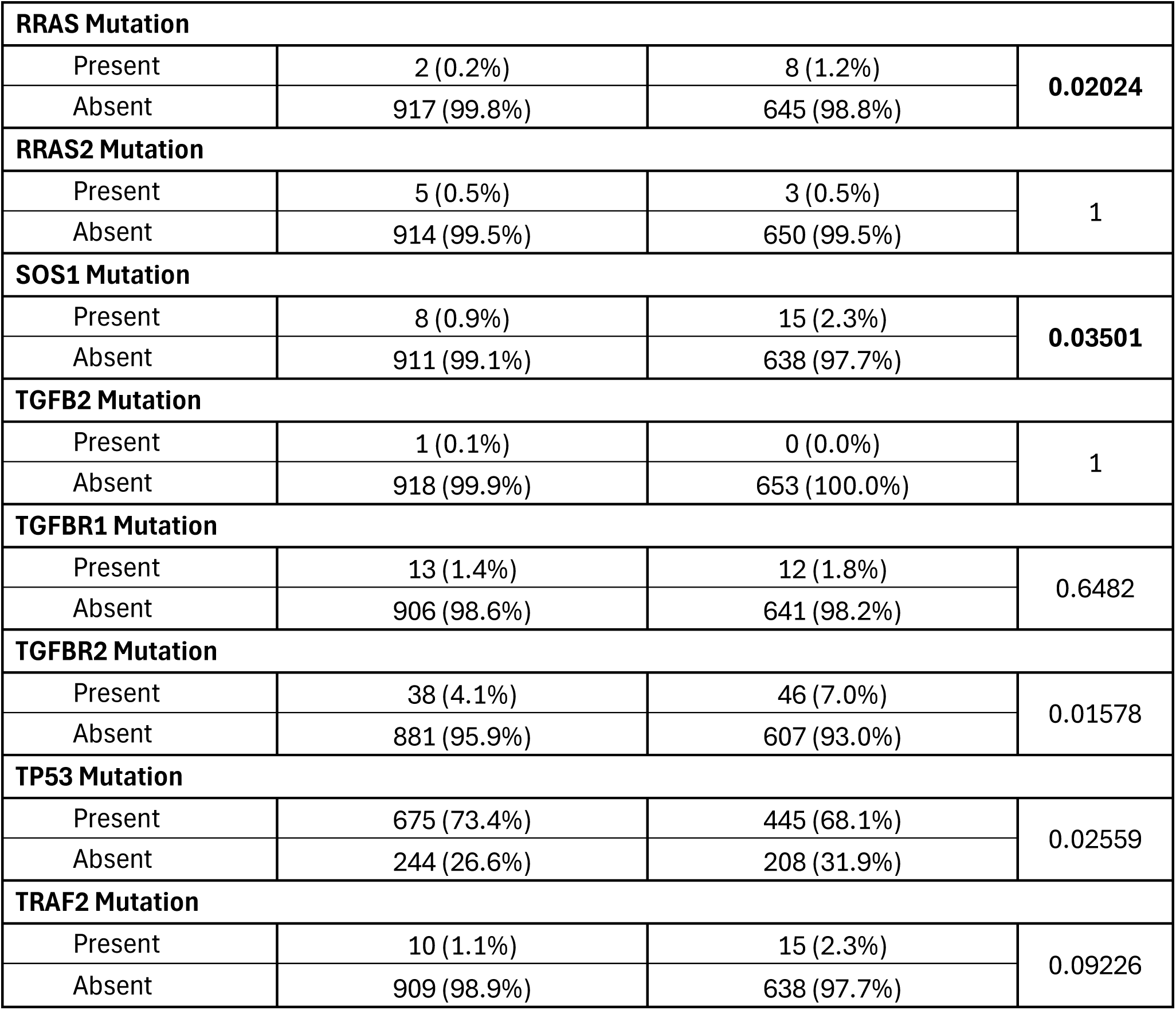
Comparison of Late-Onset Non-Hispanic White (NHW) Patients Treated with FOLFOX versus Not Treated with FOLFOX.

**Table S5.** Comparison of Early-Onset versus Late-Onset Hispanic/Latino (H/L) Patients Treated with FOLFOX.

| MAPK Pathway |  |  |  |
| --- | --- | --- | --- |
| Gene | Early-Onset Hispanic/Latino<br>Treated with FOLFOX<br>n (%) | Late-Onset Hispanic/Latino<br>Treated with FOLFOX<br>n (%) | p-value |
| ACVR1 Mutation |  |  |  |
| Present | 1 (1.4%) | 0 (0.0%) | 0.4451 |
| Absent | 72 (98.6%) | 91 (100.0%) |  |
| AKT1 Mutation |  |  |  |
| Present | 4 (5.5%) | 2 (2.2%) | 0.4082 |
| Absent | 69 (94.5%) | 89 (97.8%) |  |
| AKT2 Mutation |  |  |  |
| Present | 0 (0.0%) | 2 (2.2%) | 0.503 |
| Absent | 73 (100.0%) | 89 (97.8%) |  |
| AKT3 Mutation |  |  |  |
| Present | 2 (2.7%) | 2 (2.2%) | 1 |
| Absent | 71 (97.3%) | 89 (97.8%) |  |
| BRAF Mutation |  |  |  |
| Present | 3 (4.1%) | 15 (16.5%) | 0.01229 |
| Absent | 70 (95.9%) | 76 (83.5%) |  |
| CACNA1H Mutation |  |  |  |
| Present | 0 (0.0%) | 0 (0.0%) | 1 |
| Absent | 73 (100.0%) | 91 (100.0%) |  |
| CACNA2D1 Mutation |  |  |  |
| Present | 0 (0.0%) | 0 (0.0%) | 1 |
| Absent | 73 (100.0%) | 91 (100.0%) |  |
| CACNB1 Mutation |  |  |  |
| Present | 0 (0.0%) | 0 (0.0%) | 1 |
| Absent | 73 (100.0%) | 91 (100.0%) |  |
| CDC42 Mutation |  |  |  |
| Present | 0 (0.0%) | 1 (1.1%) | 1 |
| Absent | 73 (100.0%) | 90 (98.9%) |  |
| CRKL Mutation |  |  |  |
| Present | 0 (0.0%) | 0 (0.0%) | 1 |
| Absent | 73 (100.0%) | 91 (100.0%) |  |
| DAXX Mutation |  |  |  |
| Present | 1 (1.4%) | 0 (0.0%) | 0.4451 |
| Absent | 72 (98.6%) | 91 (100.0%) |  |
| DUSP4 Mutation |  |  |  |
| Present | 0 (0.0%) | 1 (1.1%) | 1 |
| Absent | 73 (100.0%) | 90 (98.9%) |  |
| EGFR Mutation |  |  |  |
| Present | 1 (1.4%) | 1 (1.1%) | 1 |
| Absent | 72 (98.6%) | 90 (98.9%) |  |
| FGF13 Mutation |  |  |  |
| Present | 0 (0.0%) | 1 (1.1%) | 1 |
| Absent | 73 (100.0%) | 90 (98.9%) |  |
| FGF19 Mutation |  |  |  |
| Present | 1 (1.4%) | 1 (1.1%) | 1 |
| Absent | 72 (98.6%) | 90 (98.9%) |  |
| FGF3 Mutation |  |  |  |
| Present | 0 (0.0%) | 2 (2.2%) | 0.503 |
| Absent | 73 (100.0%) | 89 (97.8%) |  |
| FGF4 Mutation |  |  |  |
| Present | 0 (0.0%) | 0 (0.0%) | 1 |
| Absent | 73 (100.0%) | 91 (100.0%) |  |
| FGFR1 Mutation |  |  |  |
| Present | 0 (0.0%) | 0 (0.0%) | 1 |
| Absent | 73 (100.0%) | 91 (100.0%) |  |
| FGFR2 Mutation |  |  |  |
| Present | 0 (0.0%) | 0 (0.0%) | 1 |
| Absent | 73 (100.0%) | 91 (100.0%) |  |
| FGFR3 Mutation |  |  |  |
| Present | 1 (1.4%) | 3 (3.3%) | 0.6296 |
| Absent | 72 (98.6%) | 88 (96.7%) |  |
| FGFR4 Mutation |  |  |  |
| Present | 1 (1.4%) | 1 (1.1%) | 1 |
| Absent | 72 (98.6%) | 90 (98.9%) |  |
| HRAS Mutation |  |  |  |
| Present | 1 (1.4%) | 0 (0.0%) | 0.4451 |
| Absent | 72 (98.6%) | 91 (100.0%) |  |
| JUN Mutation |  |  |  |
| Present | 3 (4.1%) | 0 (0.0%) | 0.08617 |
| Absent | 70 (95.9%) | 91 (100.0%) |  |
| KRAS Mutation |  |  |  |
| Present | 30 (41.1%) | 39 (42.9%) | 0.8742 |
| Absent | 43 (58.9%) | 52 (57.1%) |  |
| MAP2K1 Mutation |  |  |  |
| Present | 2 (2.7%) | 0 (0.0%) | 0.1966 |
| Absent | 71 (97.3%) | 91 (100.0%) |  |
| MAP2K2 Mutation |  |  |  |
| Present | 1 (1.4%) | 2 (2.2%) | 1 |
| Absent | 72 (98.6%) | 89 (97.8%) |  |
| MAP2K4 Mutation |  |  |  |
| Present | 3 (4.1%) | 1 (1.1%) | 0.3243 |
| Absent | 70 (95.9%) | 90 (98.9%) |  |
| MAP3K1 Mutation |  |  |  |
| Present | 2 (2.7%) | 1 (1.1%) | 0.5858 |
| Absent | 71 (97.3%) | 90 (98.9%) |  |
| MAP3K13 Mutation |  |  |  |
| Present | 2 (2.7%) | 2 (2.2%) | 1 |
| Absent | 71 (97.3%) | 89 (97.8%) |  |
| MAP3K14 Mutation |  |  |  |
| Present | 0 (0.0%) | 0 (0.0%) | 1 |
| Absent | 73 (100.0%) | 91 (100.0%) |  |
| MAPK1 Mutation |  |  |  |
| Present | 0 (0.0%) | 0 (0.0%) | 1 |
| Absent | 73 (100.0%) | 91 (100.0%) |  |
| MAPK3 Mutation |  |  |  |
| Present | 1 (1.4%) | 1 (1.1%) | 1 |
| Absent | 72 (98.6%) | 90 (98.9%) |  |
| MAX Mutation |  |  |  |
| Present | 0 (0.0%) | 0 (0.0%) | 1 |
| Absent | 73 (100.0%) | 91 (100.0%) |  |
| MYC Mutation |  |  |  |
| Present | 0 (0.0%) | 0 (0.0%) | 1 |
| Absent | 73 (100.0%) | 91 (100.0%) |  |
| NF1 Mutation |  |  |  |
| Present | 3 (4.1%) | 2 (2.2%) | 0.6566 |
| Absent | 70 (95.9%) | 89 (97.8%) |  |
| NFATC2 Mutation |  |  |  |
| Present | 0 (0.0%) | 0 (0.0%) | 1 |
| Absent | 73 (100.0%) | 91 (100.0%) |  |
| NRAS Mutation |  |  |  |
| Present | 2 (2.7%) | 6 (6.6%) | 0.3012 |
| Absent | 71 (97.3%) | 85 (93.4%) |  |
| NTRK1 Mutation |  |  |  |
| Present | 1 (1.4%) | 2 (2.2%) | 1 |
| Absent | 72 (98.6%) | 89 (97.8%) |  |
| NTRK2 Mutation |  |  |  |
| Present | 1 (1.4%) | 0 (0.0%) | 0.4451 |
| Absent | 72 (98.6%) | 91 (100.0%) |  |
| PAK1 Mutation |  |  |  |
| Present | 0 (0.0%) | 0 (0.0%) | 1 |
| Absent | 73 (100.0%) | 91 (100.0%) |  |
| PDGFRA Mutation |  |  |  |
| Present | 2 (2.7%) | 0 (0.0%) | 0.1966 |
| Absent | 71 (97.3%) | 91 (100.0%) |  |
| PDGFRB Mutation |  |  |  |
| Present | 4 (5.5%) | 0 (0.0%) | 0.03747 |
| Absent | 69 (94.5%) | 91 (100.0%) |  |
| PLA2G3 Mutation |  |  |  |
| Present | 0 (0.0%) | 0 (0.0%) | 1 |
| Absent | 73 (100.0%) | 91 (100.0%) |  |
| RAC1 Mutation |  |  |  |
| Present | 0 (0.0%) | 0 (0.0%) | 1 |
| Absent | 73 (100.0%) | 91 (100.0%) |  |
| RAC2 Mutation |  |  |  |
| Present | 0 (0.0%) | 0 (0.0%) | 1 |
| Absent | 73 (100.0%) | 91 (100.0%) |  |
| RAF1 Mutation |  |  |  |
| Present | 1 (1.4%) | 2 (2.2%) | 1 |
| Absent | 72 (98.6%) | 89 (97.8%) |  |
| RASA1 Mutation |  |  |  |
| Present | 1 (1.4%) | 4 (4.4%) | 0.3826 |
| Absent | 72 (98.6%) | 87 (95.6%) |  |
| RPS6KA4 Mutation |  |  |  |
| Present | 0 (0.0%) | 3 (3.3%) | 0.2545 |
| Absent | 73 (100.0%) | 88 (96.7%) |  |
| RPS6KA6 Mutation |  |  |  |
| Present | 0 (0.0%) | 0 (0.0%) | 1 |
| Absent | 73 (100.0%) | 91 (100.0%) |  |
| RRAS Mutation |  |  |  |
| Present | 0 (0.0%) | 0 (0.0%) | 1 |
| Absent | 73 (100.0%) | 91 (100.0%) |  |
| RRAS2 Mutation |  |  |  |
| Present | 0 (0.0%) | 1 (1.1%) | 1 |
| Absent | 73 (100.0%) | 90 (98.9%) |  |
| SOS1 Mutation |  |  |  |
| Present | 0 (0.0%) | 3 (3.3%) | 0.2545 |
| Absent | 73 (100.0%) | 88 (96.7%) |  |
| TGFB2 Mutation |  |  |  |
| Present | 0 (0.0%) | 0 (0.0%) | 1 |
| Absent | 73 (100.0%) | 91 (100.0%) |  |
| TGFBR1 Mutation |  |  |  |
| Present | 2 (2.7%) | 3 (3.3%) | 1 |
| Absent | 71 (97.3%) | 88 (96.7%) |  |
| TGFBR2 Mutation |  |  |  |
| Present | 3 (4.1%) | 5 (5.5%) | 0.7334 |
| Absent | 70 (95.9%) | 86 (94.5%) |  |
| TP53 Mutation |  |  |  |
| Present | 57 (78.1%) | 71 (78.0%) | 1 |
| Absent | 16 (21.9%) | 20 (22.0%) |  |
| TRAF2 Mutation |  |  |  |
| Present | 1 (1.4%) | 0 (0.0%) | 1 |
| Absent | 72 (98.6%) | 91 (100.0%) |  |

**Table S6.**
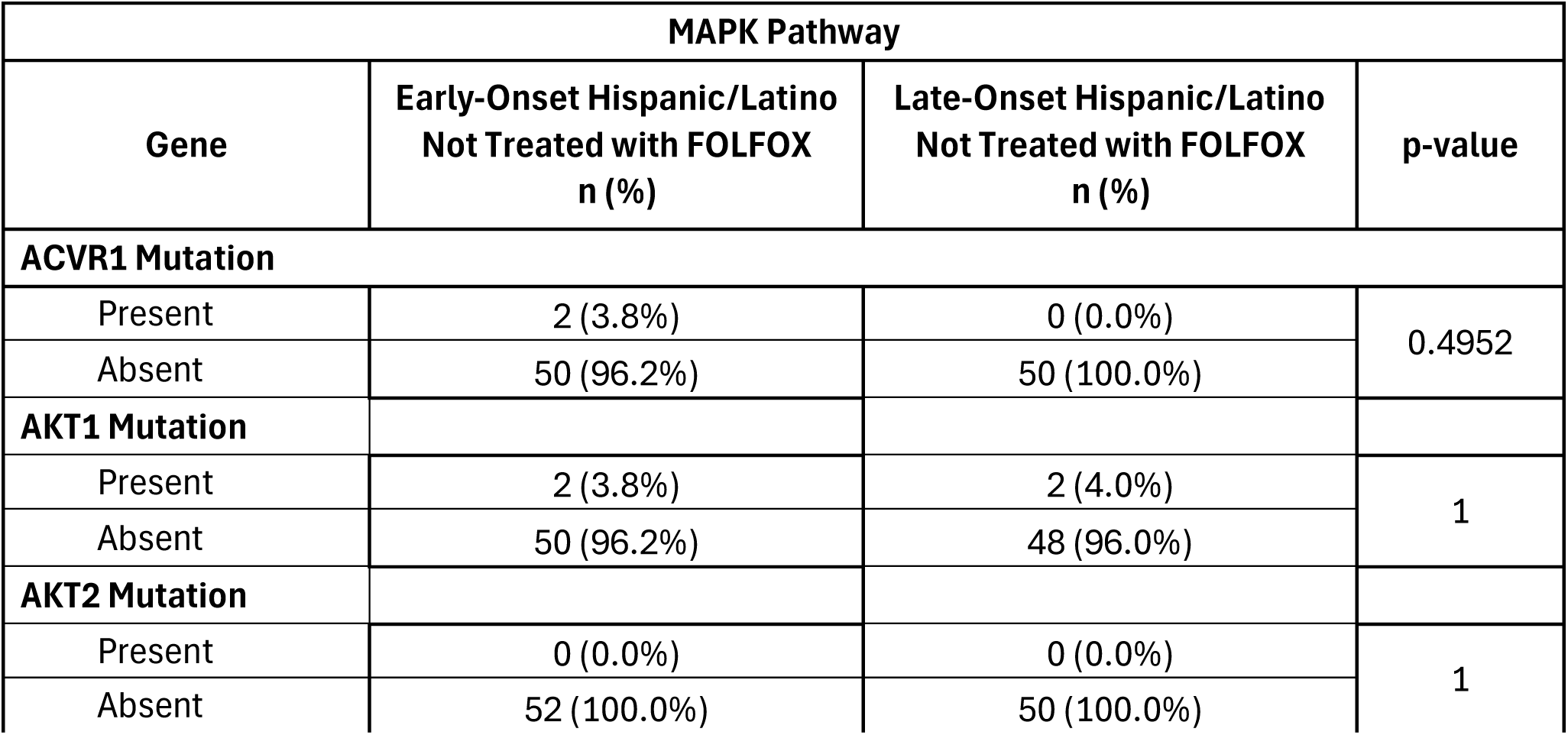
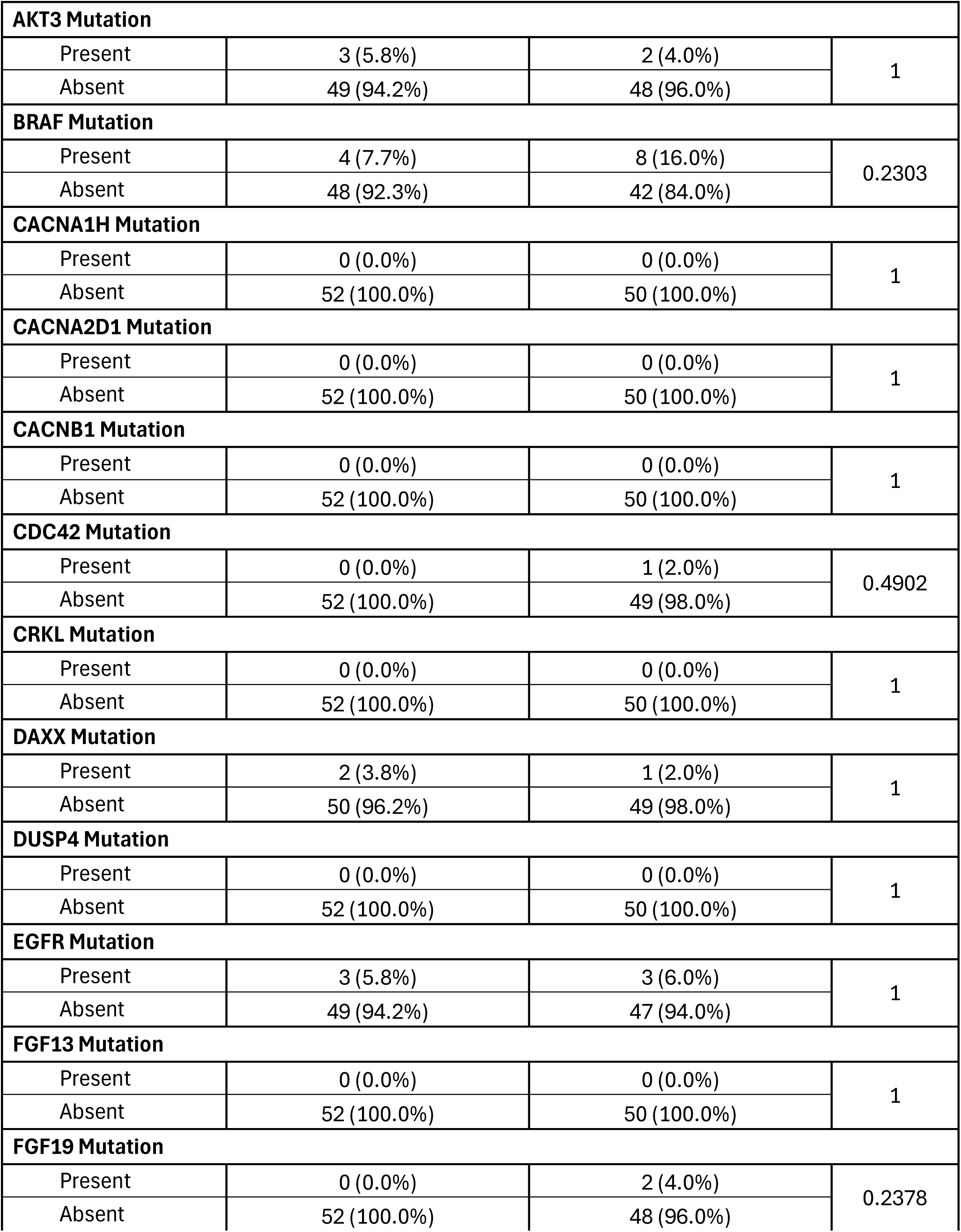
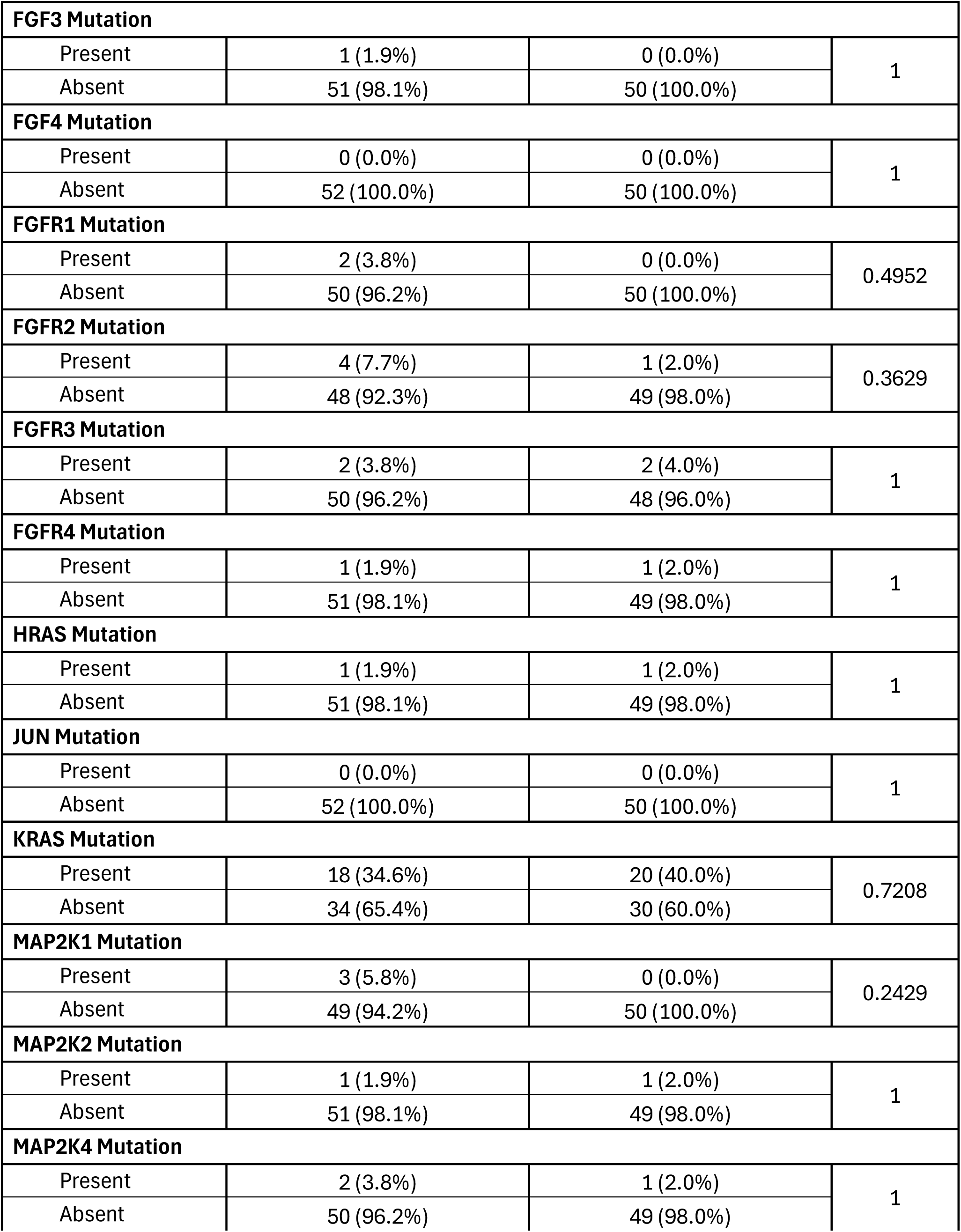
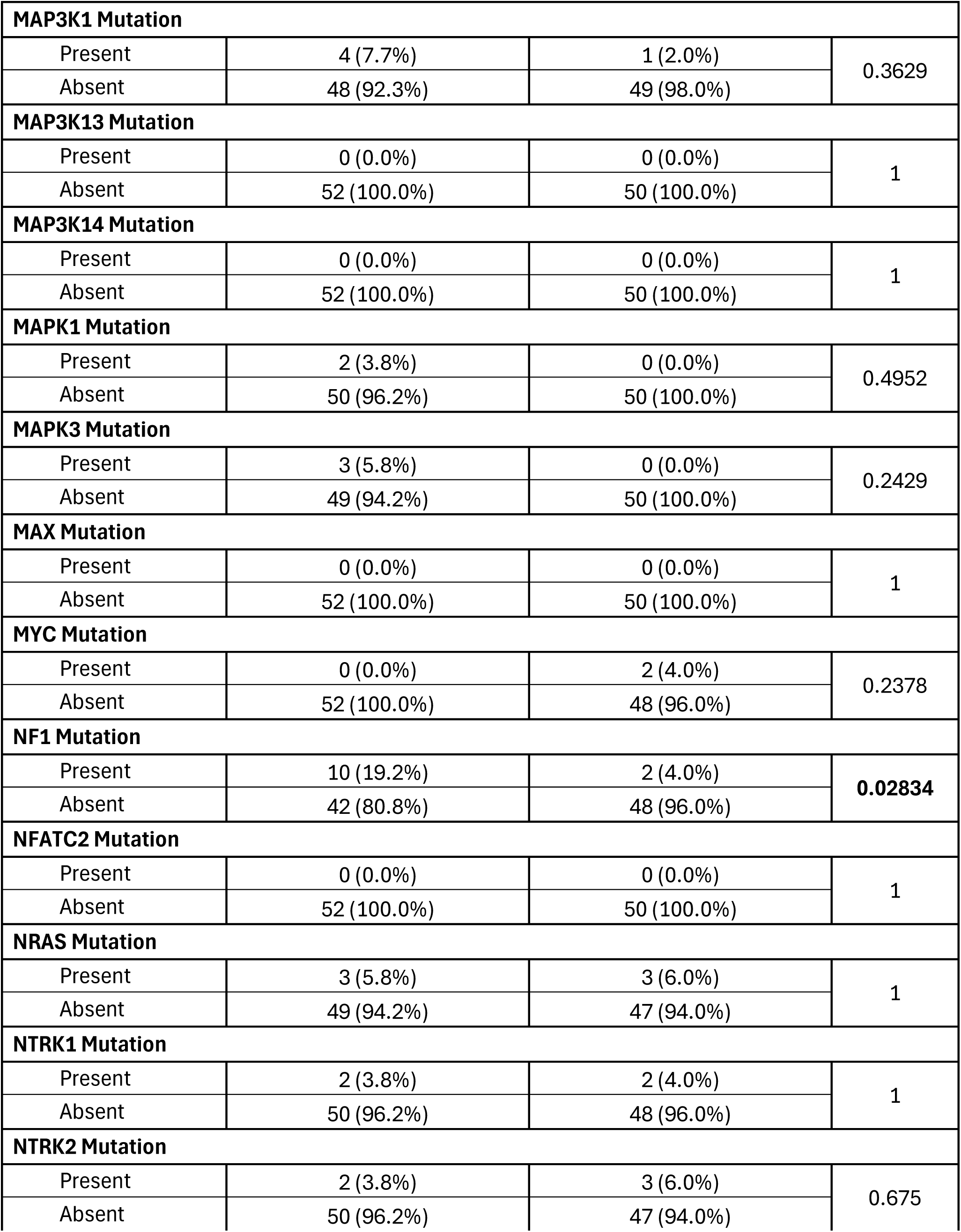
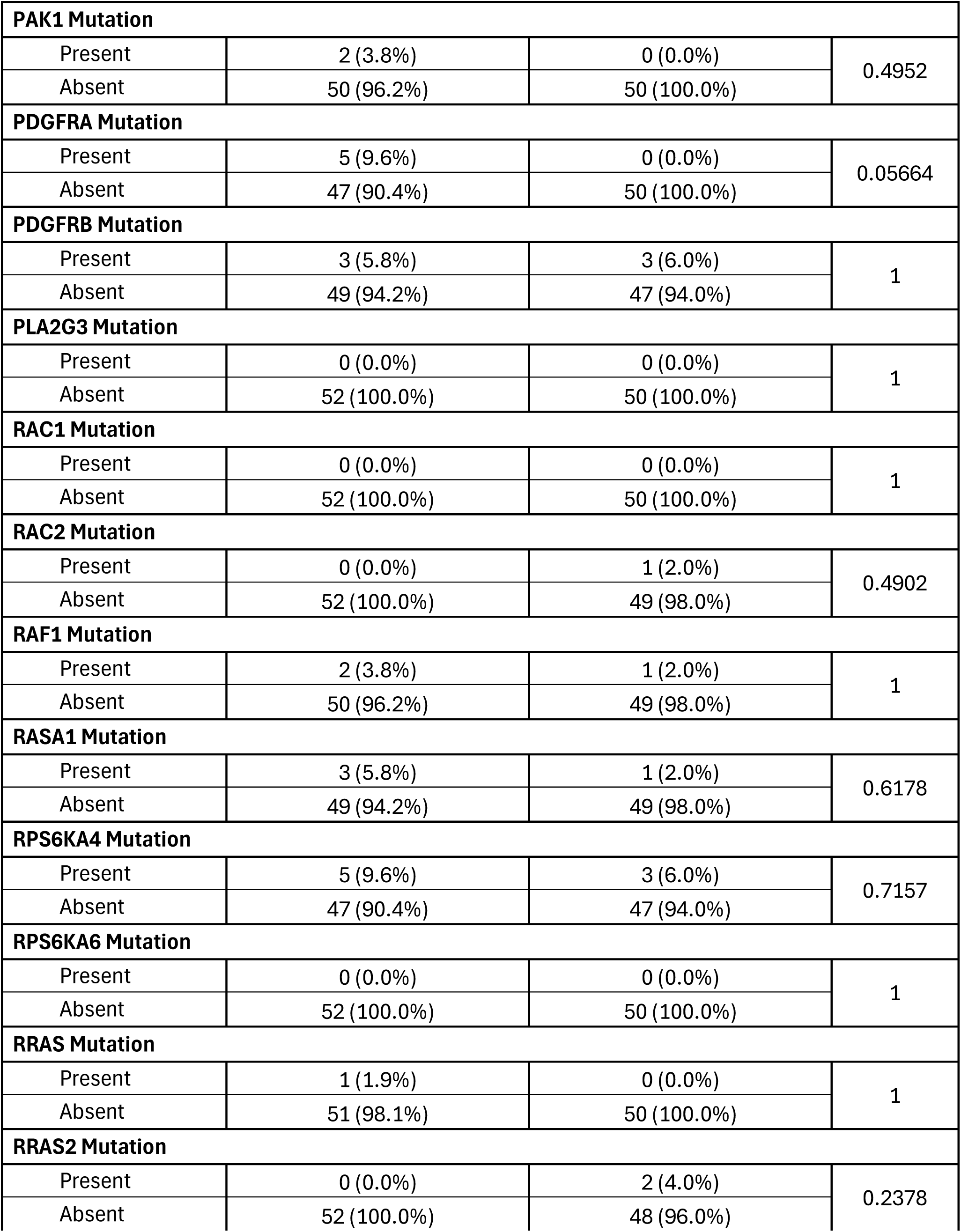
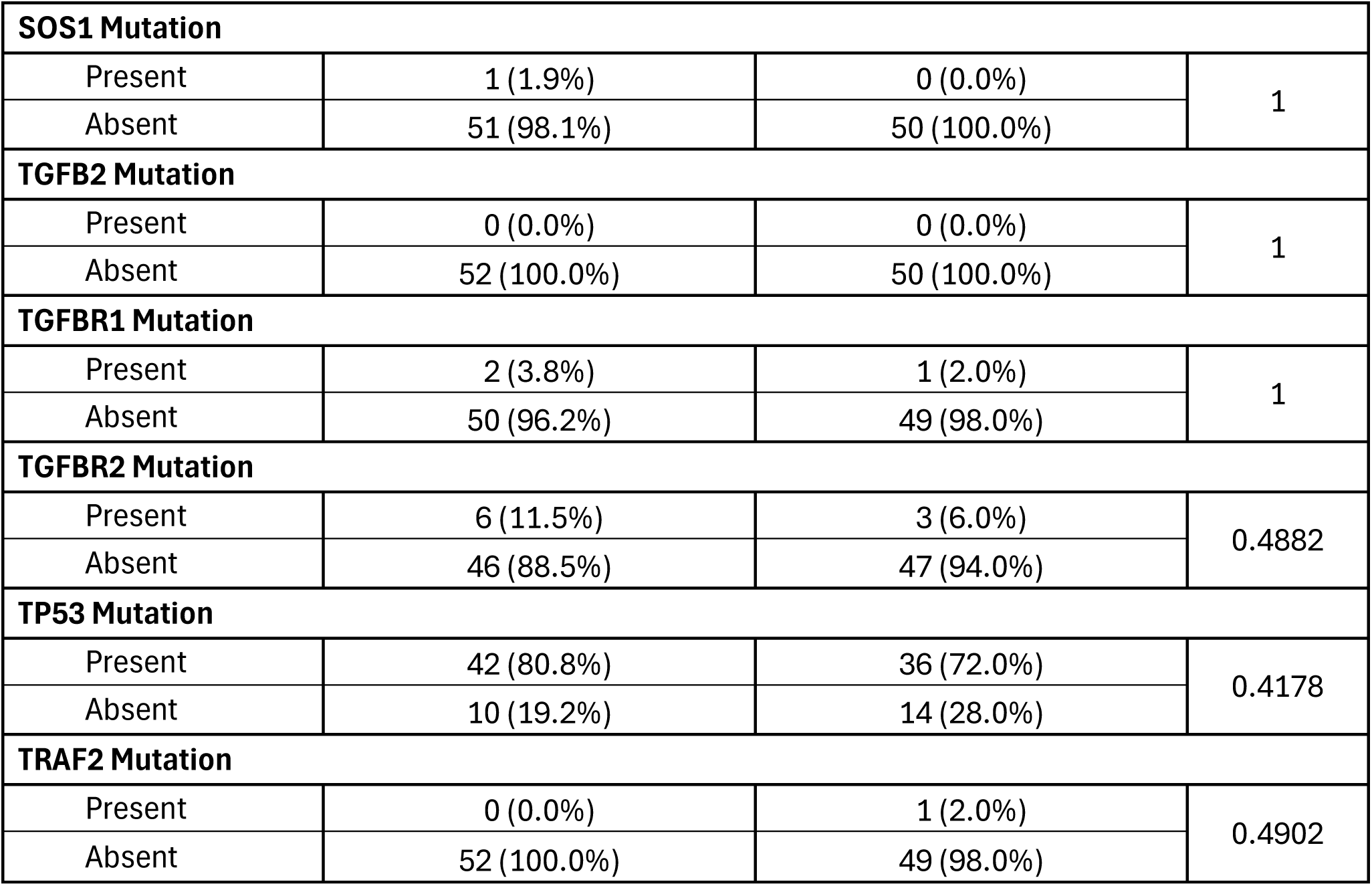
Comparison of Early-Onset versus Late-Onset Hispanic/Latino (H/L) Patients Not Treated with FOLFOX.

**Table S7.**
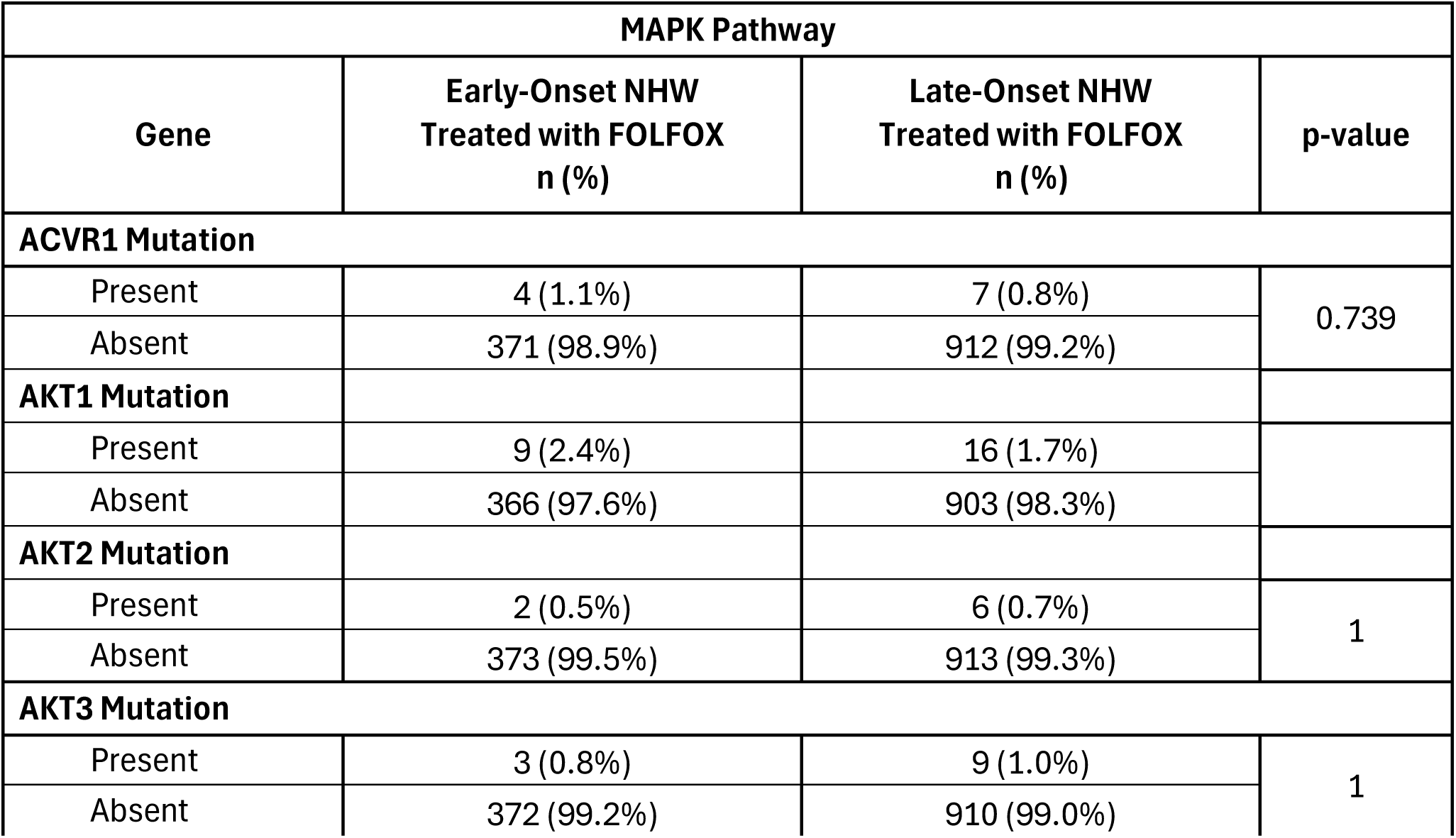
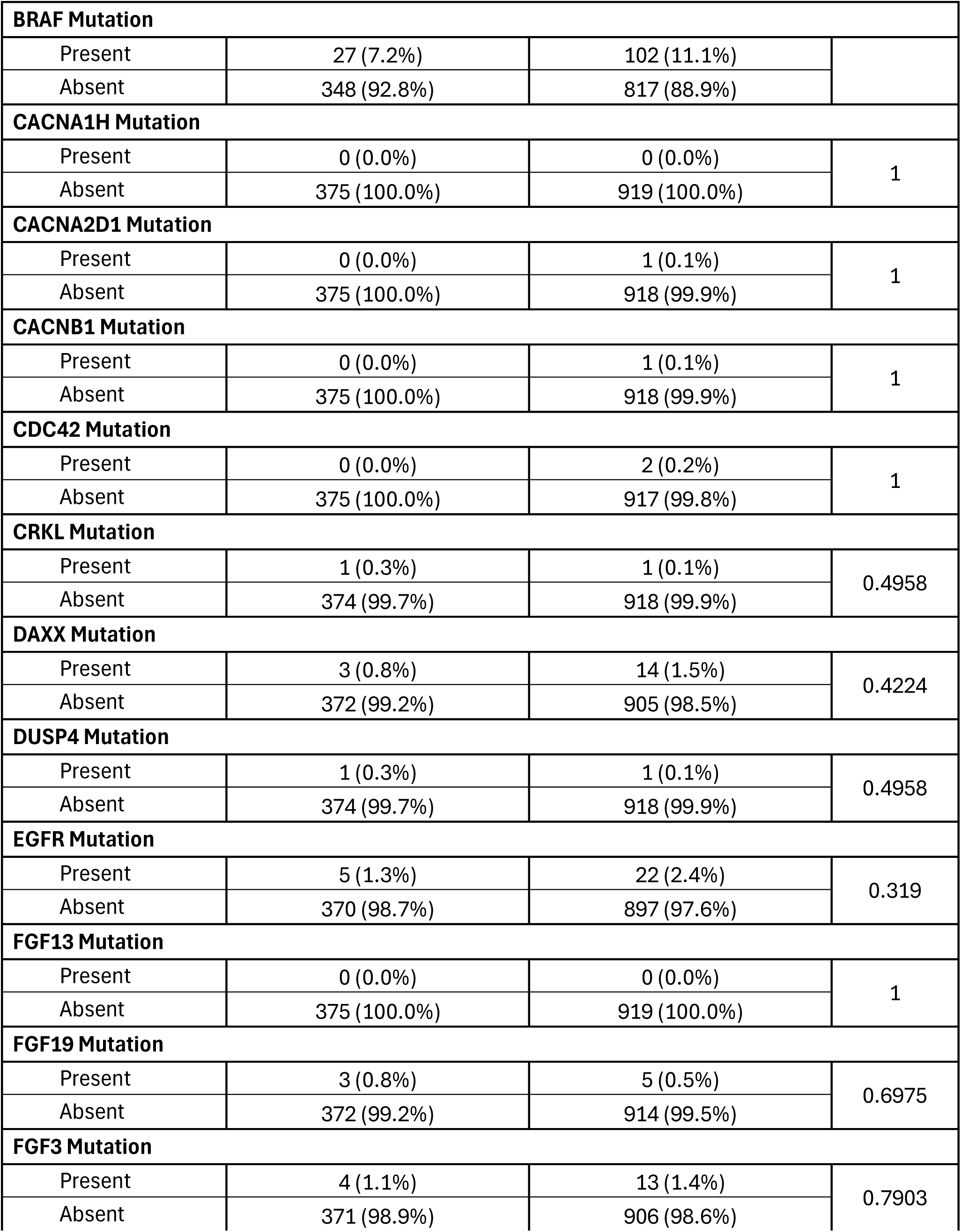
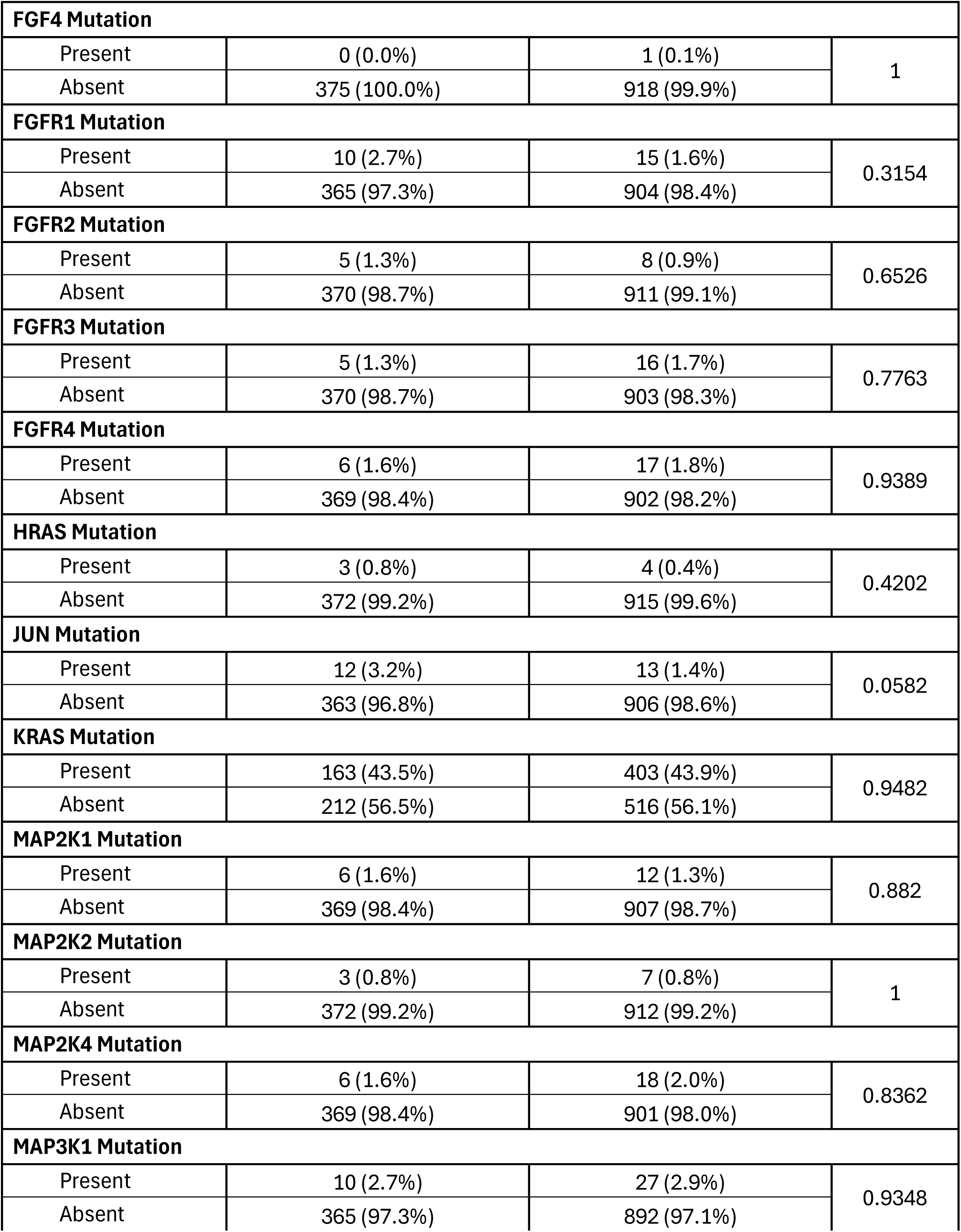
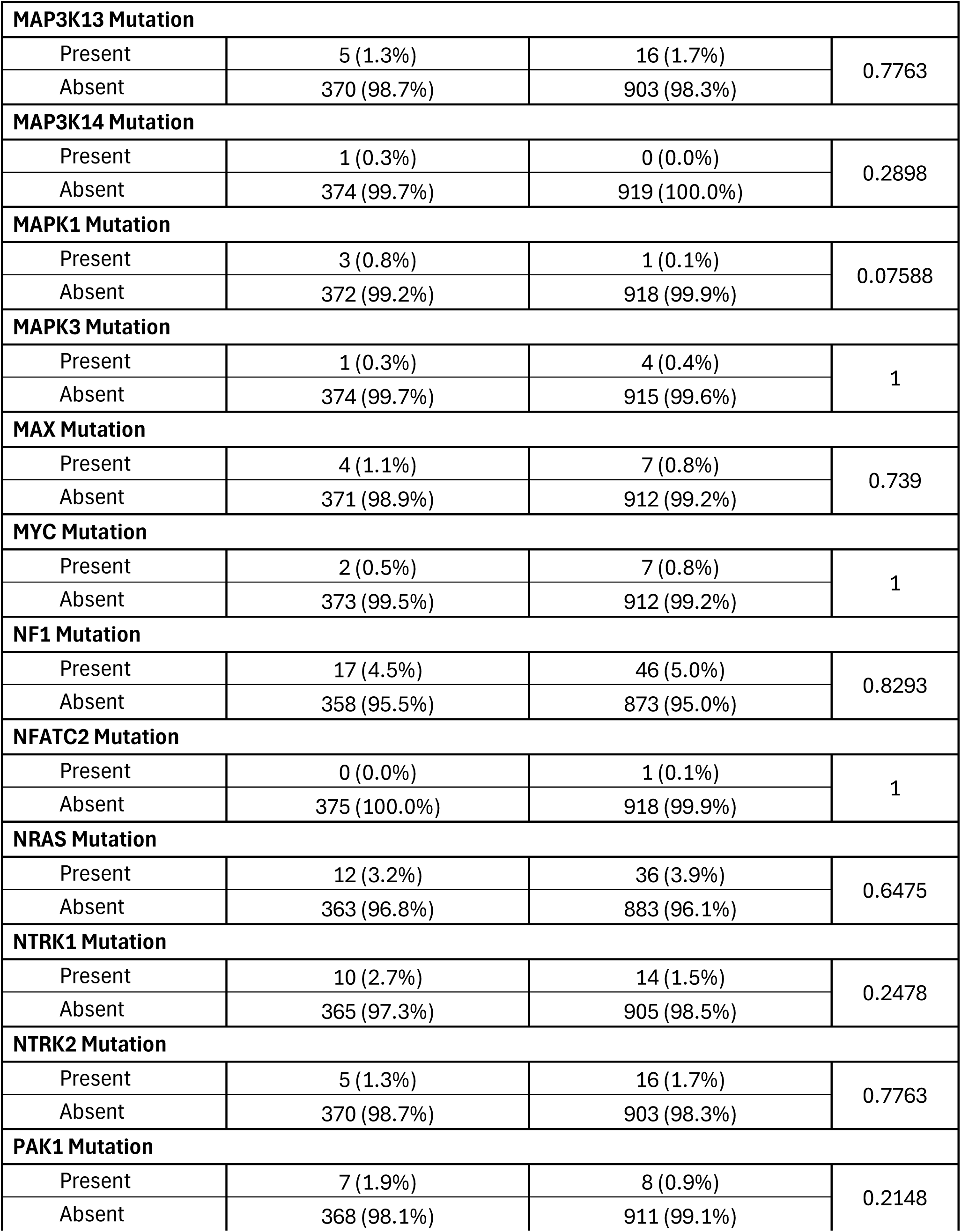
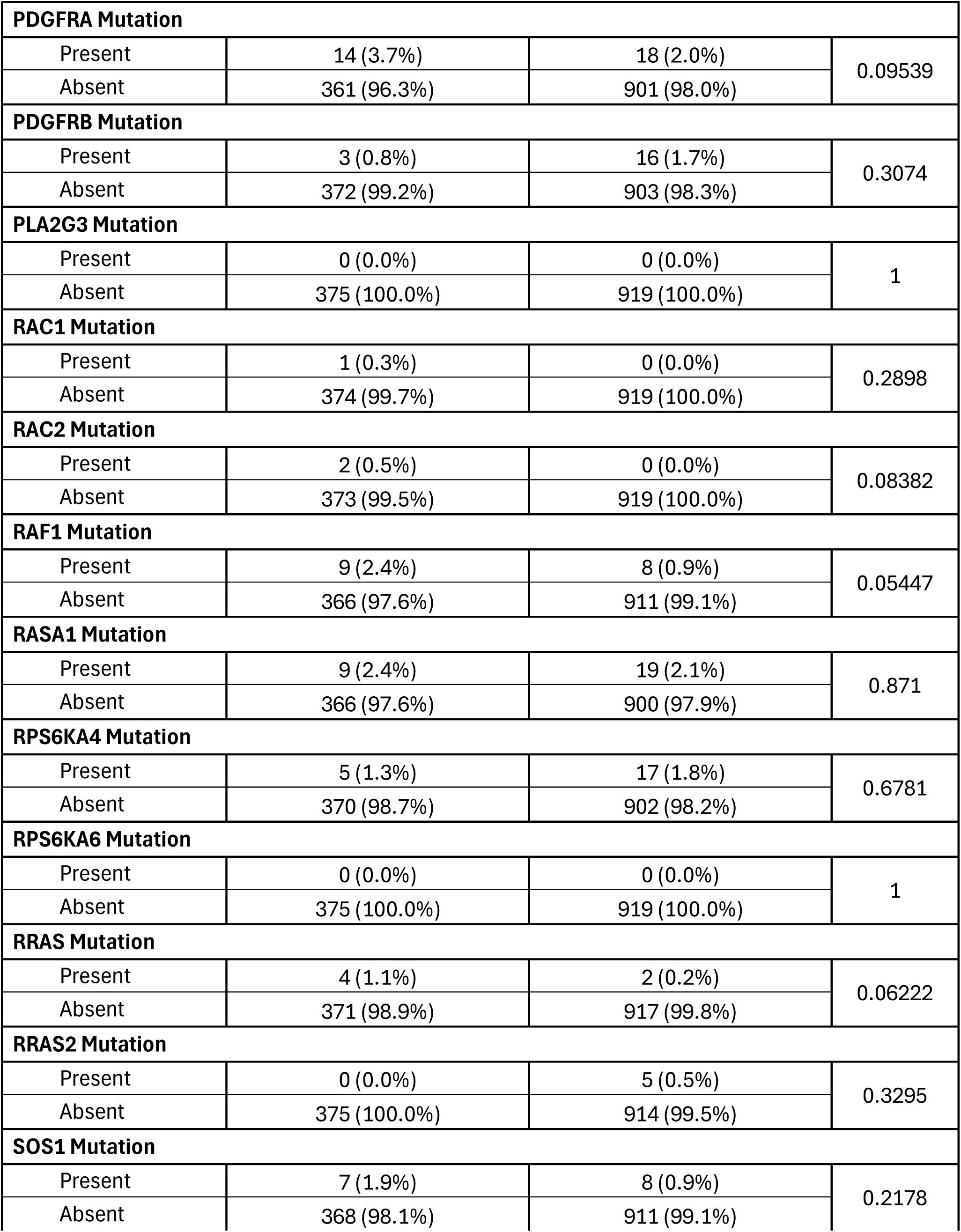
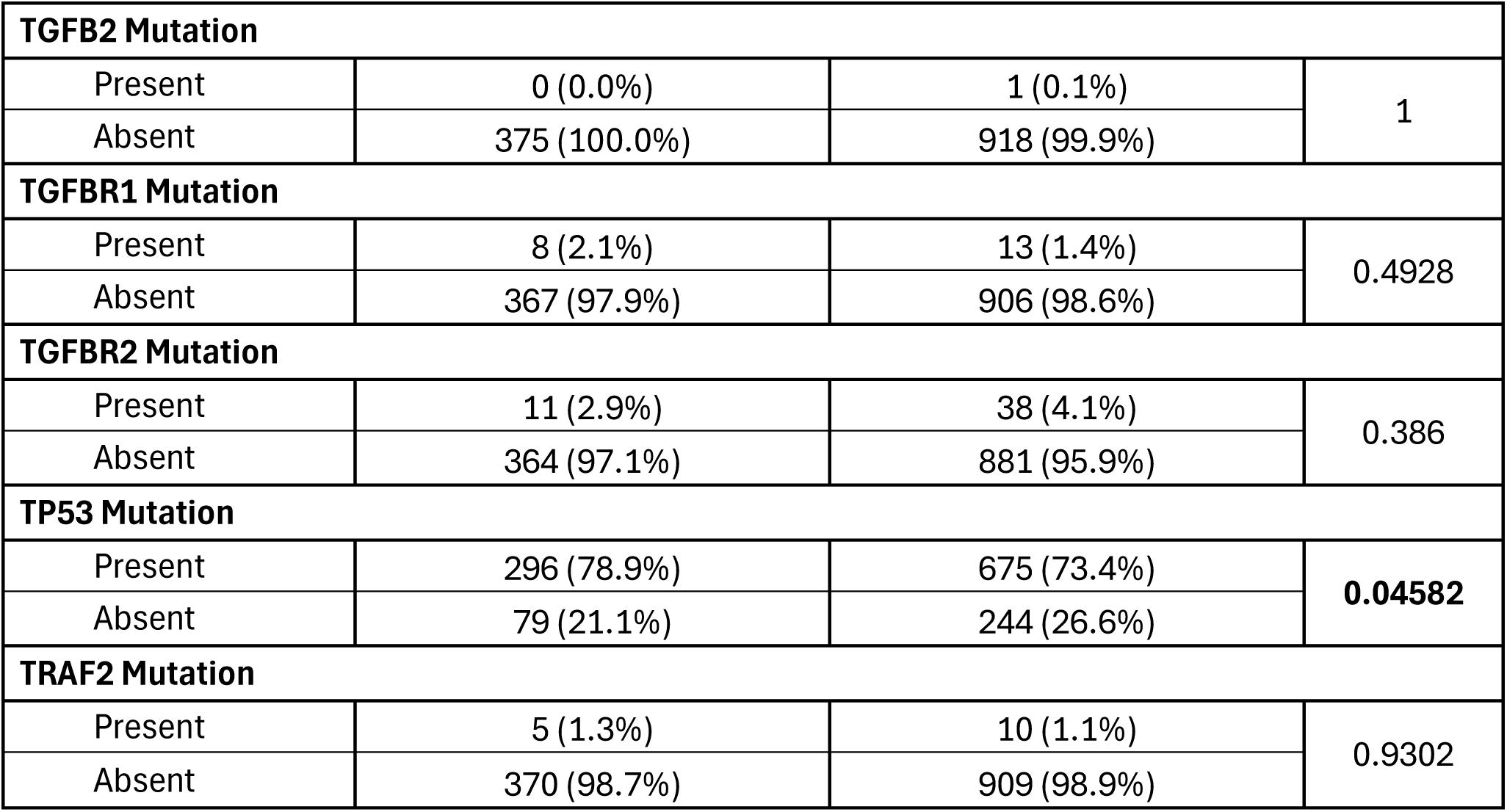
Comparison of Early-Onset versus Late-Onset Non-Hispanic White (NHW) Patients Treated with FOLFOX.

**Table S8.** Comparison of Early-Onset Hispanic/Latino (H/L) versus Early-Onset Non-Hispanic White (NHW) Patients Treated with FOLFOX.

| MAPK Pathway |  |  |  |
| --- | --- | --- | --- |
| Gene | Early-Onset NHW<br>Not Treated with FOLFOX<br>n (%) | Late-Onset NHW<br>Not Treated with FOLFOX<br>n (%) | p-value |
| ACVR1 Mutation |  |  |  |
| Present | 5 (1.7%) | 8 (1.2%) | 0.8153 |
| Absent | 297 (98.3%) | 645 (98.8%) |  |
| AKT1 Mutation |  |  |  |
| Present | 4 (1.3%) | 19 (2.9%) | 0.1748 |
| Absent | 298 (98.7%) | 634 (97.1%) |  |
| AKT2 Mutation |  |  |  |
| Present | 2 (0.7%) | 12 (1.8%) | 0.2466 |
| Absent | 300 (99.3%) | 641 (98.2%) |  |
| AKT3 Mutation |  |  |  |
| Present | 9 (3.0%) | 12 (1.8%) | 0.3777 |
| Absent | 293 (97.0%) | 641 (98.2%) |  |
| BRAF Mutation |  |  |  |
| Present | 24 (7.9%) | 93 (14.2%) | 0.007983 |
| Absent | 278 (92.1%) | 560 (85.8%) |  |
| CACNA1H Mutation |  |  |  |
| Present | 0 (0.0%) | 1 (0.2%) | 1 |
| Absent | 302 (100.0%) | 652 (99.8%) |  |
| CACNA2D1 Mutation |  |  |  |
| Present | 0 (0.0%) | 0 (0.0%) | 1 |
| Absent | 302 (100.0%) | 653 (100.0%) |  |
| CACNB1 Mutation |  |  |  |
| Present | 0 (0.0%) | 0 (0.0%) | 1 |
| Absent | 302 (100.0%) | 653 (100.0%) |  |
| CDC42 Mutation |  |  |  |
| Present | 2 (0.7%) | 2 (0.3%) | 0.5949 |
| Absent | 300 (99.3%) | 651 (99.7%) |  |
| CRKL Mutation |  |  |  |
| Present | 3 (1.0%) | 8 (1.2%) | 1 |
| Absent | 299 (99.0%) | 645 (98.8%) |  |
| DAXX Mutation |  |  |  |
| Present | 6 (2.0%) | 13 (2.0%) | 1 |
| Absent | 296 (98.0%) | 640 (98.0%) |  |
| DUSP4 Mutation |  |  |  |
| Present | 1 (0.3%) | 6 (0.9%) | 0.4424 |
| Absent | 301 (99.7%) | 647 (99.1%) |  |
| EGFR Mutation |  |  |  |
| Present | 5 (1.7%) | 24 (3.7%) | 0.1366 |
| Absent | 297 (98.3%) | 629 (96.3%) |  |
| FGF13 Mutation |  |  |  |
| Present | 0 (0.0%) | 0 (0.0%) | 1 |
| Absent | 302 (100.0%) | 653 (100.0%) |  |
| FGF19 Mutation |  |  |  |
| Present | 4 (1.3%) | 5 (0.8%) | 0.4751 |
| Absent | 298 (98.7%) | 648 (99.2%) |  |
| FGF3 Mutation |  |  |  |
| Present | 5 (1.7%) | 14 (2.1%) | 0.8 |
| Absent | 297 (98.3%) | 639 (97.9%) |  |
| FGF4 Mutation |  |  |  |
| Present | 5 (1.7%) | 4 (0.6%) | 0.151 |
| Absent | 297 (98.3%) | 649 (99.4%) |  |
| FGFR1 Mutation |  |  |  |
| Present | 6 (2.0%) | 16 (2.5%) | 0.8321 |
| Absent | 296 (98.0%) | 637 (97.5%) |  |
| FGFR2 Mutation |  |  |  |
| Present | 9 (3.0%) | 11 (1.7%) | 0.2904 |
| Absent | 293 (97.0%) | 642 (98.3%) |  |
| FGFR3 Mutation |  |  |  |
| Present | 11 (3.6%) | 19 (2.9%) | 0.6861 |
| Absent | 291 (96.4%) | 634 (97.1%) |  |
| FGFR4 Mutation |  |  |  |
| Present | 5 (1.7%) | 22 (3.4%) | 0.2021 |
| Absent | 297 (98.3%) | 631 (96.6%) |  |
| HRAS Mutation |  |  |  |
| Present | 2 (0.7%) | 6 (0.9%) | 1 |
| Absent | 300 (99.3%) | 647 (99.1%) |  |
| JUN Mutation |  |  |  |
| Present | 2 (0.7%) | 21 (3.2%) | 0.02065 |
| Absent | 300 (99.3%) | 632 (96.8%) |  |
| KRAS Mutation |  |  |  |
| Present | 124 (41.1%) | 279 (42.7%) | 0.6786 |
| Absent | 178 (58.9%) | 374 (57.3%) |  |
| MAP2K1 Mutation |  |  |  |
| Present | 4 (1.3%) | 10 (1.5%) | 1 |
| Absent | 298 (98.7%) | 643 (98.5%) |  |
| MAP2K2 Mutation |  |  |  |
| Present | 1 (0.3%) | 10 (1.5%) | 0.1885 |
| Absent | 301 (99.7%) | 643 (98.5%) |  |
| MAP2K4 Mutation |  |  |  |
| Present | 9 (3.0%) | 17 (2.6%) | 0.9054 |
| Absent | 293 (97.0%) | 636 (97.4%) |  |
| MAP3K1 Mutation |  |  |  |
| Present | 12 (4.0%) | 31 (4.7%) | 0.7125 |
| Absent | 290 (96.0%) | 622 (95.3%) |  |
| MAP3K13 Mutation |  |  |  |
| Present | 9 (3.0%) | 15 (2.3%) | 0.6856 |
| Absent | 293 (97.0%) | 638 (97.7%) |  |
| MAP3K14 Mutation |  |  |  |
| Present | 0 (0.0%) | 0 (0.0%) | 1 |
| Absent | 302 (100.0%) | 653 (100.0%) |  |
| MAPK1 Mutation |  |  |  |
| Present | 3 (1.0%) | 6 (0.9%) | 1 |
| Absent | 299 (99.0%) | 647 (99.1%) |  |
| MAPK3 Mutation |  |  |  |
| Present | 3 (1.0%) | 5 (0.8%) | 0.713 |
| Absent | 299 (99.0%) | 648 (99.2%) |  |
| MAX Mutation |  |  |  |
| Present | 1 (0.3%) | 5 (0.8%) | 0.671 |
| Absent | 301 (99.7%) | 648 (99.2%) |  |
| MYC Mutation |  |  |  |
| Present | 1 (0.3%) | 9 (1.4%) | 0.1837 |
| Absent | 301 (99.7%) | 644 (98.6%) |  |
| NF1 Mutation |  |  |  |
| Present | 18 (6.0%) | 44 (6.7%) | 0.7547 |
| Absent | 284 (94.0%) | 609 (93.3%) |  |
| NFATC2 Mutation |  |  |  |
| Present | 0 (0.0%) | 0 (0.0%) | 1 |
| Absent | 302 (100.0%) | 653 (100.0%) |  |
| NRAS Mutation |  |  |  |
| Present | 6 (2.0%) | 19 (2.9%) | 0.5401 |
| Absent | 296 (98.0%) | 634 (97.1%) |  |
| NTRK1 Mutation |  |  |  |
| Present | 9 (3.0%) | 14 (2.1%) | 0.5777 |
| Absent | 293 (97.0%) | 639 (97.9%) |  |
| NTRK2 Mutation |  |  |  |
| Present | 8 (2.6%) | 9 (1.4%) | 0.2636 |
| Absent | 294 (97.4%) | 644 (98.6%) |  |
| PAK1 Mutation |  |  |  |
| Present | 3 (1.0%) | 6 (0.9%) | 1 |
| Absent | 299 (99.0%) | 647 (99.1%) |  |
| PDGFRA Mutation |  |  |  |
| Present | 16 (5.3%) | 24 (3.7%) | 0.322 |
| Absent | 286 (94.7%) | 629 (96.3%) |  |
| PDGFRB Mutation |  |  |  |
| Present | 7 (2.3%) | 16 (2.5%) | 1 |
| Absent | 295 (97.7%) | 637 (97.5%) |  |
| PLA2G3 Mutation |  |  |  |
| Present | 0 (0.0%) | 1 (0.2%) | 1 |
| Absent | 302 (100.0%) | 652 (99.8%) |  |
| RAC1 Mutation |  |  |  |
| Present | 3 (1.0%) | 3 (0.5%) | 0.3873 |
| Absent | 299 (99.0%) | 650 (99.5%) |  |
| RAC2 Mutation |  |  |  |
| Present | 2 (0.7%) | 3 (0.5%) | 0.6539 |
| Absent | 300 (99.3%) | 650 (99.5%) |  |
| RAF1 Mutation |  |  |  |
| Present | 9 (3.0%) | 19 (2.9%) | 1 |
| Absent | 293 (97.0%) | 634 (97.1%) |  |
| RASA1 Mutation |  |  |  |
| Present | 14 (4.6%) | 24 (3.7%) | 0.5975 |
| Absent | 288 (95.4%) | 629 (96.3%) |  |
| RPS6KA4 Mutation |  |  |  |
| Present | 8 (2.6%) | 34 (5.2%) | 0.1046 |
| Absent | 294 (97.4%) | 619 (94.8%) |  |
| RPS6KA6 Mutation |  |  |  |
| Present | 1 (0.3%) | 0 (0.0%) | 0.3162 |
| Absent | 301 (99.7%) | 653 (100.0%) |  |
| RRAS Mutation |  |  |  |
| Present | 0 (0.0%) | 8 (1.2%) | 0.0617 |
| Absent | 302 (100.0%) | 645 (98.8%) |  |
| RRAS2 Mutation |  |  |  |
| Present | 4 (1.3%) | 3 (0.5%) | 0.2164 |
| Absent | 298 (98.7%) | 650 (99.5%) |  |
| SOS1 Mutation |  |  |  |
| Present | 11 (3.6%) | 15 (2.3%) | 0.33 |
| Absent | 291 (96.4%) | 638 (97.7%) |  |
| TGFB2 Mutation |  |  |  |
| Present | 0 (0.0%) | 0 (0.0%) | 1 |
| Absent | 302 (100.0%) | 653 (100.0%) |  |

| TGFBR1 Mutation |  |  |  |
| --- | --- | --- | --- |
| Present | 9 (3.0%) | 12 (1.8%) | 0.3777 |
| Absent | 293 (97.0%) | 641 (98.2%) |  |
| TGFBR2 Mutation |  |  |  |
| Present | 16 (5.3%) | 46 (7.0%) | 0.3803 |
| Absent | 286 (94.7%) | 607 (93.0%) |  |
| TP53 Mutation |  |  |  |
| Present | 228 (75.5%) | 445 (68.1%) | 0.02516 |
| Absent | 74 (24.5%) | 208 (31.9%) |  |
| TRAF2 Mutation |  |  |  |
| Present | 4 (1.3%) | 15 (2.3%) | 0.4556 |
| Absent | 298 (98.7%) | 638 (97.7%) |  |

**Table S9.** Comparison of Early-Onset Hispanic/Latino (H/L) versus Early-Onset Non-Hispanic White (NHW) Patients Not Treated with FOLFOX.

| MAPK Pathway |  |  |  |
| --- | --- | --- | --- |
| Gene | Early-Onset Hispanic/Latino<br>Treated with FOLFOX<br>n (%) | Early-Onset NHW<br>Treated with FOLFOX<br>n (%) | p-value |
| ACVR1 Mutation |  |  |  |
| Present | 1 (1.4%) | 4 (1.1%) | 0.5909 |
| Absent | 72 (98.6%) | 371 (98.9%) |  |
| AKT1 Mutation |  |  |  |
| Present | 4 (5.5%) | 9 (2.4%) | 0.2414 |
| Absent | 69 (94.5%) | 366 (97.6%) |  |
| AKT2 Mutation |  |  |  |
| Present | 0 (0.0%) | 2 (0.5%) | 1 |
| Absent | 73 (100.0%) | 373 (99.5%) |  |
| AKT3 Mutation |  |  |  |
| Present | 2 (2.7%) | 3 (0.8%) | 0.1883 |
| Absent | 71 (97.3%) | 372 (99.2%) |  |
| BRAF Mutation |  |  |  |
| Present | 3 (4.1%) | 27 (7.2%) | 0.4472 |
| Absent | 70 (95.9%) | 348 (92.8%) |  |
| CACNA1H Mutation |  |  |  |
| Present | 0 (0.0%) | 0 (0.0%) | 1 |
| Absent | 73 (100.0%) | 375 (100.0%) |  |
| CACNA2D1 Mutation |  |  |  |
| Present | 0 (0.0%) | 0 (0.0%) | 1 |
| Absent | 73 (100.0%) | 375 (100.0%) |  |
| CACNB1 Mutation |  |  |  |
| Present | 0 (0.0%) | 0 (0.0%) | 1 |
| Absent | 73 (100.0%) | 375 (100.0%) |  |
| CDC42 Mutation |  |  |  |
| Present | 0 (0.0%) | 0 (0.0%) | 1 |
| Absent | 73 (100.0%) | 375 (100.0%) |  |
| CRKL Mutation |  |  |  |
| Present | 0 (0.0%) | 1 (0.3%) | 1 |
| Absent | 73 (100.0%) | 374 (99.7%) |  |
| DAXX Mutation |  |  |  |
| Present | 1 (1.4%) | 3 (0.8%) | 0.5104 |
| Absent | 72 (98.6%) | 372 (99.2%) |  |
| DUSP4 Mutation |  |  |  |
| Present | 0 (0.0%) | 1 (0.3%) | 1 |
| Absent | 73 (100.0%) | 374 (99.7%) |  |
| EGFR Mutation |  |  |  |
| Present | 1 (1.4%) | 5 (1.3%) | 1 |
| Absent | 72 (98.6%) | 370 (98.7%) |  |
| FGF13 Mutation |  |  |  |
| Present | 0 (0.0%) | 0 (0.0%) | 1 |
| Absent | 73 (100.0%) | 375 (100.0%) |  |
| FGF19 Mutation |  |  |  |
| Present | 1 (1.4%) | 3 (0.8%) | 0.5104 |
| Absent | 72 (98.6%) | 372 (99.2%) |  |
| FGF3 Mutation |  |  |  |
| Present | 0 (0.0%) | 4 (1.1%) | 1 |
| Absent | 73 (100.0%) | 371 (98.9%) |  |
| FGF4 Mutation |  |  |  |
| Present | 0 (0.0%) | 0 (0.0%) | 1 |
| Absent | 73 (100.0%) | 375 (100.0%) |  |
| FGFR1 Mutation |  |  |  |
| Present | 0 (0.0%) | 10 (2.7%) | 0.3783 |
| Absent | 73 (100.0%) | 365 (97.3%) |  |
| FGFR2 Mutation |  |  |  |
| Present | 0 (0.0%) | 5 (1.3%) | 1 |
| Absent | 73 (100.0%) | 370 (98.7%) |  |
| FGFR3 Mutation |  |  |  |
| Present | 1 (1.4%) | 5 (1.3%) | 1 |
| Absent | 72 (98.6%) | 370 (98.7%) |  |
| FGFR4 Mutation |  |  |  |
| Present | 1 (1.4%) | 6 (1.6%) | 1 |
| Absent | 72 (98.6%) | 369 (98.4%) |  |
| HRAS Mutation |  |  |  |
| Present | 1 (1.4%) | 3 (0.8%) | 0.5104 |
| Absent | 72 (98.6%) | 372 (99.2%) |  |
| JUN Mutation |  |  |  |
| Present | 3 (4.1%) | 12 (3.2%) | 0.7203 |
| Absent | 70 (95.9%) | 363 (96.8%) |  |
| KRAS Mutation |  |  |  |
| Present | 30 (41.1%) | 163 (43.5%) | 0.8064 |
| Absent | 43 (58.9%) | 212 (56.5%) |  |
| MAP2K1 Mutation |  |  |  |
| Present | 2 (2.7%) | 6 (1.6%) | 0.6222 |
| Absent | 71 (97.3%) | 369 (98.4%) |  |
| MAP2K2 Mutation |  |  |  |
| Present | 1 (1.4%) | 3 (0.8%) | 0.5104 |
| Absent | 72 (98.6%) | 372 (99.2%) |  |
| MAP2K4 Mutation |  |  |  |
| Present | 3 (4.1%) | 6 (1.6%) | 0.1678 |
| Absent | 70 (95.9%) | 369 (98.4%) |  |
| MAP3K1 Mutation |  |  |  |
| Present | 2 (2.7%) | 10 (2.7%) | 1 |
| Absent | 71 (97.3%) | 365 (97.3%) |  |
| MAP3K13 Mutation |  |  |  |
| Present | 2 (2.7%) | 5 (1.3%) | 0.3197 |
| Absent | 71 (97.3%) | 370 (98.7%) |  |
| MAP3K14 Mutation |  |  |  |
| Present | 0 (0.0%) | 1 (0.3%) | 1 |
| Absent | 73 (100.0%) | 374 (99.7%) |  |
| MAPK1 Mutation |  |  |  |
| Present | 0 (0.0%) | 3 (0.8%) | 1 |
| Absent | 73 (100.0%) | 372 (99.2%) |  |
| MAPK3 Mutation |  |  |  |
| Present | 1 (1.4%) | 1 (0.3%) | 0.2996 |
| Absent | 72 (98.6%) | 374 (99.7%) |  |
| MAX Mutation |  |  |  |
| Present | 0 (0.0%) | 4 (1.1%) | 1 |
| Absent | 73 (100.0%) | 371 (98.9%) |  |
| MYC Mutation |  |  |  |
| Present | 0 (0.0%) | 2 (0.5%) | 1 |
| Absent | 73 (100.0%) | 373 (99.5%) |  |
| NF1 Mutation |  |  |  |
| Present | 3 (4.1%) | 17 (4.5%) | 1 |
| Absent | 70 (95.9%) | 358 (95.5%) |  |
| NFATC2 Mutation |  |  |  |
| Present | 0 (0.0%) | 0 (0.0%) | 1 |
| Absent | 73 (100.0%) | 375 (100.0%) |  |
| NRAS Mutation |  |  |  |
| Present | 2 (2.7%) | 12 (3.2%) | 1 |
| Absent | 71 (97.3%) | 363 (96.8%) |  |
| NTRK1 Mutation |  |  |  |
| Present | 1 (1.4%) | 10 (2.7%) | 1 |
| Absent | 72 (98.6%) | 365 (97.3%) |  |
| NTRK2 Mutation |  |  |  |
| Present | 1 (1.4%) | 5 (1.3%) | 1 |
| Absent | 72 (98.6%) | 370 (98.7%) |  |
| PAK1 Mutation |  |  |  |
| Present | 0 (0.0%) | 7 (1.9%) | 0.6049 |
| Absent | 73 (100.0%) | 368 (98.1%) |  |
| PDGFRA Mutation |  |  |  |
| Present | 2 (2.7%) | 14 (3.7%) | 1 |
| Absent | 71 (97.3%) | 361 (96.3%) |  |
| PDGFRB Mutation |  |  |  |
| Present | 4 (5.5%) | 3 (0.8%) | 0.01547 |
| Absent | 69 (94.5%) | 372 (99.2%) |  |
| PLA2G3 Mutation |  |  |  |
| Present | 0 (0.0%) | 0 (0.0%) | 1 |
| Absent | 73 (100.0%) | 375 (100.0%) |  |
| RAC1 Mutation |  |  |  |
| Present | 0 (0.0%) | 1 (0.3%) | 1 |
| Absent | 73 (100.0%) | 374 (99.7%) |  |
| RAC2 Mutation |  |  |  |
| Present | 0 (0.0%) | 2 (0.5%) | 1 |
| Absent | 73 (100.0%) | 373 (99.5%) |  |
| RAF1 Mutation |  |  |  |
| Present | 1 (1.4%) | 9 (2.4%) | 1 |
| Absent | 72 (98.6%) | 366 (97.6%) |  |
| RASA1 Mutation |  |  |  |
| Present | 1 (1.4%) | 9 (2.4%) | 1 |
| Absent | 72 (98.6%) | 366 (97.6%) |  |
| RPS6KA4 Mutation |  |  |  |
| Present | 0 (0.0%) | 5 (1.3%) | 1 |
| Absent | 73 (100.0%) | 370 (98.7%) |  |
| RPS6KA6 Mutation |  |  |  |
| Present | 0 (0.0%) | 0 (0.0%) | 1 |
| Absent | 73 (100.0%) | 375 (100.0%) |  |
| RRAS Mutation |  |  |  |
| Present | 0 (0.0%) | 4 (1.1%) | 1 |
| Absent | 73 (100.0%) | 371 (98.9%) |  |
| RRAS2 Mutation |  |  |  |
| Present | 0 (0.0%) | 0 (0.0%) | 1 |
| Absent | 73 (100.0%) | 375 (100.0%) |  |
| SOS1 Mutation |  |  |  |
| Present | 0 (0.0%) | 7 (1.9%) | 0.6049 |
| Absent | 73 (100.0%) | 368 (98.1%) |  |
| TGFB2 Mutation |  |  |  |
| Present | 0 (0.0%) | 0 (0.0%) | 1 |
| Absent | 73 (100.0%) | 375 (100.0%) |  |
| TGFBR1 Mutation |  |  |  |
| Present | 2 (2.7%) | 8 (2.1%) | 0.6698 |
| Absent | 71 (97.3%) | 367 (97.9%) |  |

| TGFBR2 Mutation |  |  |  |
| --- | --- | --- | --- |
| Present | 3 (4.1%) | 11 (2.9%) | 0.7102 |
| Absent | 70 (95.9%) | 364 (97.1%) |  |
| TP53 Mutation |  |  |  |
| Present | 57 (78.1%) | 296 (78.9%) | 0.995 |
| Absent | 16 (21.9%) | 79 (21.1%) |  |
| TRAF2 Mutation |  |  |  |
| Present | 1 (1.4%) | 5 (1.3%) | 1 |
| Absent | 72 (98.6%) | 370 (98.7%) |  |

**Table S10.** Comparison of Late-Onset Hispanic/Latino (H/L) versus Late-Onset Non-Hispanic White (NHW) Patients Treated with FOLFOX.

| MAPK Pathway |  |  |  |
| --- | --- | --- | --- |
| Gene | Early-Onset Hispanic/Latino<br>Not Treated with FOLFOX<br>n (%) | Early-Onset NHW<br>Not Treated with FOLFOX<br>n (%) | p-value |
| ACVR1 Mutation |  |  |  |
| Present | 2 (3.8%) | 5 (1.7%) | 0.2743 |
| Absent | 50 (96.2%) | 297 (98.3%) |  |
| AKT1 Mutation |  |  |  |
| Present | 2 (3.8%) | 4 (1.3%) | 0.2153 |
| Absent | 50 (96.2%) | 298 (98.7%) |  |
| AKT2 Mutation |  |  |  |
| Present | 0 (0.0%) | 2 (0.7%) | 1 |
| Absent | 52 (100.0%) | 300 (99.3%) |  |
| AKT3 Mutation |  |  |  |
| Present | 3 (5.8%) | 9 (3.0%) | 0.3954 |
| Absent | 49 (94.2%) | 293 (97.0%) |  |
| BRAF Mutation |  |  |  |
| Present | 4 (7.7%) | 24 (7.9%) | 1 |
| Absent | 48 (92.3%) | 278 (92.1%) |  |
| CACNA1H Mutation |  |  |  |
| Present | 0 (0.0%) | 0 (0.0%) | 1 |
| Absent | 52 (100.0%) | 302 (100.0%) |  |
| CACNA2D1 Mutation |  |  |  |
| Present | 0 (0.0%) | 0 (0.0%) | 1 |
| Absent | 52 (100.0%) | 302 (100.0%) |  |
| CACNB1 Mutation |  |  |  |
| Present | 0 (0.0%) | 0 (0.0%) | 1 |
| Absent | 52 (100.0%) | 302 (100.0%) |  |
| CDC42 Mutation |  |  |  |
| Present | 0 (0.0%) | 2 (0.7%) | 1 |
| Absent | 52 (100.0%) | 300 (99.3%) |  |
| CRKL Mutation |  |  |  |
| Present | 0 (0.0%) | 3 (1.0%) | 1 |
| Absent | 52 (100.0%) | 299 (99.0%) |  |
| DAXX Mutation |  |  |  |
| Present | 2 (3.8%) | 6 (2.0%) | 0.3331 |
| Absent | 50 (96.2%) | 296 (98.0%) |  |
| DUSP4 Mutation |  |  |  |
| Present | 0 (0.0%) | 1 (0.3%) | 1 |
| Absent | 52 (100.0%) | 301 (99.7%) |  |
| EGFR Mutation |  |  |  |
| Present | 3 (5.8%) | 5 (1.7%) | 0.09778 |
| Absent | 49 (94.2%) | 297 (98.3%) |  |
| FGF13 Mutation |  |  |  |
| Present | 0 (0.0%) | 0 (0.0%) | 1 |
| Absent | 52 (100.0%) | 302 (100.0%) |  |
| FGF19 Mutation |  |  |  |
| Present | 0 (0.0%) | 4 (1.3%) | 1 |
| Absent | 52 (100.0%) | 298 (98.7%) |  |
| FGF3 Mutation |  |  |  |
| Present | 1 (1.9%) | 5 (1.7%) | 1 |
| Absent | 51 (98.1%) | 297 (98.3%) |  |
| FGF4 Mutation |  |  |  |
| Present | 0 (0.0%) | 5 (1.7%) | 1 |
| Absent | 52 (100.0%) | 297 (98.3%) |  |
| FGFR1 Mutation |  |  |  |
| Present | 2 (3.8%) | 6 (2.0%) | 0.3331 |
| Absent | 50 (96.2%) | 296 (98.0%) |  |
| FGFR2 Mutation |  |  |  |
| Present | 4 (7.7%) | 9 (3.0%) | 0.1073 |
| Absent | 48 (92.3%) | 293 (97.0%) |  |
| FGFR3 Mutation |  |  |  |
| Present | 2 (3.8%) | 11 (3.6%) | 1 |
| Absent | 50 (96.2%) | 291 (96.4%) |  |
| FGFR4 Mutation |  |  |  |
| Present | 1 (1.9%) | 5 (1.7%) | 1 |
| Absent | 51 (98.1%) | 297 (98.3%) |  |
| HRAS Mutation |  |  |  |
| Present | 1 (1.9%) | 2 (0.7%) | 0.38 |
| Absent | 51 (98.1%) | 300 (99.3%) |  |
| JUN Mutation |  |  |  |
| Present | 0 (0.0%) | 2 (0.7%) | 1 |
| Absent | 52 (100.0%) | 300 (99.3%) |  |
| KRAS Mutation |  |  |  |
| Present | 18 (34.6%) | 124 (41.1%) | 0.47 |
| Absent | 34 (65.4%) | 178 (58.9%) |  |
| MAP2K1 Mutation |  |  |  |
| Present | 3 (5.8%) | 4 (1.3%) | 0.06807 |
| Absent | 49 (94.2%) | 298 (98.7%) |  |
| MAP2K2 Mutation |  |  |  |
| Present | 1 (1.9%) | 1 (0.3%) | 0.2726 |
| Absent | 51 (98.1%) | 301 (99.7%) |  |
| MAP2K4 Mutation |  |  |  |
| Present | 2 (3.8%) | 9 (3.0%) | 0.668 |
| Absent | 50 (96.2%) | 293 (97.0%) |  |
| MAP3K1 Mutation |  |  |  |
| Present | 4 (7.7%) | 12 (4.0%) | 0.2694 |
| Absent | 48 (92.3%) | 290 (96.0%) |  |
| MAP3K13 Mutation |  |  |  |
| Present | 0 (0.0%) | 9 (3.0%) | 0.3669 |
| Absent | 52 (100.0%) | 293 (97.0%) |  |
| MAP3K14 Mutation |  |  |  |
| Present | 0 (0.0%) | 0 (0.0%) | 1 |
| Absent | 52 (100.0%) | 302 (100.0%) |  |
| MAPK1 Mutation |  |  |  |
| Present | 2 (3.8%) | 3 (1.0%) | 0.158 |
| Absent | 50 (96.2%) | 299 (99.0%) |  |
| MAPK3 Mutation |  |  |  |
| Present | 3 (5.8%) | 3 (1.0%) | 0.04335 |
| Absent | 49 (94.2%) | 299 (99.0%) |  |
| MAX Mutation |  |  |  |
| Present | 0 (0.0%) | 1 (0.3%) | 1 |
| Absent | 52 (100.0%) | 301 (99.7%) |  |
| MYC Mutation |  |  |  |
| Present | 0 (0.0%) | 1 (0.3%) | 1 |
| Absent | 52 (100.0%) | 301 (99.7%) |  |
| NF1 Mutation |  |  |  |
| Present | 10 (19.2%) | 18 (6.0%) | 0.002728 |
| Absent | 42 (80.8%) | 284 (94.0%) |  |
| NFATC2 Mutation |  |  |  |
| Present | 0 (0.0%) | 0 (0.0%) | 1 |
| Absent | 52 (100.0%) | 302 (100.0%) |  |
| NRAS Mutation |  |  |  |
| Present | 3 (5.8%) | 6 (2.0%) | 0.1318 |
| Absent | 49 (94.2%) | 296 (98.0%) |  |
| NTRK1 Mutation |  |  |  |
| Present | 2 (3.8%) | 9 (3.0%) | 0.668 |
| Absent | 50 (96.2%) | 293 (97.0%) |  |
| NTRK2 Mutation |  |  |  |
| Present | 2 (3.8%) | 8 (2.6%) | 0.6456 |
| Absent | 50 (96.2%) | 294 (97.4%) |  |
| PAK1 Mutation |  |  |  |
| Present | 2 (3.8%) | 3 (1.0%) | 0.158 |
| Absent | 50 (96.2%) | 299 (99.0%) |  |
| PDGFRA Mutation |  |  |  |
| Present | 5 (9.6%) | 16 (5.3%) | 0.3684 |
| Absent | 47 (90.4%) | 286 (94.7%) |  |
| PDGFRB Mutation |  |  |  |
| Present | 3 (5.8%) | 7 (2.3%) | 0.1693 |
| Absent | 49 (94.2%) | 295 (97.7%) |  |
| PLA2G3 Mutation |  |  |  |
| Present | 0 (0.0%) | 0 (0.0%) | 1 |
| Absent | 52 (100.0%) | 302 (100.0%) |  |
| RAC1 Mutation |  |  |  |
| Present | 0 (0.0%) | 3 (1.0%) | 1 |
| Absent | 52 (100.0%) | 299 (99.0%) |  |
| RAC2 Mutation |  |  |  |
| Present | 0 (0.0%) | 2 (0.7%) | 1 |
| Absent | 52 (100.0%) | 300 (99.3%) |  |
| RAF1 Mutation |  |  |  |
| Present | 2 (3.8%) | 9 (3.0%) | 0.668 |
| Absent | 50 (96.2%) | 293 (97.0%) |  |
| RASA1 Mutation |  |  |  |
| Present | 3 (5.8%) | 14 (4.6%) | 0.7245 |
| Absent | 49 (94.2%) | 288 (95.4%) |  |
| RPS6KA4 Mutation |  |  |  |
| Present | 5 (9.6%) | 8 (2.6%) | 0.03866 |
| Absent | 47 (90.4%) | 294 (97.4%) |  |
| RPS6KA6 Mutation |  |  |  |
| Present | 0 (0.0%) | 1 (0.3%) | 1 |
| Absent | 52 (100.0%) | 301 (99.7%) |  |
| RRAS Mutation |  |  |  |
| Present | 1 (1.9%) | 0 (0.0%) | 0.1469 |
| Absent | 51 (98.1%) | 302 (100.0%) |  |
| RRAS2 Mutation |  |  |  |
| Present | 0 (0.0%) | 4 (1.3%) | 1 |
| Absent | 52 (100.0%) | 298 (98.7%) |  |
| SOS1 Mutation |  |  |  |
| Present | 1 (1.9%) | 11 (3.6%) | 1 |
| Absent | 51 (98.1%) | 291 (96.4%) |  |
| TGFB2 Mutation |  |  |  |
| Present | 0 (0.0%) | 0 (0.0%) | 1 |
| Absent | 52 (100.0%) | 302 (100.0%) |  |
| TGFBR1 Mutation |  |  |  |
| Present | 2 (3.8%) | 9 (3.0%) | 0.668 |
| Absent | 50 (96.2%) | 293 (97.0%) |  |
| TGFBR2 Mutation |  |  |  |
| Present | 6 (11.5%) | 16 (5.3%) | 0.1583 |
| Absent | 46 (88.5%) | 286 (94.7%) |  |

| TP53 Mutation |  |  |  |
| --- | --- | --- | --- |
| Present | 42 (80.8%) | 228 (75.5%) | 0.5163 |
| Absent | 10 (19.2%) | 74 (24.5%) |  |
| TRAF2 Mutation |  |  |  |
| Present | 0 (0.0%) | 4 (1.3%) | 1 |
| Absent | 52 (100.0%) | 298 (98.7%) |  |

**Table S11.** Comparison of Late-Onset Hispanic/Latino (H/L) versus Late-Onset Non-Hispanic White (NHW) Patients Not Treated with FOLFOX.

| MAPK Pathway |  |  |  |
| --- | --- | --- | --- |
| Gene | Late-Onset Hispanic/Latino<br>Treated with FOLFOX<br>n (%) | Late-Onset NHW<br>Treated with FOLFOX<br>n (%) | p-value |
| ACVR1 Mutation |  |  |  |
| Present | 0 (0.0%) | 7 (0.8%) | 1 |
| Absent | 91 (100.0%) | 912 (99.2%) |  |
| AKT1 Mutation |  |  |  |
| Present | 2 (2.2%) | 16 (1.7%) | 0.6731 |
| Absent | 89 (97.8%) | 903 (98.3%) |  |
| AKT2 Mutation |  |  |  |
| Present | 2 (2.2%) | 6 (0.7%) | 0.1574 |
| Absent | 89 (97.8%) | 913 (99.3%) |  |
| AKT3 Mutation |  |  |  |
| Present | 2 (2.2%) | 9 (1.0%) | 0.2603 |
| Absent | 89 (97.8%) | 910 (99.0%) |  |
| BRAF Mutation |  |  |  |
| Present | 15 (16.5%) | 102 (11.1%) | 0.1741 |
| Absent | 76 (83.5%) | 817 (88.9%) |  |
| CACNA1H Mutation |  |  |  |
| Present | 0 (0.0%) | 0 (0.0%) | 1 |
| Absent | 91 (100.0%) | 919 (100.0%) |  |
| CACNA2D1 Mutation |  |  |  |
| Present | 0 (0.0%) | 1 (0.1%) | 1 |
| Absent | 91 (100.0%) | 918 (99.9%) |  |
| CACNB1 Mutation |  |  |  |
| Present | 0 (0.0%) | 1 (0.1%) | 1 |
| Absent | 91 (100.0%) | 918 (99.9%) |  |
| CDC42 Mutation |  |  |  |
| Present | 1 (1.1%) | 2 (0.2%) | 0.2469 |
| Absent | 90 (98.9%) | 917 (99.8%) |  |
| CRKL Mutation |  |  |  |
| Present | 0 (0.0%) | 1 (0.1%) | 1 |
| Absent | 91 (100.0%) | 918 (99.9%) |  |
| DAXX Mutation |  |  |  |
| Present | 0 (0.0%) | 14 (1.5%) | 0.6284 |
| Absent | 91 (100.0%) | 905 (98.5%) |  |
| DUSP4 Mutation |  |  |  |
| Present | 1 (1.1%) | 1 (0.1%) | 0.1722 |
| Absent | 90 (98.9%) | 918 (99.9%) |  |
| EGFR Mutation |  |  |  |
| Present | 1 (1.1%) | 22 (2.4%) | 0.7141 |
| Absent | 90 (98.9%) | 897 (97.6%) |  |
| FGF13 Mutation |  |  |  |
| Present | 1 (1.1%) | 0 (0.0%) | 0.0901 |
| Absent | 90 (98.9%) | 919 (100.0%) |  |
| FGF19 Mutation |  |  |  |
| Present | 1 (1.1%) | 5 (0.5%) | 0.4333 |
| Absent | 90 (98.9%) | 914 (99.5%) |  |
| FGF3 Mutation |  |  |  |
| Present | 2 (2.2%) | 13 (1.4%) | 0.6379 |
| Absent | 89 (97.8%) | 906 (98.6%) |  |
| FGF4 Mutation |  |  |  |
| Present | 0 (0.0%) | 1 (0.1%) | 1 |
| Absent | 91 (100.0%) | 918 (99.9%) |  |
| FGFR1 Mutation |  |  |  |
| Present | 0 (0.0%) | 15 (1.6%) | 0.386 |
| Absent | 91 (100.0%) | 904 (98.4%) |  |
| FGFR2 Mutation |  |  |  |
| Present | 0 (0.0%) | 8 (0.9%) | 1 |
| Absent | 91 (100.0%) | 911 (99.1%) |  |
| FGFR3 Mutation |  |  |  |
| Present | 3 (3.3%) | 16 (1.7%) | 0.241 |
| Absent | 88 (96.7%) | 903 (98.3%) |  |
| FGFR4 Mutation |  |  |  |
| Present | 1 (1.1%) | 17 (1.8%) | 1 |
| Absent | 90 (98.9%) | 902 (98.2%) |  |
| HRAS Mutation |  |  |  |
| Present | 0 (0.0%) | 4 (0.4%) | 1 |
| Absent | 91 (100.0%) | 915 (99.6%) |  |
| JUN Mutation |  |  |  |
| Present | 0 (0.0%) | 13 (1.4%) | 0.6207 |
| Absent | 91 (100.0%) | 906 (98.6%) |  |
| KRAS Mutation |  |  |  |
| Present | 39 (42.9%) | 403 (43.9%) | 0.9428 |
| Absent | 52 (57.1%) | 516 (56.1%) |  |
| MAP2K1 Mutation |  |  |  |
| Present | 0 (0.0%) | 12 (1.3%) | 0.6152 |
| Absent | 91 (100.0%) | 907 (98.7%) |  |
| MAP2K2 Mutation |  |  |  |
| Present | 2 (2.2%) | 7 (0.8%) | 0.191 |
| Absent | 89 (97.8%) | 912 (99.2%) |  |
| MAP2K4 Mutation |  |  |  |
| Present | 1 (1.1%) | 18 (2.0%) | 1 |
| Absent | 90 (98.9%) | 901 (98.0%) |  |
| MAP3K1 Mutation |  |  |  |
| Present | 1 (1.1%) | 27 (2.9%) | 0.5044 |
| Absent | 90 (98.9%) | 892 (97.1%) |  |
| MAP3K13 Mutation |  |  |  |
| Present | 2 (2.2%) | 16 (1.7%) | 0.6731 |
| Absent | 89 (97.8%) | 903 (98.3%) |  |
| MAP3K14 Mutation |  |  |  |
| Present | 0 (0.0%) | 0 (0.0%) | 1 |
| Absent | 91 (100.0%) | 919 (100.0%) |  |
| MAPK1 Mutation |  |  |  |
| Present | 0 (0.0%) | 1 (0.1%) | 1 |
| Absent | 91 (100.0%) | 918 (99.9%) |  |
| MAPK3 Mutation |  |  |  |
| Present | 1 (1.1%) | 4 (0.4%) | 0.3769 |
| Absent | 90 (98.9%) | 915 (99.6%) |  |
| MAX Mutation |  |  |  |
| Present | 0 (0.0%) | 7 (0.8%) | 1 |
| Absent | 91 (100.0%) | 912 (99.2%) |  |
| MYC Mutation |  |  |  |
| Present | 0 (0.0%) | 7 (0.8%) | 1 |
| Absent | 91 (100.0%) | 912 (99.2%) |  |
| NF1 Mutation |  |  |  |
| Present | 2 (2.2%) | 46 (5.0%) | 0.3063 |
| Absent | 89 (97.8%) | 873 (95.0%) |  |
| NFATC2 Mutation |  |  |  |
| Present | 0 (0.0%) | 1 (0.1%) | 1 |
| Absent | 91 (100.0%) | 918 (99.9%) |  |
| NRAS Mutation |  |  |  |
| Present | 6 (6.6%) | 36 (3.9%) | 0.3449 |
| Absent | 85 (93.4%) | 883 (96.1%) |  |
| NTRK1 Mutation |  |  |  |
| Present | 2 (2.2%) | 14 (1.5%) | 0.6487 |
| Absent | 89 (97.8%) | 905 (98.5%) |  |
| NTRK2 Mutation |  |  |  |
| Present | 0 (0.0%) | 16 (1.7%) | 0.3866 |
| Absent | 91 (100.0%) | 903 (98.3%) |  |
| PAK1 Mutation |  |  |  |
| Present | 0 (0.0%) | 8 (0.9%) | 1 |
| Absent | 91 (100.0%) | 911 (99.1%) |  |
| PDGFRA Mutation |  |  |  |
| Present | 0 (0.0%) | 18 (2.0%) | 0.3962 |
| Absent | 91 (100.0%) | 901 (98.0%) |  |
| PDGFRB Mutation |  |  |  |
| Present | 0 (0.0%) | 16 (1.7%) | 0.3866 |
| Absent | 91 (100.0%) | 903 (98.3%) |  |
| PLA2G3 Mutation |  |  |  |
| Present | 0 (0.0%) | 0 (0.0%) | 1 |
| Absent | 91 (100.0%) | 919 (100.0%) |  |
| RAC1 Mutation |  |  |  |
| Present | 0 (0.0%) | 0 (0.0%) | 1 |
| Absent | 91 (100.0%) | 919 (100.0%) |  |
| RAC2 Mutation |  |  |  |
| Present | 0 (0.0%) | 0 (0.0%) | 1 |
| Absent | 91 (100.0%) | 919 (100.0%) |  |
| RAF1 Mutation |  |  |  |
| Present | 2 (2.2%) | 8 (0.9%) | 0.2255 |
| Absent | 89 (97.8%) | 911 (99.1%) |  |
| RASA1 Mutation |  |  |  |
| Present | 4 (4.4%) | 19 (2.1%) | 0.1454 |
| Absent | 87 (95.6%) | 900 (97.9%) |  |
| RPS6KA4 Mutation |  |  |  |
| Present | 3 (3.3%) | 17 (1.8%) | 0.4148 |
| Absent | 88 (96.7%) | 902 (98.2%) |  |
| RPS6KA6 Mutation |  |  |  |
| Present | 0 (0.0%) | 0 (0.0%) | 1 |
| Absent | 91 (100.0%) | 919 (100.0%) |  |
| RRAS Mutation |  |  |  |
| Present | 0 (0.0%) | 2 (0.2%) | 1 |
| Absent | 91 (100.0%) | 917 (99.8%) |  |
| RRAS2 Mutation |  |  |  |
| Present | 1 (1.1%) | 5 (0.5%) | 0.4333 |
| Absent | 90 (98.9%) | 914 (99.5%) |  |
| SOS1 Mutation |  |  |  |
| Present | 3 (3.3%) | 8 (0.9%) | 0.06862 |
| Absent | 88 (96.7%) | 911 (99.1%) |  |
| TGFB2 Mutation |  |  |  |
| Present | 0 (0.0%) | 1 (0.1%) | 1 |
| Absent | 91 (100.0%) | 918 (99.9%) |  |
| TGFBR1 Mutation |  |  |  |
| Present | 3 (3.3%) | 13 (1.4%) | 0.1685 |
| Absent | 88 (96.7%) | 906 (98.6%) |  |
| TGFBR2 Mutation |  |  |  |
| Present | 5 (5.5%) | 38 (4.1%) | 0.7334 |
| Absent | 86 (94.5%) | 881 (95.9%) |  |
| TP53 Mutation |  |  |  |
| Present | 71 (78.0%) | 675 (73.4%) | 0.4111 |
| Absent | 20 (22.0%) | 244 (26.6%) |  |

| TRAF2 Mutation |  |  |  |
| --- | --- | --- | --- |
| Present | 0 (0.0%) | 10 (1.1%) | 1 |
| Absent | 91 (100.0%) | 909 (98.9%) |  |

**Table S12.** Comparison of Late-Onset Hispanic/Latino (H/L) versus Late-Onset Non-Hispanic White (NHW) Patients Not Treated with FOLFOX.

| MAPK Pathway |  |  |  |
| --- | --- | --- | --- |
| Gene | Late-Onset Hispanic/Latino<br>Not Treated with FOLFOX<br>n (%) | Late-Onset NHW<br>Not Treated with FOLFOX<br>n (%) | p-value |
| ACVR1 Mutation |  |  |  |
| Present | 0 (0.0%) | 8 (1.2%) | 1 |
| Absent | 50 (100.0%) | 645 (98.8%) |  |
| AKT1 Mutation |  |  |  |
| Present | 2 (4.0%) | 19 (2.9%) | 0.6558 |
| Absent | 48 (96.0%) | 634 (97.1%) |  |
| AKT2 Mutation |  |  |  |
| Present | 0 (0.0%) | 12 (1.8%) | 1 |
| Absent | 50 (100.0%) | 641 (98.2%) |  |
| AKT3 Mutation |  |  |  |
| Present | 2 (4.0%) | 12 (1.8%) | 0.2621 |
| Absent | 48 (96.0%) | 641 (98.2%) |  |
| BRAF Mutation |  |  |  |
| Present | 8 (16.0%) | 93 (14.2%) | 0.8947 |
| Absent | 42 (84.0%) | 560 (85.8%) |  |
| CACNA1H Mutation |  |  |  |
| Present | 0 (0.0%) | 1 (0.2%) | 1 |
| Absent | 50 (100.0%) | 652 (99.8%) |  |
| CACNA2D1 Mutation |  |  |  |
| Present | 0 (0.0%) | 0 (0.0%) | 1 |
| Absent | 50 (100.0%) | 653 (100.0%) |  |
| CACNB1 Mutation |  |  |  |
| Present | 0 (0.0%) | 0 (0.0%) | 1 |
| Absent | 50 (100.0%) | 653 (100.0%) |  |
| CDC42 Mutation |  |  |  |
| Present | 1 (2.0%) | 2 (0.3%) | 0.1988 |
| Absent | 49 (98.0%) | 651 (99.7%) |  |
| CRKL Mutation |  |  |  |
| Present | 0 (0.0%) | 8 (1.2%) | 1 |
| Absent | 50 (100.0%) | 645 (98.8%) |  |
| DAXX Mutation |  |  |  |
| Present | 1 (2.0%) | 13 (2.0%) | 1 |
| Absent | 49 (98.0%) | 640 (98.0%) |  |
| DUSP4 Mutation |  |  |  |
| Present | 0 (0.0%) | 6 (0.9%) | 1 |
| Absent | 50 (100.0%) | 647 (99.1%) |  |
| EGFR Mutation |  |  |  |
| Present | 3 (6.0%) | 24 (3.7%) | 0.4311 |
| Absent | 47 (94.0%) | 629 (96.3%) |  |
| FGF13 Mutation |  |  |  |
| Present | 0 (0.0%) | 0 (0.0%) | 1 |
| Absent | 50 (100.0%) | 653 (100.0%) |  |
| FGF19 Mutation |  |  |  |
| Present | 2 (4.0%) | 5 (0.8%) | 0.08273 |
| Absent | 48 (96.0%) | 648 (99.2%) |  |
| FGF3 Mutation |  |  |  |
| Present | 0 (0.0%) | 14 (2.1%) | 0.6146 |
| Absent | 50 (100.0%) | 639 (97.9%) |  |
| FGF4 Mutation |  |  |  |
| Present | 0 (0.0%) | 4 (0.6%) | 1 |
| Absent | 50 (100.0%) | 649 (99.4%) |  |
| FGFR1 Mutation |  |  |  |
| Present | 0 (0.0%) | 16 (2.5%) | 0.62 |
| Absent | 50 (100.0%) | 637 (97.5%) |  |
| FGFR2 Mutation |  |  |  |
| Present | 1 (2.0%) | 11 (1.7%) | 1 |
| Absent | 49 (98.0%) | 642 (98.3%) |  |
| FGFR3 Mutation |  |  |  |
| Present | 2 (4.0%) | 19 (2.9%) | 0.6558 |
| Absent | 48 (96.0%) | 634 (97.1%) |  |
| FGFR4 Mutation |  |  |  |
| Present | 1 (2.0%) | 22 (3.4%) | 1 |
| Absent | 49 (98.0%) | 631 (96.6%) |  |
| HRAS Mutation |  |  |  |
| Present | 1 (2.0%) | 6 (0.9%) | 0.4047 |
| Absent | 49 (98.0%) | 647 (99.1%) |  |
| JUN Mutation |  |  |  |
| Present | 0 (0.0%) | 21 (3.2%) | 0.3898 |
| Absent | 50 (100.0%) | 632 (96.8%) |  |
| KRAS Mutation |  |  |  |
| Present | 20 (40.0%) | 279 (42.7%) | 0.8202 |
| Absent | 30 (60.0%) | 374 (57.3%) |  |
| MAP2K1 Mutation |  |  |  |
| Present | 0 (0.0%) | 10 (1.5%) | 1 |
| Absent | 50 (100.0%) | 643 (98.5%) |  |
| MAP2K2 Mutation |  |  |  |
| Present | 1 (2.0%) | 10 (1.5%) | 0.5585 |
| Absent | 49 (98.0%) | 643 (98.5%) |  |
| MAP2K4 Mutation |  |  |  |
| Present | 1 (2.0%) | 17 (2.6%) | 1 |
| Absent | 49 (98.0%) | 636 (97.4%) |  |
| MAP3K1 Mutation |  |  |  |
| Present | 1 (2.0%) | 31 (4.7%) | 0.7202 |
| Absent | 49 (98.0%) | 622 (95.3%) |  |
| MAP3K13 Mutation |  |  |  |
| Present | 0 (0.0%) | 15 (2.3%) | 0.6164 |
| Absent | 50 (100.0%) | 638 (97.7%) |  |
| MAP3K14 Mutation |  |  |  |
| Present | 0 (0.0%) | 0 (0.0%) | 1 |
| Absent | 50 (100.0%) | 653 (100.0%) |  |
| MAPK1 Mutation |  |  |  |
| Present | 0 (0.0%) | 6 (0.9%) | 1 |
| Absent | 50 (100.0%) | 647 (99.1%) |  |
| MAPK3 Mutation |  |  |  |
| Present | 0 (0.0%) | 5 (0.8%) | 1 |
| Absent | 50 (100.0%) | 648 (99.2%) |  |
| MAX Mutation |  |  |  |
| Present | 0 (0.0%) | 5 (0.8%) | 1 |
| Absent | 50 (100.0%) | 648 (99.2%) |  |
| MYC Mutation |  |  |  |
| Present | 2 (4.0%) | 9 (1.4%) | 0.1809 |
| Absent | 48 (96.0%) | 644 (98.6%) |  |
| NF1 Mutation |  |  |  |
| Present | 2 (4.0%) | 44 (6.7%) | 0.7642 |
| Absent | 48 (96.0%) | 609 (93.3%) |  |
| NFATC2 Mutation |  |  |  |
| Present | 0 (0.0%) | 0 (0.0%) | 1 |
| Absent | 50 (100.0%) | 653 (100.0%) |  |
| NRAS Mutation |  |  |  |
| Present | 3 (6.0%) | 19 (2.9%) | 0.2011 |
| Absent | 47 (94.0%) | 634 (97.1%) |  |
| NTRK1 Mutation |  |  |  |
| Present | 2 (4.0%) | 14 (2.1%) | 0.3169 |
| Absent | 48 (96.0%) | 639 (97.9%) |  |
| NTRK2 Mutation |  |  |  |
| Present | 3 (6.0%) | 9 (1.4%) | 0.0472 |
| Absent | 47 (94.0%) | 644 (98.6%) |  |
| PAK1 Mutation |  |  |  |
| Present | 0 (0.0%) | 6 (0.9%) | 1 |
| Absent | 50 (100.0%) | 647 (99.1%) |  |
| PDGFRA Mutation |  |  |  |
| Present | 0 (0.0%) | 24 (3.7%) | 0.4049 |
| Absent | 50 (100.0%) | 629 (96.3%) |  |
| PDGFRB Mutation |  |  |  |
| Present | 3 (6.0%) | 16 (2.5%) | 0.1466 |
| Absent | 47 (94.0%) | 637 (97.5%) |  |
| PLA2G3 Mutation |  |  |  |
| Present | 0 (0.0%) | 1 (0.2%) | 1 |
| Absent | 50 (100.0%) | 652 (99.8%) |  |
| RAC1 Mutation |  |  |  |
| Present | 0 (0.0%) | 3 (0.5%) | 1 |
| Absent | 50 (100.0%) | 650 (99.5%) |  |
| RAC2 Mutation |  |  |  |
| Present | 1 (2.0%) | 3 (0.5%) | 0.256 |
| Absent | 49 (98.0%) | 650 (99.5%) |  |
| RAF1 Mutation |  |  |  |
| Present | 1 (2.0%) | 19 (2.9%) | 1 |
| Absent | 49 (98.0%) | 634 (97.1%) |  |
| RASA1 Mutation |  |  |  |
| Present | 1 (2.0%) | 24 (3.7%) | 1 |
| Absent | 49 (98.0%) | 629 (96.3%) |  |
| RPS6KA4 Mutation |  |  |  |
| Present | 3 (6.0%) | 34 (5.2%) | 0.7411 |
| Absent | 47 (94.0%) | 619 (94.8%) |  |
| RPS6KA6 Mutation |  |  |  |
| Present | 0 (0.0%) | 0 (0.0%) | 1 |
| Absent | 50 (100.0%) | 653 (100.0%) |  |
| RRAS Mutation |  |  |  |
| Present | 0 (0.0%) | 8 (1.2%) | 1 |
| Absent | 50 (100.0%) | 645 (98.8%) |  |
| RRAS2 Mutation |  |  |  |
| Present | 2 (4.0%) | 3 (0.5%) | 0.04318 |
| Absent | 48 (96.0%) | 650 (99.5%) |  |
| SOS1 Mutation |  |  |  |
| Present | 0 (0.0%) | 15 (2.3%) | 0.6164 |
| Absent | 50 (100.0%) | 638 (97.7%) |  |
| TGFB2 Mutation |  |  |  |
| Present | 0 (0.0%) | 0 (0.0%) | 1 |
| Absent | 50 (100.0%) | 653 (100.0%) |  |
| TGFBR1 Mutation |  |  |  |
| Present | 1 (2.0%) | 12 (1.8%) | 1 |
| Absent | 49 (98.0%) | 641 (98.2%) |  |
| TGFBR2 Mutation |  |  |  |
| Present | 3 (6.0%) | 46 (7.0%) | 1 |
| Absent | 47 (94.0%) | 607 (93.0%) |  |
| TP53 Mutation |  |  |  |
| Present | 36 (72.0%) | 445 (68.1%) | 0.684 |
| Absent | 14 (28.0%) | 208 (31.9%) |  |
| TRAF2 Mutation |  |  |  |
| Present | 1 (2.0%) | 15 (2.3%) | 1 |
| Absent | 49 (98.0%) | 638 (97.7%) |  |

**Table S12.**
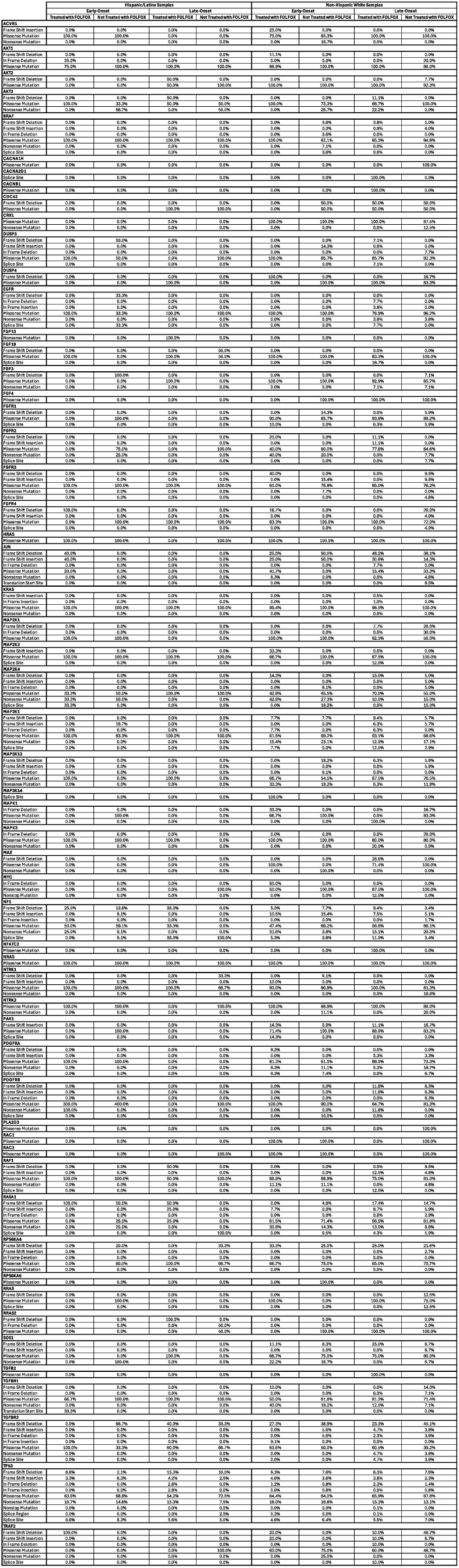
Distribution of MAPK Pathway Mutation Classes by Ancestry, Age at Diagnosis, and FOLFOX Exposure in Colorectal Cancer. This supplementary table details the spectrum of somatic mutation classes observed in MAPK signaling pathway genes across colorectal cancer subgroups stratified by ancestral background (Hispanic/Latino [H/L] vs. non-Hispanic White [NHW]), age of onset (early-onset vs. late-onset), and FOLFOX chemotherapy status (treated vs. not treated). Mutation categories include missense substitutions, nonsense mutations, frameshift insertions and deletions, in-frame insertions/deletions, splice-site and splice-region variants, nonstop mutations, and translation start-site alterations. For each subgroup, percentages reflect the relative contribution of each mutation class to the total number of MAPK pathway alterations identified within that stratum. By comparing mutation-type distributions across demographic and treatment-defined cohorts, this table provides insight into how ancestry, age at diagnosis, and chemotherapy exposure influence the qualitative nature of MAPK pathway genomic disruption, complementing gene-level frequency and pathway-level analyses presented in the main text.

**Figure S1.**
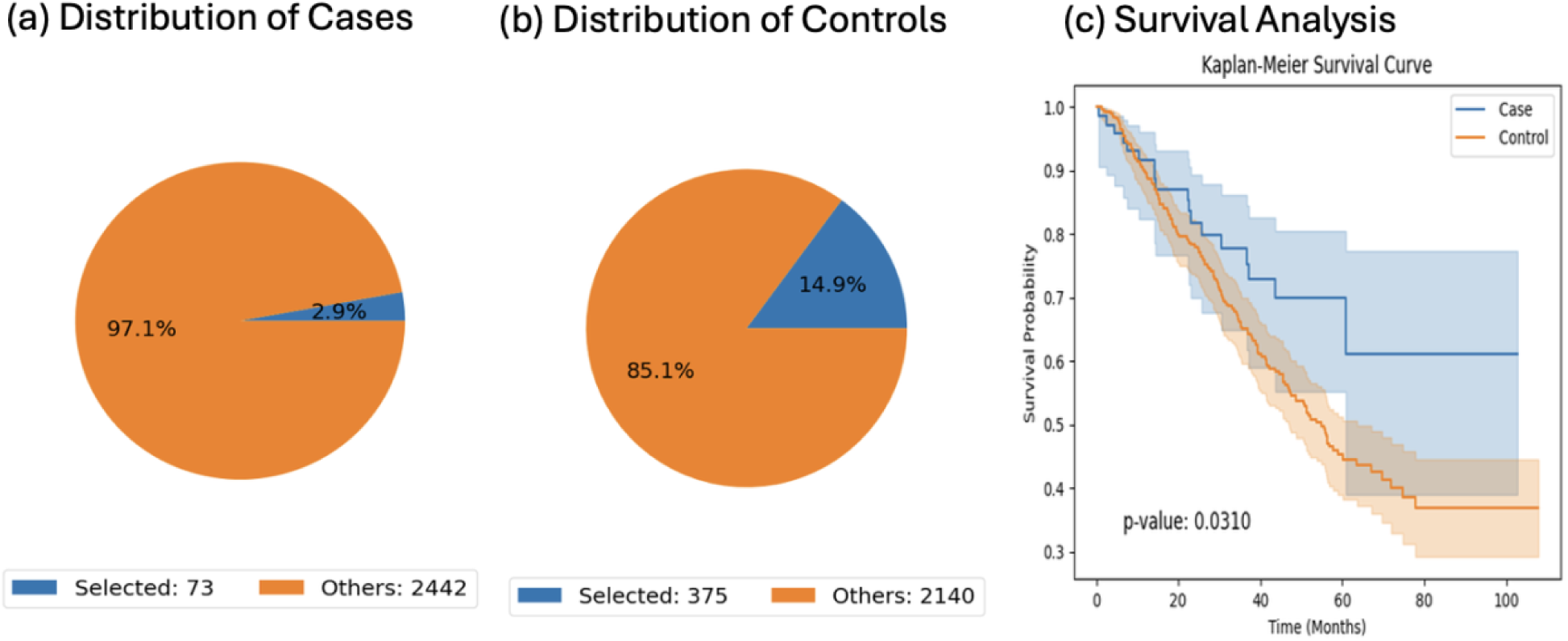
AI-directed cohort selection and survival comparison for early-onset treated Hispanic/Latino (H/L) and Non-Hispanic White (NHW) colorectal cancer (CRC) patients. This figure illustrates how the conversational AI platform identified matched case and control groups using user-defined clinical criteria and subsequently evaluated differences in overall survival (OS). (a) Within early-onset H/L CRC cases treated with FOLFOX, the AI tool isolated 73 eligible samples (2.9% of all available records), while the remaining 97.1% were not selected based on inclusion parameters. (b) Among early-onset NHW CRC controls treated with FOLFOX, 375 patients (14.9% of all available records) met the same selection rules, with 85.1% of records excluded. (c) Kaplan-Meier analysis visualizes OS differences between case and control cohorts, demonstrating a statistically significant survival distinction (log-rank p = 0.0310), with the case cohort displaying improved outcomes over time. Confidence intervals (95%) are shown for each curve, reflecting uncertainty around survival estimates. The figure highlights the ability of AI-driven cohort generation to support survival comparisons across demographic groups sharing matched clinical features.

**Figure S2.**
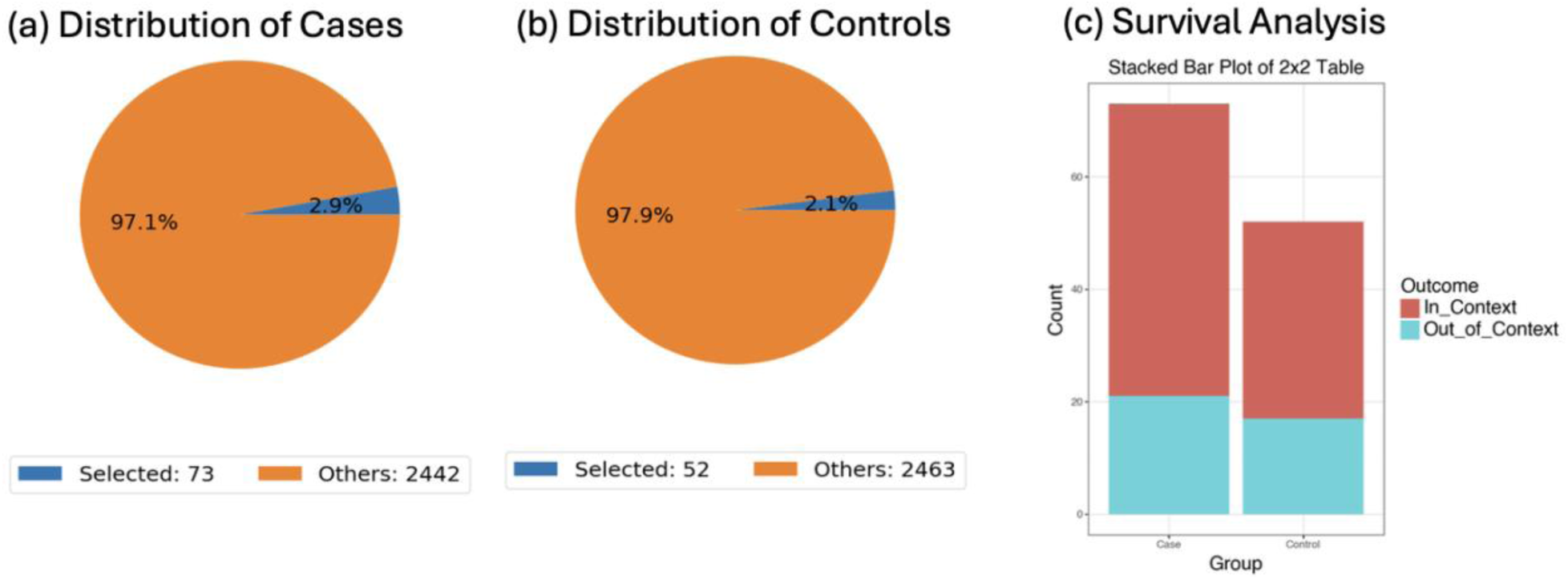
AI-driven analysis of MSI stability patterns among early-onset Hispanic/Latino colorectal cancer patients stratified by FOLFOX treatment exposure. This figure demonstrates how conversational querying was used to isolate and compare two subgroups within early-onset H/L colorectal cancer (CRC): individuals treated with FOLFOX (case cohort; n = 73) and those who did not receive FOLFOX (control cohort; n = 52). Panels (a) and (b) show the distribution of selected samples (blue) relative to the total available dataset (orange) for each subgroup, highlighting the proportion of patients meeting the defined criteria. Panel (c) presents a stacked bar visualization of a 2×2 odds ratio test evaluating whether microsatellite stability (MSI-stable genotype) differed between treatment groups. Fisher’s exact test revealed no significant association between FOLFOX exposure and MSI stability status (p = 0.785), with similar proportions of MSI-stable tumors observed in both groups. These findings suggest that, within early-onset H/L CRC, MSI stability patterns remain consistent regardless of FOLFOX treatment, and further demonstrate the utility of AI-guided cohort construction for efficient molecular comparison and validation across clinically defined strata.

**Figure S3.**
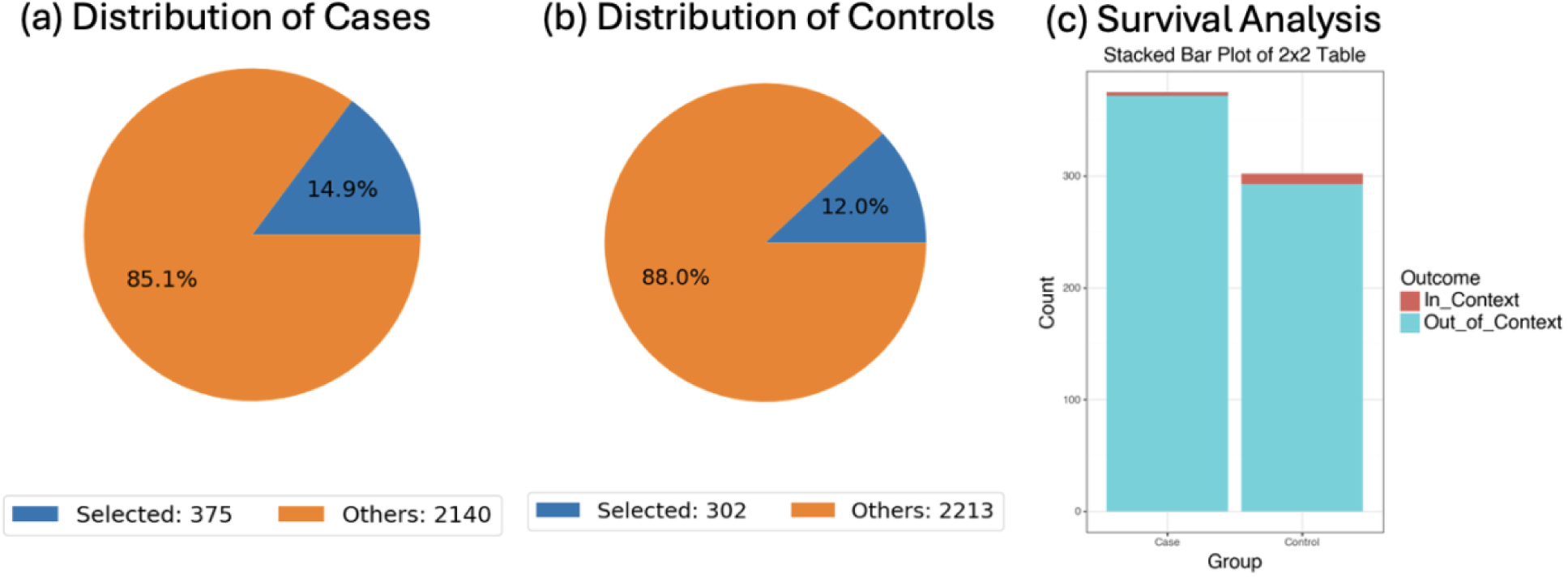
AI-guided assessment of AKT3 mutation frequency among early-onset Non-Hispanic White (NHW) colorectal cancer patients stratified by FOLFOX exposure. This figure illustrates how conversational filtering was used to compare AKT3 mutation prevalence between early-onset NHW CRC patients treated with FOLFOX (case cohort; n = 375) and those who did not receive FOLFOX (control cohort; n = 302). Panels (a) and (b) show pie charts representing the proportion of selected samples meeting the AKT3 mutation criterion (“in-context”; blue) versus all other samples (“out-of-context”; orange) within each subgroup. Panel (c) displays a stacked bar visualization of the odds ratio analysis, contrasting AKT3-mutated and non-mutated cases across treatment strata. Fisher’s exact testing revealed no statistically significant association between FOLFOX exposure and AKT3 mutation status (p = 0.065), although mutation frequency was numerically lower in the treated cohort. These results suggest that AKT3 mutation prevalence remains relatively consistent regardless of chemotherapy exposure in early-onset NHW CRC and demonstrate the utility of AI-based cohort selection for rapid, treatment-specific genomic comparisons.

**Figure S4.**
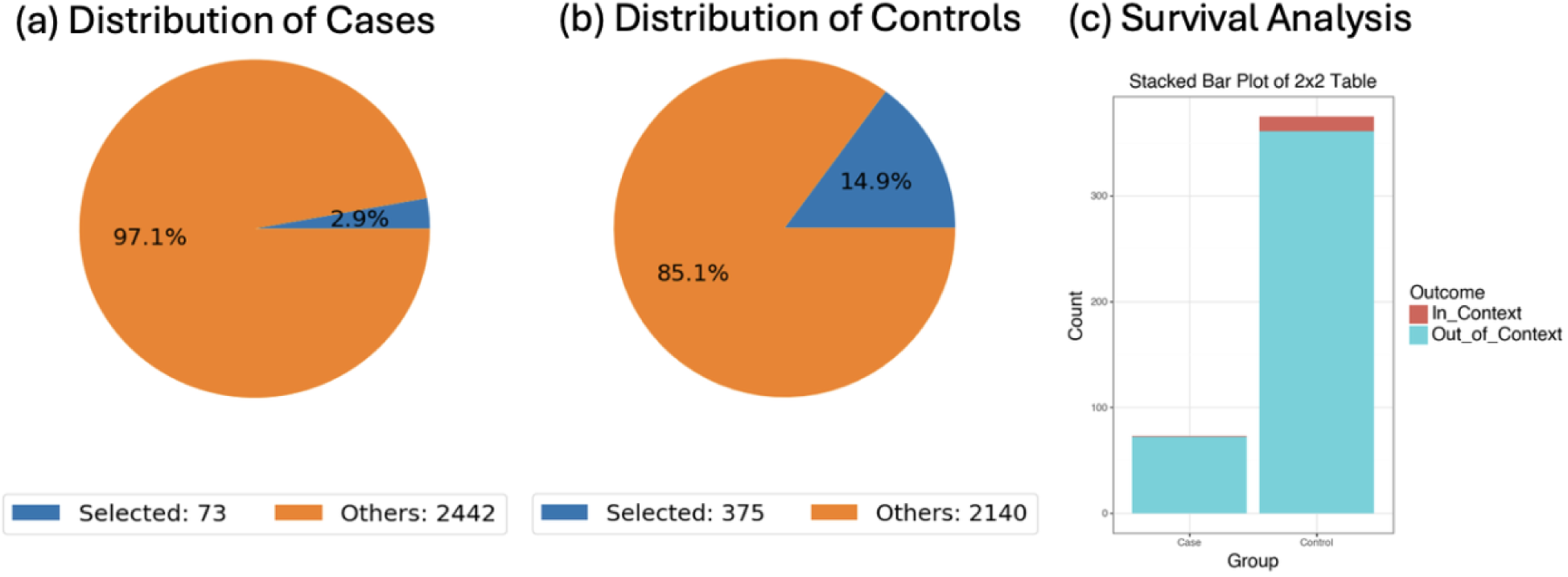
AI-supported evaluation of PDGFRB mutation prevalence in early-onset colorectal cancer patients treated with FOLFOX across Hispanic/Latino (H/L) and Non-Hispanic White (NHW) populations. This figure shows how the conversational AI framework was used to generate matched cohorts for comparing PDGFRB mutation rates between early-onset H/L CRC patients treated with FOLFOX (case cohort; n = 73) and early-onset NHW patients also treated with FOLFOX (control cohort; n = 375). Pie charts in panels (a) and (b) depict the proportion of cases meeting the PDGFRB mutation criterion (“selected”) relative to the total number of available samples within each subgroup. Panel (c) presents a stacked bar visualization of the odds ratio analysis, comparing the number of PDGFRB-mutated and non-mutated samples across the two ancestry groups. Fisher’s exact testing did not reveal a statistically significant difference in PDGFRB mutation prevalence between treated H/L and NHW patients (p = 0.502), although mutation frequency was numerically lower in the H/L cohort. These findings suggest broadly comparable PDGFRB mutation rates across FOLFOX-treated early-onset patients irrespective of ancestry, and highlight the value of AI-driven cohort selection in enabling rapid, ancestry-aware molecular comparisons.

**Figure S5.**
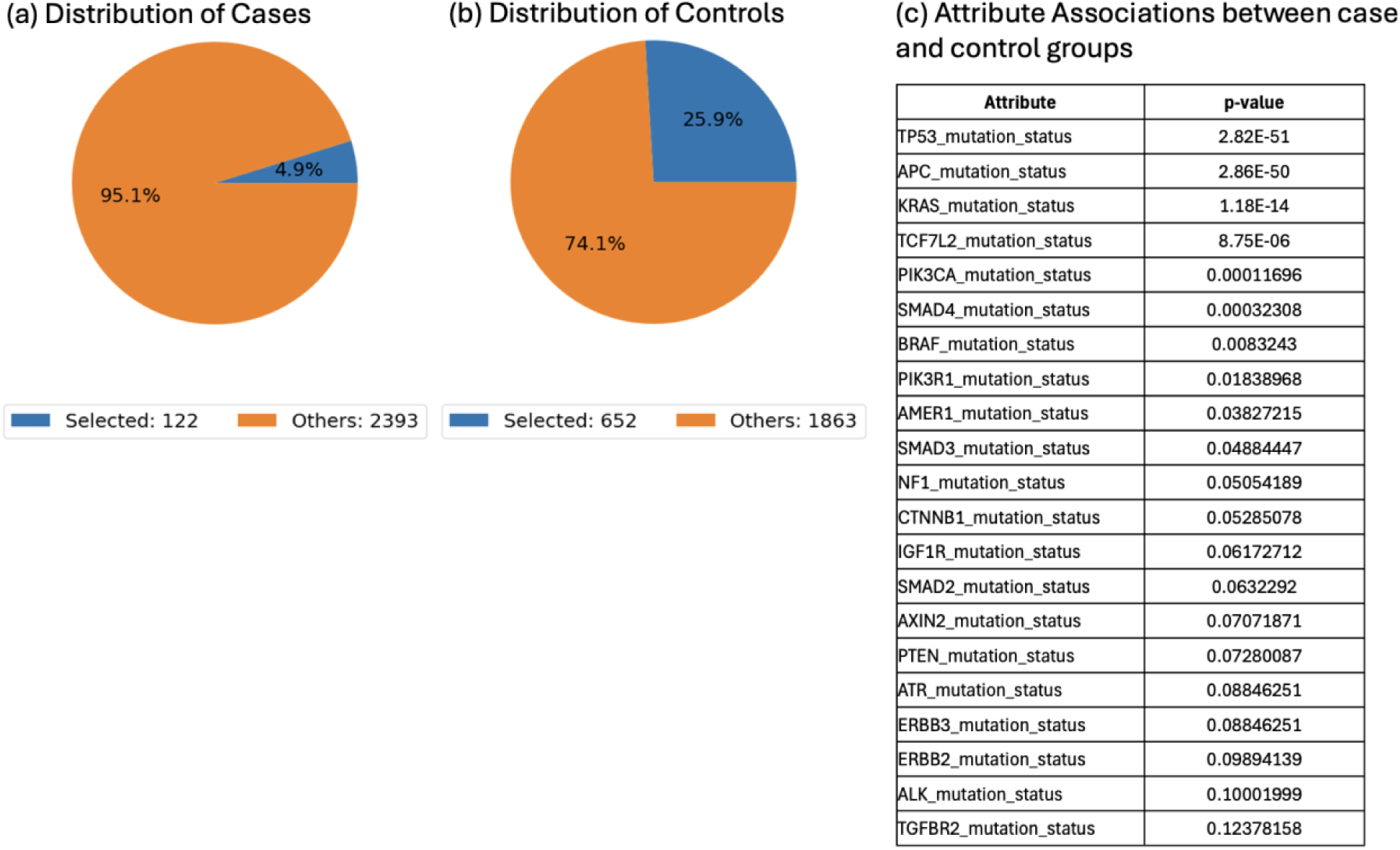
AI-enabled comparison of MAPK-altered early-onset colorectal cancer tumors reveals ancestry-linked molecular differences. This figure demonstrates how the conversational AI platform was used to evaluate mutation-level attributes distinguishing early-onset Hispanic/Latino (H/L) colorectal cancer (CRC) tumors from early-onset Non-Hispanic White (NHW) tumors, restricted to MAPK-altered samples. Pie charts in panels (a) and (b) display the proportion of selected (MAPK-altered) samples relative to all available cases within each ancestry group, showing a larger selected fraction among NHW patients (652/2,515; 25.9%) compared with H/L patients (122/2,515; 4.9%). Panel (c) summarizes mutation attributes that differed significantly between groups based on AI-driven statistical prioritization. The most strongly associated alterations included TP53, APC, KRAS, TCF7L2, and PIK3CA (all p < 0.001), followed by others such as SMAD4, BRAF, and NF1. The Figure highlight pronounced ancestry-associated molecular heterogeneity within MAPK-altered early-onset CRC and illustrate how an interactive AI framework can rapidly surface genomic features driving differences between clinical populations.

